# Dietary protein consumption profiles show contrasting impacts on environmental and health indicators

**DOI:** 10.1101/2022.07.07.22277350

**Authors:** Elie Perraud, Juhui Wang, Marion Salomé, François Mariotti, Emmanuelle Kesse-Guyot

**Affiliations:** Université Paris-Saclay, AgroParisTech, INRAE, UMR PNCA, 75005, Paris, France; Sorbonne Paris Nord University, Institut National de la Santé et de la Recherche Médicale (INSERM), Institut National de Recherche pour l’Agriculture, l’Alimentation et l’Environnement (INRAE), Conservatoire National des Arts et Métiers (CNAM), Nutritional Epidemiology Research Team (EREN), Epidemiology and Statistics Research Center–University of Paris (CRESS), Bobigny, France

## Abstract

Patterns of protein intake are strong characteristics of diets, and protein sources have been linked to the environmental and nutrition/health impacts of diets. However, few studies have worked on protein profiles, and most of them have focused on specific diets like vegetarian or vegan diets. Furthermore, the description of the environmental impact of diets has often been limited to greenhouse gas emissions (GHGe) and land use. This paper analyzes the alignment of environmental pressures and nutritional impacts in a diversity of representative protein profiles of a western population.

Using data from a representative survey in France (INCA3, n = 1,125), we identified protein profiles using hierarchical ascendant classification on protein intake (g) from main protein sources (refined grains, whole grains, dairy, eggs, ruminant meat, poultry, pork, processed meat, fish, fruits & vegetables, pulses). We assessed their diet quality using 6 dietary scores, including assessment of long-term risk for health, and associated 14 environmental pressure indicators using the Agribalyse database completed by the SHARP database for GHGe.

Five protein profiles were identified according to the high contributions of ruminant meat, pork, poultry, fish, or, conversely, as low contribution from meat. The profile including the lowest protein from meat had the lowest impact on almost all environmental indicators and had the lowest long-term risk. Conversely, the profile with high protein from ruminant-based foods had the highest pressures on most environmental indicators, including GHGe.

We found that the protein profile with low contribution from meat has great potential for human health and environment preservation. Shifting a large part of the population toward this profile could be an easy first step toward building a more sustainable diet.

**Graphical abstract:** 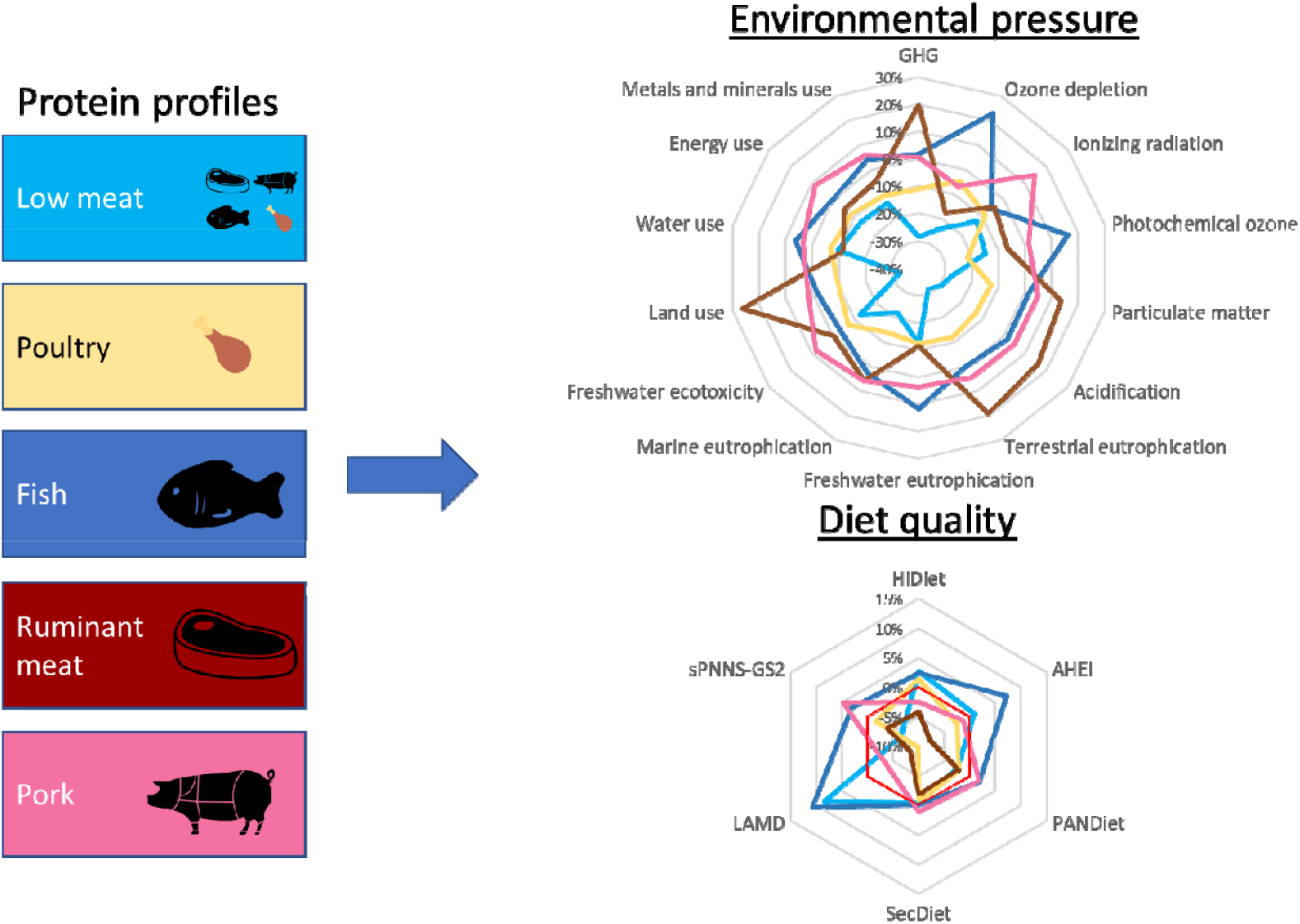

**Environmental pressure by profile:** All data are in percentage compared to the mean value of the population. GHG is the emission of greenhouse gases measured in kg CO_2_ eq. The ozone depletion is in kg CFC-11eq. The photochemical ozone formation is in kg of Non-Methane Volatile Organic Compounds eq. Particulate matter is in kg of PM_2.5_emitted. The acidification is in mol H^+^ eq. The terrestrial eutrophication is in mol N eq, the freshwater eutrophication is in kg P eq, and the marine eutrophication is in kg N eq. The freshwater ecotoxicity is based on the USEtox model. Land use is in kg C deficit, water use in m^3^, fossils resource use in MJ, and Metals and minerals use in kg SB eq.

**Nutritional and health indicators by profile:** All data are in percentage of difference to the mean value of the population. The HiDiet was used to assess the diet impact on long-term mortality and morbidity (variation between -1 and 1). The Alternative Healthy Eating Index (AHEI-2010) is a modified Healthy Eating Index, assessing the adherence to *Dietary Guidelines for Americans*, improving target food choices and macronutrient sources associated with reduced chronic disease risk (maxpoint = 100). The PANDiet evaluates the probability of adequate nutrient intake (maxpoint = 100). The SecDiet evaluates the nutrient risk of overt deficiency (maxpoint = 1). The Literature-Based Adherence Score to the Mediterranean Diet (LAMD) assesses adherence to the Mediterranean diet (maxpoint = 16). sPNNS-GS2 the adherence to the French Food-based Dietary Guidelines (maxpoint = 10.5).

## 1 Introduction

Chronic diseases strongly influenced by diets are leading causes of morbidity and mortality around the world (GBD 2013 Mortality and Causes of Death Collaborators, 2015). The food systems underlying these diets are responsible for a third of global greenhouse gas emissions (GHGe) (Crippa et al., 2021) and influence several other environmental indicators (Willett et al., 2019). More and more studies are showing the significant environmental pressures of meat and dairy food production compared to plant food production (González et al., 2011; Nijdam et al., 2012). For example, beef produces 7.1 g protein /kg CO_2_ eq compared to beans producing 246 g protein/kg CO_2_ eq (González et al., 2011). The same relationship was seen for land use with beef protein needing between 15 m^2^. y^-1^.kg^-1^ in industrial systems to 420 m^2^. y^-1^ .kg^-1^ for extensive pastoral systems in contrast to pulses needing between 3 to 8 m^2^.y.kg^-1^ for the production of 1kg of protein (Nijdam et al., 2012). The amount of water used for protein production shows similar extremes of variation, with beef using 728 L/100 grams of protein, and peas and other pulses using around 200 L/100 grams of protein (Ritchie and Roser, 2020).

Protein profiles largely define diets (De Gavelle et al., 2018; Marini et al., 2021), since protein food sources contribute other nutrients that tend to cluster together as a “protein package”. They are also associated with other types of foods as part of dietary patterns (De Gavelle et al., 2018; de Gavelle et al., 2019; Mariotti, 2019). Protein profiles in diets are thus key to both environmental and health issues (Clarys et al., 2014; González-García et al., 2018; Seconda et al., 2018).

Several diets have been modeled to improve the population’s health while concomitantly considering the environmental issues. These diets display drastic differences in protein profile compared to current westernized diets (Kramer et al., 2017; Seconda et al., 2021; Willett et al., 2019; Wilson et al., 2019). For example, the EAT-Lancet diet includes higher consumption of fruit, vegetable, and whole grains, and is very low in red and processed meat (Willett et al., 2019). This shift in plant and animal sources tend to reverse the plant-animal protein ratio compared to the one observed in modern diets. In the EPIC Oxford study, when compared to a high meat diet, a low meat diet (<50 g/day), was associated with a lower carbon footprint by 920 kg CO_2_eq every year (Scarborough et al., 2014). Similarly, the association between the consumption of food group sources of protein and long-term health has been widely studied (Micha et al., 2017a; Turner and Lloyd, 2017). For example, red and processed meats have been associated with several non-communicable diseases like colorectal carcinoma, type 2 diabetes, and cardiovascular disease (Boada et al., 2016; Micha et al., 2010). In contrast, plant protein intakes have been associated with long-term health benefits (Mariotti, 2019; Naghshi et al., 2020). Thus, compliance with dietary guidelines (Herforth et al., 2019), including those from France (Chaltiel et al., 2019), implies a shift of the dietary protein patterns to diets that include lowering meat consumption in some subgroups of the population in Western countries (Brunin et al., 2021; Dagevos, 2021). Many healthy dietary patterns promoted via states official national guidelines include specific recommendations regarding protein sources (Canada, 2020; Chang and Koegel, 2017; ODPHP, 2021). However, contemporary data specifying the link between diet and its impact on both the health and environment is primarily based on single food groups rather than dietary profiles, and does not focus specifically on protein patterns.

It is important to note that studies that intended to depict the sustainability of diet profiles rather than of isolated food groups have focused mostly on restrictive diets consumed by a minority, such as vegetarian or vegan diets, which were compared with the whole omnivore population (Chai et al., 2019; Fresán and Sabaté, 2019; Rabès et al., 2020; Rosi et al., 2017). Still, no literature to date has studied the intermediate proteins profiles that represent most of the profiles seen in the population.

Furthermore, while several criteria are used to estimate the nutritional or health value of diets, very few environmental indicators are reported. Indeed, most studies limit their indicators to GHGe and land occupation (Aleksandrowicz et al., 2016; Jones et al., 2016). Although these two indicators are important, often a markers of the whole environmental footprint (Kramer et al., 2017), they remain limited in depicting the environmental impact along several dimensions.

In order to inform the future diet transition, it is essential to advance accurate knowledge on the relationship between dietary profiles as naturally occurring in the diversity of people diets, and environmental pressures, and nutritional/health quality. In the present study, we aimed to analyze the potential alignment between environmental pressures and nutritional/health impacts of the main protein profiles identified in a French representative population.

## 2 Methods

### 2.1 Studied population and dietary data

This study used data from the third individual and national study on food consumption survey (INCA3) performed in France between 2014 and 2015 (Dubuisson et al., 2019; French Agency for Food, Environmental and Occupational Health Safety (ANSES), 2017). Participants identified as under-reporters were excluded using the basal metabolic rate as estimated by the Henry equation (Henry, 2005), using the cut-off values recommended by Black (Black, 2000). Since nutrient requirements for older adults differ from younger ones, they were excluded (above 54 years old for women and 64 years old for men). The final sample contained 1125 adults, including 564 men and 561 women.

Data were collected by three non-consecutive 24h-dietary recalls (two during the week and one at the weekend) over a 3-week period. The nutrient content values of the food were extracted from the 2016 database of the *French Centre d’Information sur la Qualité des Aliments* (CIQUAL) (ANSES, 2016).

### 2.2 Diet quality assessment

Six scores were used to compare the different impacts of the diet on health and nutritional status.

Three of them evaluated the adherence to a specific diet or dietary recommendations were calculated. The Literature-Based Adherence Score to the Mediterranean Diet (LAMD) assesses adherence to the Mediterranean diet. The LAMD score includes three negative components (meat, dairy products, and alcohol) and six positive components (fruits, vegetables, legumes, grains, fish, and olive oil). The consumption of each food group is classified into three categories using cutoff points (Supplemental Table 1)(Sofi et al., 2014). The Alternative Healthy Eating Index (AHEI-2010) is a modified Healthy Eating Index, assessing the adherence to *Dietary Guidelines for Americans*, improving target food choices and macronutrient sources associated with reduced chronic disease risk (Chiuve et al., 2012). Since information on *trans* fatty acids was not available, only ten parameters were considered, namely vegetables, fruit, whole grains, sugar-sweetened beverages and fruit juice, nuts and legumes, red/processed meat, long-chain (n-3) fatty acids (EPA + DHA), polyunsaturated fatty acids (PUFA), sodium, and alcohol. All AHEI-2010 components were scored from 0 (worst) to 10 (best), resulting in a total AHEI-2010 score ranging from 0 (non-adherence) to 100 (perfect adherence). The scoring criteria are described in Supplemental Table 2. The sPNNS-GS2 (simplified *Programme National Nutrition Santé* Guideline Score 2) assesses the adherence to the French Food-based Dietary Guidelines (Chaltiel et al., 2019). Components, scoring and weighting are shown in Supplemental Table 3.

The PANDiet evaluates the probability of adequate nutrient intake. This score is a 100-point probabilistic score evaluating adequate overall nutrient intake. It combines an adequacy sub-score and a moderation sub-score. The adequacy sub-score is calculated as the average probability of adequacy of nutrients for which the usual intake should be above a reference value, multiplied by 100. For a given nutrient, the probability of adequacy was determined from either the Estimated Average Requirement or the Adequate Intake, and its variability. The moderation sub-score (MS) was calculated as the average of probabilities of inadequacy of six nutrients for which an upper bound reference value exists, together with penalty values. For the six nutrients, the probability of adequacy was calculated, using the upper bound of the acceptable macronutrient distribution range (De Gavelle et al., 2018).

The SecDiet evaluates nutrient deficiency risk. This score is based on the intake of twelve critical nutrients for nutritional risk of overt deficiency (Salomé et al., 2021). For each nutrient, we calculated the probability of having sufficient-enough intake to avoid overt nutrient deficiency. We used the probability distribution of the standard normal distribution for nutrient requirements while taking into account the mean intake, the day-to-day intake variability, the inter-individual variability and the nutrient deficiency threshold.

The HiDiet was used to assess the diet impact on long-term mortality and morbidity. The HiDiet score is based on the principle of the Comparative Risk Assessment but applied to the risk of one individual and so is an individual version of the EpiDiet (Evaluate the Potential Impact of a Diet) (Dussiot et al., 2022; Kesse-Guyot et al., 2020). The conceptual basis and methodological foundation of the two models are the same which are laid out in the Comparative Risk Assessment framework. It allows for the evaluation of the potential impacts of dietary changes on the long-term morbidity and mortality caused by some diet-related diseases.

In this study, we set up the HiDiet model with values reported in a series of validated international meta-analyses published by a European team (Bechthold et al., 2019; Schwingshackl et al., 2019, 2018, 2017a, 2017b). As in our previous study (Dussiot et al., 2022), we selected 12 diet-related factors (including consumption of fruit, vegetables, nuts or seeds, whole grains, unprocessed red meats, processed meats, sugar-sweetened beverages, fish, dairy products, eggs, refined grains, and legumes) and 4 diet-related diseases (including coronary heart disease, stroke, type 2 diabetes, and colorectal cancer).

The reference population considered was the French adult population (18 and 64 years old) in 2014, stratified into sub-populations by 5-year age-bands and sex. The population demographics and national disease-specific deaths were provided by the National Institute of Statistics and Economic Studies (INSEE, 2017), and by the Epidemiological Centre on Medical Causes of Death (CépiDc, 2018), respectively. For each individual, two dietary scenarios were built. The baseline scenario corresponds to the average daily intake of each food and beverage group consumed per capita in the sub-population, and the counterfactual one to the distribution of the daily amount of each food and beverage group consumed by the individual during the survey.

### 2.3 Protein profile

Protein foods were defined as foods having more than 10% of their energy as protein and providing at least 5g/d of protein in high consumers (i.e. at the 90th percentile of the INCA3) (De Gavelle et al., 2018). Protein foods were then classified into twelve groups of protein sources (refined grains, whole grains, dairy, eggs, ruminant meat, poultry, pork, processed meat, fish, fruits – vegetables, pulses, and others) exhibiting differences in nutritional properties and the way they are eaten. Each group’s daily protein intake was then calculated for each individual.

### 2.4 Agribalyse database and environmental database

The Agribalyse database was used to assess environmental indicators of the diet. This database was developed by the *Agence de la transition écologique* (ADEME) using life cycle analyses (LCA) (ADEME, 2020a). Agribalyse is a database reporting 14 environmental indicators (Supplemental Method 1): greenhouse gas emissions in carbon dioxide equivalent (kg CO2 eq); exposure ionizing radiation in equivalent of kilobecquerels of Uranium 235 (kg U235 eq); photochemical ozone (O3) formation in equivalent of kilograms of non-methane volatile organic compounds (kg NMVOC eq); ozone depletion in equivalent of kilograms of trichlorofluromethane (Freon-11); emission of particulate matter in change in mortality due to particulate matter emissions; acidification in equivalent of moles hydron (mol H+ eq); terrestrial eutrophication in equivalent of moles of nitrogen (mol N eq); freshwater eutrophication in equivalent of kilograms of phosphorus (kg P eq); marine eutrophication in equivalent of kilograms of nitrogen (kg N eq); freshwater ecotoxicity in Comparative Toxic Unit for ecosystems (CTUe) an indicator based on a model called USEtox; water use in cubic meters of water; land use in loss of soil organic matter content in kilograms of carbon deficit (kg C deficit); fossils resource use in MJ; and metals and minerals resource use in equivalent of kilograms of antimony (kg Sb eq). Further description of the Agribalyse method is presented in Supplemental Method 1.

Here the Agribalyse database was used to assess the food eaten as listed in INCA3 (n= 1761) (Supplemental Figure 1). Two different methods were used to complete the 236 missing data: one specifically for GHGe and a second one for the other environmental impacts in Agribalyse (the method is described in detail in Supplemental Figure 2 and Supplemental Figure 3).

Data from the SHARP database were used to complete missing values for the GHGe indicator (Mertens et al., 2019). This database also includes data on land use but because the unit is different, we were unable to impute the data. Supplemental method 2 explains the process of merging the Agribalyse and SHARP database to the foods eaten in the population.

### 2.5 Statistical analysis

A Non-negative Matrix Factorization (NMF) was performed on 11 proteins groups. The protein group composed of the other type of protein food was not included. NMF is a method for identifying independent factors adapted for non-negative data with excess zeros (Lee and Seung, 1999) and has already been used by our team to identify dietary profiles in another survey (De Gavelle et al., 2018). Specifically, the Ls-nmf method (Wang et al., 2006) was used, and decomposition in six factors was conserved since it had the best R^2^ value. Then hierarchical ascendant classification on the factors was performed to identify protein profiles in the population. The number of clusters chosen was based on both the elbow and silhouette methods (Kaufman and Rousseeuw, 2005). The consumption of the 12 proteins groups between all profiles was compared using an ANOVA. The environmental indicators and diet quality scores of the protein profiles were compared using an ANCOVA adjusted for energy intake. Statistical analyses were performed using SAS software (version 9.4, SAS Institute Inc, Cary, NC, USA) and R version 4.0.3.

## 3 Results

### 3.1 Food group consumption and environmental and nutritional indicators

#### 3.1.1 Consumption

The hierarchical ascendant classification identified five distinct profiles (figure 1). The characteristics of the population and the different profiles are described in Table 1. Profile 1 (Low meat profile) represents consumers with a low protein intake from meat (9 g/day vs. 20.8 g/day in the total population), who also have a relatively higher intake of protein from dairy products (13.9 g/day vs. 9.8 g/day in the total population). Supplemental Table 5 shows that the higher dairy protein intake mostly comes from the higher consumption of milk (88 g/day vs. 80 g/day in the total population) and cheese (54 g/day vs. 42 g/day in the total population).

**Figure 1.**
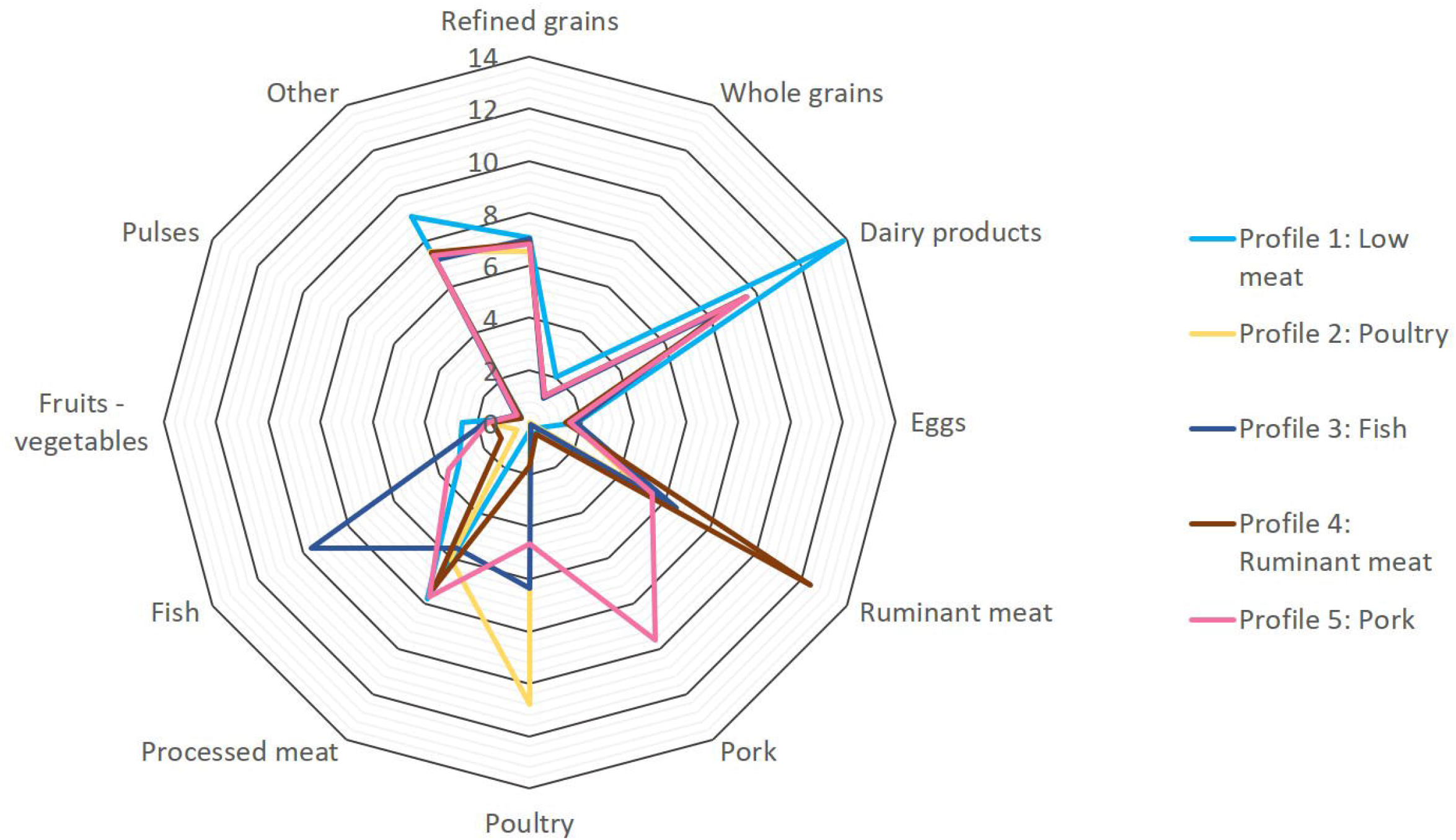
Consumption (g/day) of protein groups in individuals in the 5 protein profile as identified in the dietary survey (INCA3).

**Table 1.**
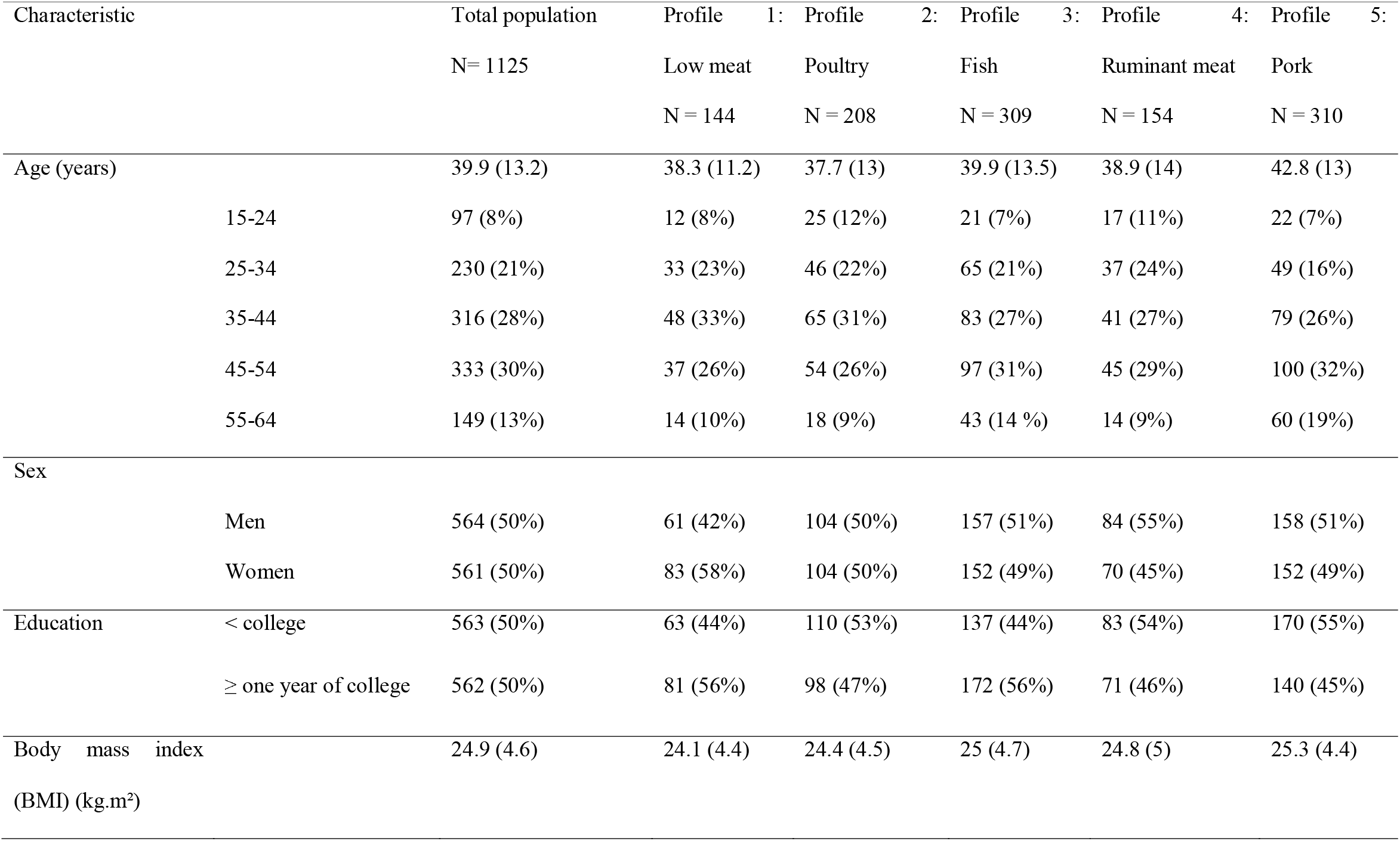
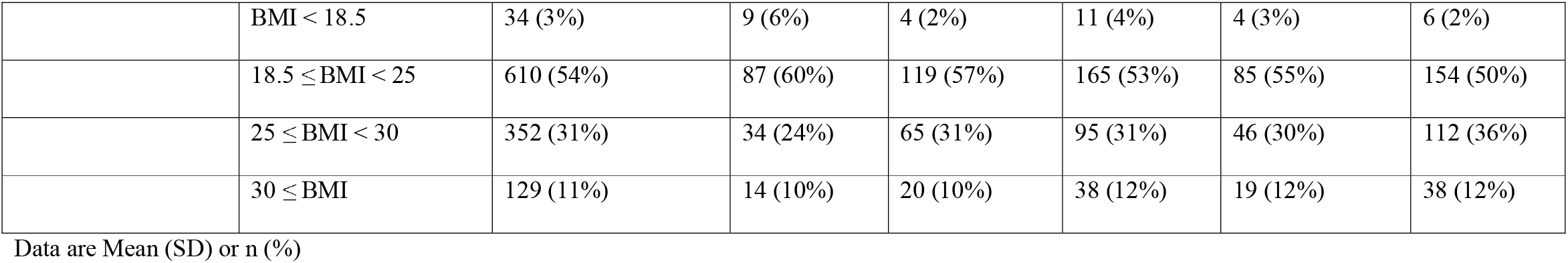
Characteristics of the final sample and the five protein profiles (INCA3, 2015-2016, n=1125)

Profile 2 (Poultry profile) represents people with high intakes of protein from poultry (10.8 g/day vs. 5.3 g/day in the total population), while profile 3 (Fish profile) represents individuals with the highest fish protein intake (9.6 g/day vs. 4.4 g/day in the total population). Profile 4 (Ruminant meat profile) represents individuals whose protein intake is largely contributed by ruminant meat proteins (12.4 g/day vs. 6.1 g/day in the total population), and profile 5 (Pork profile) represents people with high intakes of protein from pork (9.6 g/day vs. 2.7 g/day in the total population).

The consumption of 33 food groups in grams per day is also presented in Supplemental Table 5.

#### 3.1.2 Environmental indicators

Figure 2 presents the environmental indicators for each protein profile (see Supplemental Table 6). The profile with high intake of protein from ruminant has the highest environmental pressure for most of these indicators. In fact, this profile has the highest values for 5 of the 14 indicators. Notably, this profile also has the highest GHGe (7.7 kg CO2 eq/day vs. 6.4 kg CO2 eq/day in the total population). Compared to the total population, the land use, the emission of particulate matter, the acidification, and the terrestrial eutrophication are also at their highest for this profile, being 26%, 14%, 16%, and 19% higher, respectively.

**Figure 2.**
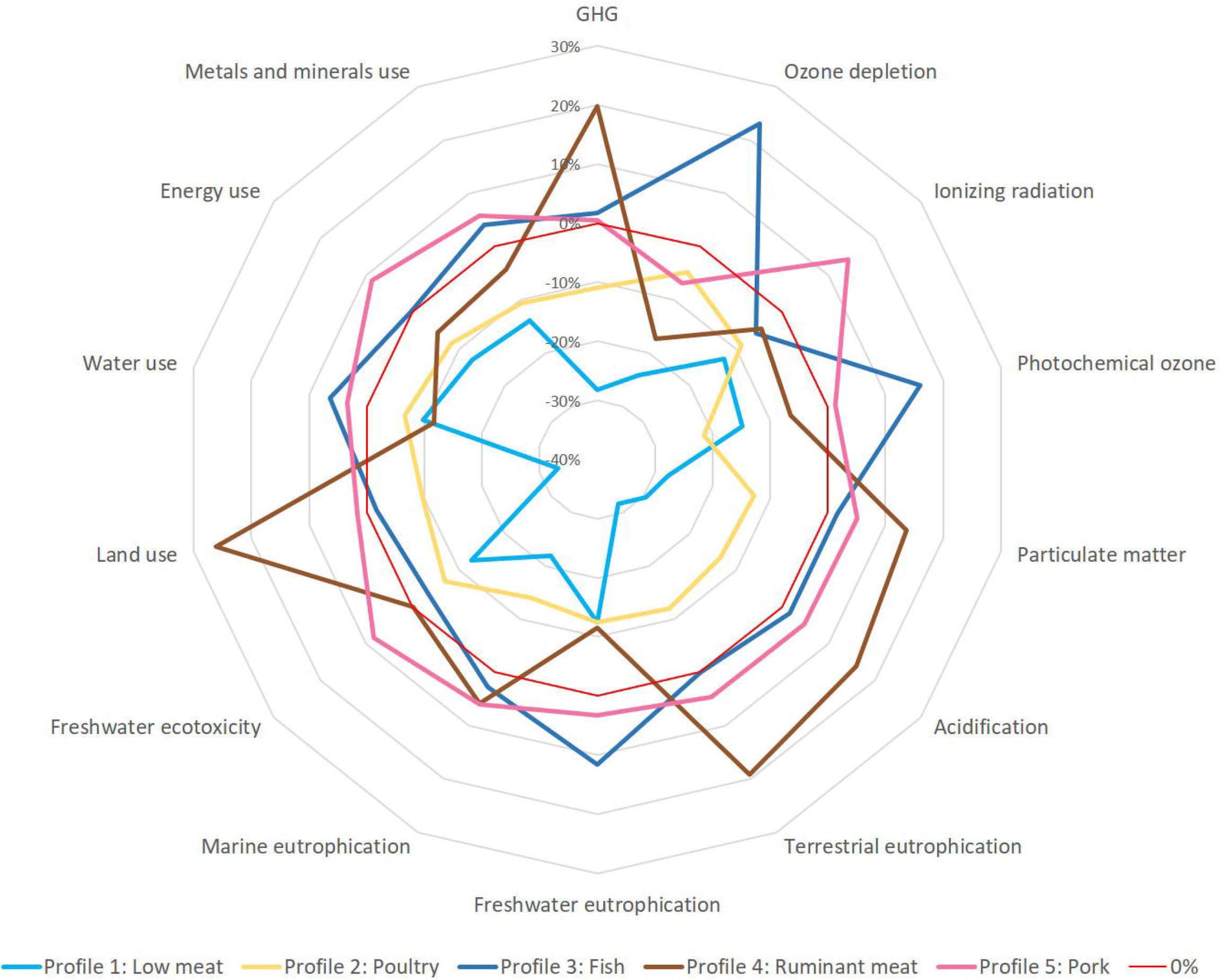
Environmental indicators associated with the diet consumed by individuals in the 5 protein profiles as identified in the dietary survey (INCA3). Data are in percentage of difference to the mean value of the total population. GHGe is the emission of greenhouse gases measured in kg CO_2_ eq. The ozone depletion is in kg CFC-11eq. The photochemical ozone formation is in kg of Non-Methane Volatile Organic Compounds eq. Particulate matter is in kg of PM_2.5_ emitted. The acidification is in mol H^+^ eq. The terrestrial eutrophication is in mol N eq, the freshwater eutrophication is in kg P eq, and the marine eutrophication is in kg N eq. The freshwater ecotoxicity is based on the USEtox model. Land use is in kg C deficit, water use in m^3^, fossils resource use in MJ, and Metals and minerals use in kg SB eq.

The profile with high intake of protein from fish and the one with the highest pork protein intake have the next most elevated GHGe (6.6 kg CO2 eq/day and 6.4 kg CO2 eq/day). For the other indicators, these two profiles are different. The profile with high intake of protein from pork contributes the most to freshwater ecotoxicity (164 CTUe/day vs. 151 CTUe/day in the total population), marine eutrophication (26 kg N eq/day vs. 25 kg N eq/day in the total population), ionizing radiation (1.7 kg U235eq/day vs. 1.5 kg U235eq/day in the total population), energy use (67 MJ/day vs. 62 MJ/day in the total population), and metals and minerals use (10.3 kg Sb eq/day vs. 9.7 kg Sb eq/day in the total population). For profile with high intake of protein from fish, it contributes the most to ozone depletion (0.73 Freon-11/day vs. 0.59 Freon-11/day in the total population), photochemical ozone (20 kg NMVOC/day vs. 17 kg NMVOC/day in the total population), freshwater eutrophication (1.15kg P eq/day vs. 1.03 kg P eq/day in the total population) and water use (7.1m3/day of water vs. 6.7 1m3/day of water in the total population).

The profiles with the lowest environmental impacts are the poultry profile and the low meat profile. These profiles emit the smallest amount of GHGe with 5.6 kg CO2 eq/day for the high consumers of poultry protein and 4.6 for the low meat consumers. This last profile has the lowest impact for most environmental indicators (11 from the 14 indicators) and is the second-best profile for the three others.

Supplemental Figures 3 to 16 show the variability of the environmental indicators for each food group, and Supplemental Table 7 describes each food group’s contribution to every environmental indicator for each profile.

#### 3.1.3 Nutritional quality

Figure 3 shows the nutritional and health scores of the different profiles. The profile with high fish consumers has the highest scores for the AHEI, the LAMD, and the second-best score for the PNNSGS2, the SecDiet, the PANDiet, and the HiDiet. The profile with the lowest meat protein consumption has the highest HiDiet score, and the one with the highest ruminant meat protein intake has the lowest HiDiet score (respectively +3% and -4% compared to the total population). The profile with the lowest meat protein consumption also has the lowest PANDiet (−3%) and a neutral SecDiet (0%) score. The value of the health scores are presented in Supplemental Table 8.

**Figure 3.**
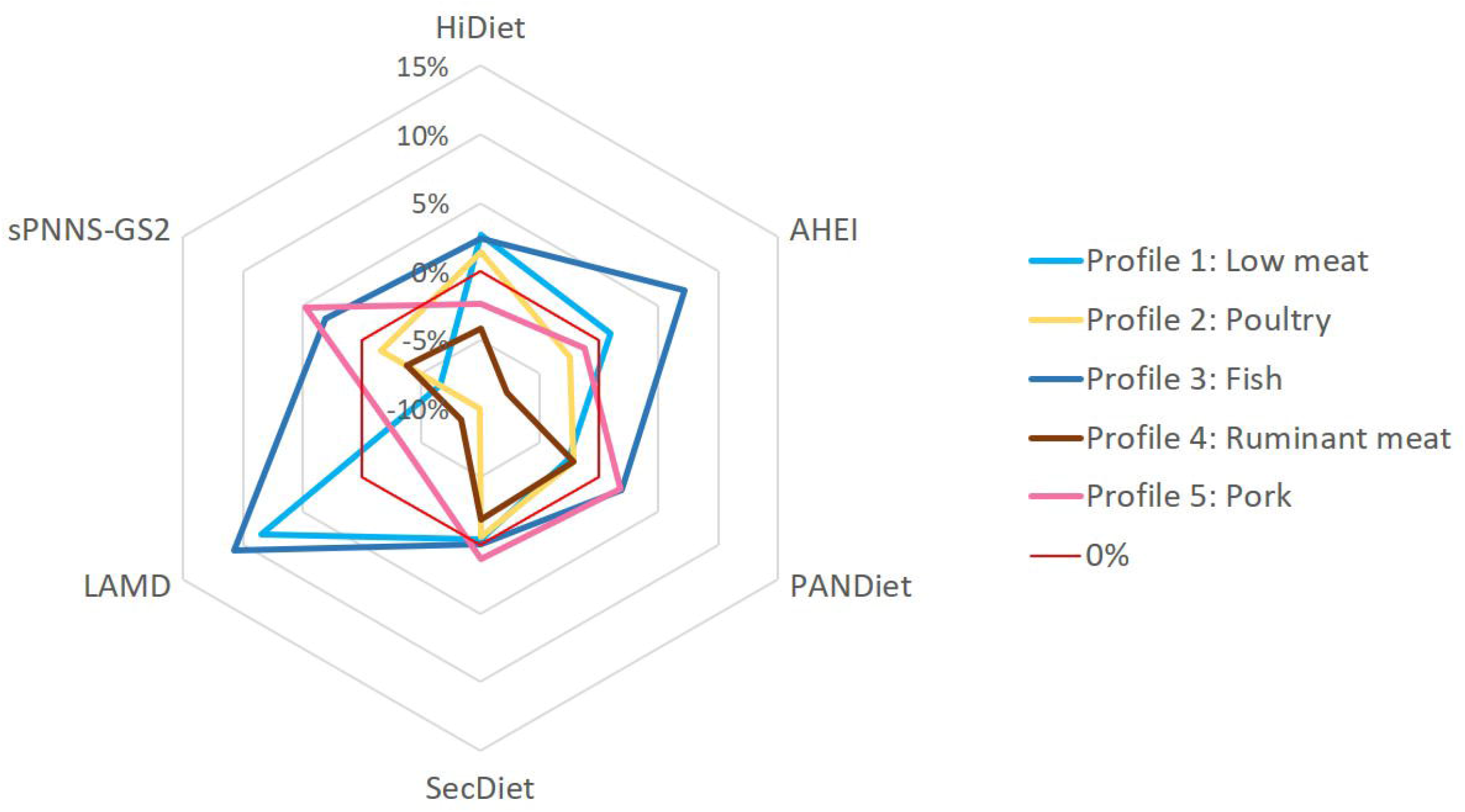
Nutritional and health indicators by profile. All data are in percentage of difference to the mean value of the population. The HiDiet was used to assess the diet impact on long-term mortality and morbidity (variation between -1 and 1). The Alternative Healthy Eating Index (AHEI-2010) is a modified Healthy Eating Index, assessing the adherence to *Dietary Guidelines for Americans*, improving target food choices and macronutrient sources associated with reduced chronic disease risk (maxpoint = 100). The PANDiet evaluates the probability of adequate nutrient intake (maxpoint = 100). The SecDiet evaluates the nutrient risk of overt deficiency (maxpoint = 1). The Literature-Based Adherence Score to the Mediterranean Diet (LAMD) assesses adherence to the Mediterranean diet (maxpoint = 16). sPNNS-GS2 the adherence to the French Food-based Dietary Guidelines (maxpoint = 10.5).

## 4 Discussion

In this study, we have identified five distinct profiles based on protein consumption, which show the variety of protein consumption patterns. These patterns are easy to interpret, and display contrasting characteristics on both environmental and nutritional/health dimensions.

### 4.1 Environmental differences between profiles

The high ruminant meat protein consumption profile was one of the worst regarding health scores and environmental indicators. This is in line with the modeled diets and dietary guidelines featuring low red meat intake (Kesse-Guyot et al., 2020; Willett et al., 2019). Although ruminant meat is well known for its high environmental pressures from GHGe and land use (Clark et al., 2018; Poore and Nemecek, 2018), our study has shown that actual protein intake from meat is also associated with other environmental pressures. The particulate matter (PM) production is significantly higher in the diet of people with high intake of protein from ruminant meat, with a high production of PM coming from manure (Garcia et al., 2013). It could be argued that this is not an important issue given the small percentage of particulate matter coming from agriculture compared to other sources like transport (Cambra-López et al., 2010). However, this percentage is growing, with PM emissions from agriculture being estimated at about 25% of total emissions in the Netherlands (Cambra-López et al., 2010). The Global Burden of Disease estimated that air pollution is responsible for about 1.2% of premature deaths and 0.5% of lost life years (Cohen et al., 2005). It is considered as the ninth larger risk factor contributing to deaths in the United States (The US Burden of Disease Collaborators, 2018). Acidification causing soil degradation (Goulding, 2016) and acid rain are other environmental impact of animal production. The dominant source of acidification in agriculture comes from ammonia emissions released used for feed production. With a higher feed conversion ratio, ruminants thus have a higher acidification power per meat produced (Röös et al., 2013).

The low meat profile had the smallest environmental pressure. This is in line with the literature as most publications pointed out that meat, especially from ruminant and to a lesser extent from monogastric, has by far the most significant environmental impact, specifically for GHGe (Poore and Nemecek, 2018). Indeed, low GHGe in this profile should be ascribed to the very low consumption of ruminant meat. This is similar with other studies showing plant-based diets having lower GHEe compare to omnivorous diet (Rabès et al., 2020). Note that dairy intake was relatively high for this profile – in fact the highest of the 5 profiles – which may be considered a practical conflict between the production of ruminant meat and the production of milk. This is because of the co-production factor; if the bovine dairy protein intake is above 0.43 times the bovine meat protein intake, there is an excess meat production (Barré et al., 2018). In the case of the low meat consumer profile, the bovine dairy protein consumption exceeded this number. However, this issue only occurred in the hypothetical situation where the entire population follows this diet, with no meat exportation or dairy importation.

It is also important to note that a high share of red meat consumed comes from dairy cows in France (Assmann, 2020). Thus, the allocation of the environmental pressure using biophysics systems in the life cycle inventory (LCI) is considered. The literature showed different types of allocations underlying different choices. These choices came from 4 criteria: compliance with recommendations, stability in time and space, and practical aspects (Wilfart et al., 2021). However, on a food basis, the indicators already studied, like GHGe, have a similar order of magnitude of allocations as other studies (Clune et al., 2017), indicating the choices made in Agribalyse (ADEME, 2020b) are similar to that of most published studies.

This low-meat protein profile also has drawbacks on a nutritional aspect, with the lowest PANDiet values of all profiles. This is mainly explained by the low intake of nutrient-dense food sources, in particular red meat, which is not offset by higher intakes of fruits and vegetables. It should be noted that even if it is the most plant-based profile, the consumption of pulses, whole grains and other recommended plant food groups is far from what is recommended in optimization studies (Dussiot et al., 2022; Willett et al., 2019). Moreover, the nutrient security in this low-meat profile was not lower than that of the general population, confirming that even if there are some slightly lower intakes of some nutrients, there is no overall increase in the risk of overt deficiency. As far as long-term health is concerned, this profile has the lowest estimated risk of long-term morbidity or mortality from coronary heart disease, stroke, type 2 diabetes, and colorectal cancer, as assessed using the HiDiet score. Given that the small increase in nutrient inadequacy does not increase the risk of overt deficiency, this profile shows strong co-benefits for environmental and human health.

Compared to the other profiles high in animal flesh (meat or fish), the profile with high intakes of protein from poultry has a lower impact on most of the environmental indicators. In fact, GHGe are specifically one of the advantages of this profile. Similar results from previous studies show that poultry produces the lowest GHGe among all types of meat (González et al., 2011; Nijdam et al., 2012).

Surprisingly, this profile exhibited poor dietary scores with values under the population average for all scores except the HiDiet. This latter score was the only one in line with the epidemiological data showing that poultry consumption was not associated with a higher risk of mortality or morbidity (Du et al., 2020; Makiuchi et al., 2020; Micha et al., 2017a, 2017b). The low scores for the other nutritional and health scores may be explained by the low fish consumption and the fact that other food categories with a positive weight in these health and nutritional scores are low in the poultry profile.

The profile with a high fish consumption has high values for all health and nutritional scores. This was expected as fish consumption is positively weighted in all of the scores considered in this study. Fish is a very good contributor to bioavailable nutrients and has well-known health benefits against chronic disease (Bogard et al., 2019). The high intake of indispensable nutrients associated with the higher level of protein for fish but also an overall dietary profile of higher quality explains the high PANDiet and SecDiet score. In contrast, this high fish intake has some important drawbacks on the environmental indicators. As described before, water use and photochemical ozone formation are two important issues of fish consumption (Ruiz-Salmón et al., 2021). It is important to note that there is significant variability between fish species, and it may be possible to improve the health and environmental impact by consuming different fish species (ADEME, 2020a). However, these results need to be verified with other LCA as environmental pressures from fish are not precise, being mostly measured indirectly.

### 4.2 Limits and strengths

First, it should be noted that INCA3 survey was conducted between 2014 and 2015, so diets may have changed since then, and the sample of 1125 individuals that we used is small. However, it is the latest French representative survey to date. The environmental database Agribalyse has also some limitations. For example, the quantification of soil carbon storage and removal is not considered in the calculation of the GHGe. Biodiversity and the impact of phytosanitary products are also not yet calculated. Another drawback is that there is no information about farming practices (such as organic production) for foods production that are consumed in the survey, and the environmental indicators are only for mean of consumed foods differing in farming practices. However, this database includes a large variety of environmental indicators covering the entire food chain from production to the plate and virtually all the foods consumed by the French population since it was created for this purpose.

Finally, the major strength of this study is the variety of indicators and food described in a representative survey. Using this information, it was possible to precisely describe 14 environmental indicators and 6 nutritional and health dietary scores for five protein profiles in a representative sample of the French population

## 5 Conclusion

In the present study, we showed that the profiles of protein intake of the population are varied and have contrasting associations with health and environmental impacts. The protein profile marked by ruminant meat had the worst scores on both health and environmental aspects. Conversely, the most environment-friendly protein profile is very low in meat, and this profile also had the lowest risk for long-term morbidity and mortality. As we studied real profiles identified in the general population, the differences in health and environmental impacts between profiles may be useful to consider realistic targets for acceptable changes in the diet. These changes may be more practical than those identified by modeling studies. Our results thus support the importance of protein profiles for health and environmental impacts.

## Data Availability

All data produced are available online at:
https://www.data.gouv.fr/fr/datasets/donnees-de-consommations-et-habitudes-alimentaires-de-letude-inca-3/
https://agribalyse.ademe.fr/
https://easy.dans.knaw.nl/ui/datasets/id/easy-dataset:132158

https://agribalyse.ademe.fr/

https://www.data.gouv.fr/fr/datasets/donnees-de-consommations-et-habitudes-alimentaires-de-letude-inca-3/

## Acknowledgments and statement of authors’ contributions to manuscript

EP, FM, and EK-G: designed the research; EP: conducted the research; EP, FM, and EK-G analyzed the data; MS, and J-FH provided methodological support; EP drafted the first version of the manuscript, EP, FM, and EK-G wrote the manuscript and all authors provided critical comments on the manuscript. EP, FM, and EK-G had primary responsibility for the final content, and all authors read and approved the final manuscript.

## Notes

**Financial support:** This research received no external funding.

**Conflict of Interest** None.

### Competing Interest Statement

The authors have declared no competing interest.

### Funding Statement

This study did not receive any funding

### Author Declarations

The study used only openly available human data that were originally located at: https://www.data.gouv.fr/fr/datasets/donnees-de-consommations-et-habitudes-alimentaires-de-letude-inca-3/ https://agribalyse.ademe.fr/ https://easy.dans.knaw.nl/ui/datasets/id/easy-dataset:132158

