## Supplement data for "Dietary protein consumption profiles show contrasting impacts on environmental and health indicators"

SUPPLEMENTAL MATERIAL

Supplemental Table 1. Scoring for Each Component of the Literature-based Adherence Score to the Mediterranean Diet

| **Component** | **Attribution of points** | | |
| --- | --- | --- | --- |
| **To consume in large amounts** | 0 | 1 | 2 |
| Fruits | < 150g/d | 150-225g/d | >225 g/d |
| Vegetables | < 100g/d | 100-250g/d | >250 g/d |
| Legumes | < 10g/d | 10-20g/d | >20 g/d |
| Cereals | < 130g/d | 130-195g/d | >195 g/d |
| Fish | < 14.29g/d | 14.29-35.71g/d | >35.71g/d |
| **To consume in moderation** |  |  |  |
| Alcohol | >24 g/d | < 12g/d | 12-24g/d |
| **To consume in limited amounts** |  |  |  |
| Meat (including poultry) | >120 g/d | 80-120g/d | < 80g/d |
| Dairy | >270 g/d | 180-270g/d | < 180g/d |

Supplemental Table 2 The AHEI-2010 scoring method

| **Component** | **Serving size (g)** | **Criteria for minimum score (0)** | **Criteria for maximum score (10)** |
| --- | --- | --- | --- |
| Vegetables, *servings/day* | 118.3 | 0 | **≥**5 |
| Fruits, *servings/day* | 118.3 | 0 | **≥4** |
| Whole grains, *g/day* |  |  |  |
| Women |  | 0 | **75** |
| Men |  | 0 | **95** |
| Sugar-sweetened beverages and fruit juice, *servings/day* | 226.8 | **≥1** | 0 |
| Nuts and legumes, *servings/day* | 28.4 | 0 | **≥1** |
| Red/processed meat, *servings/day* | Red meat 113.4 Processed meat 42.5 | **≥1.5** | 0 |
| Long-chain (n-3) fats (EPA + DHA), *mg/day* |  | 0 | 250 |
| PUFA, *% of energy* |  | ≤2 | **≥10** |
| Sodium, *mg/day* |  | Highest decile | Lowest decile |
| Alcohol, *drinks/day* | 10 grams of ethanol |  |  |
| Women |  | **≥2.5** | 0.5-1.5 |
| Men |  | **≥3.5** | 0.5-2.0 |
| **Total** |  | 0 | 100 |

Supplemental table 3 Components, scoring and weighting used for sPNNS-GS2 computation

| **Dietary components** | **Recommendation** | **Criteria** | **Score** |
| --- | --- | --- | --- |
| Fruit and | At least 5 serv/d, | [0 - 3.5[ | 0 |
| vegetables | with 1 max as juice and 1 | [3.5 - 5[ | 0.5 |
| (weight=3) | max as dried | [5 - 7.5[ | 1 |
|  |  | ≥7.5 | 2 |
| Nuts | A handful/d | 0 | 0 |
| (weight=1) |  | ]0 – 0.5[ | 0.5 |
|  |  | [0.5- 1.5[ | 1 |
|  |  | ≥1.5 | 0 |
| Legumes | At least 2 serv/w | 0 /w | 0 |
| (weight=1) |  | ]0-2[ /w | 0.5 |
|  |  | ≥2 /w | 1 |
| Whole-grain | Every day | 0 | 0 |
| Food |  | ]0 - 1[ | 0.5 |
| (weight=2) |  | [1 - 2[ | 1 |
|  |  | ≥2 | 1.5 |
| Milk and | 2 serv/d | [0 - 0.5[ | 0 |
| dairy products |  | [0.5 - 1.5[ | 0.5 |
| (weight=1) |  | [1.5 - 2.5[ | 1 |
|  |  | ≥2.5 | 0 |
| Red meat | Limit consumption | ≥750 g/w | -2 |
| (weight=2) |  | [500 - 750[ g/w | -1 |
|  |  | [0 - 500[ g/w | 0 |
| Processed meat | Limit consumption | ≥300 g/w | -2 |
| (weight=3) |  | [150 - 300[ g/w | -1 |
|  |  | [0 - 150[ g/w | 0 |
| Fish and | 2 serv/w | [0 - 1.5[serv /w | 0 |
| Seafood |  | [1.5 - 2.5[serv /w | 1 |
| (weight=2) |  | [2.5 - 3.5[serv /w | 0.5 |
|  |  | ≥3.5 serv /w | 0 |
| Added fat | Avoid overeating | >16% of EIWA ^c^ | 0 |
| (weight=2) |  | ≤16% of EIWA | 1.5 |
| Sugary foods | Limit consumption | ≥15% of EIWA | -2 |
| (weight=3) |  | [10-15[% of EIWA | -1 |
|  |  | <10 % of EIWA | 0 |
| Sweet-tasting | Limit consumption | ≥ 750mL mL/d | -2 |
| beverages |  | [250 - 750[ mL/d | -1 |
| (weight=3) |  | ]0 - 250[ mL/d | -0.5 |
|  |  | 0 mL/d | 0 |
| Alcoholic | Limit consumption | >200 g/d | -2 |
| beverages |  | ]100-200] g/d | -1 |
| (weight=3) |  | ]0-100] g/d | 0 |
|  |  | 0 g/d | 0.5 |
| Salt | Limit consumption | >12 g/d | -2 |
| (weight=3) |  | ]10-12] g/d | -1 |
|  |  | ]8-10] g/d | -0.5 |
|  |  | ]6-8] g/d | 0 |
|  |  | ≤6 g/d | 1 |

Abbreviations: EIWA: Energy intake without alcohol, w: week

Supplemental table 4 Protein intake (g/d and % energy intake without alcohol) from protein food groups in the total population and across the five protein profiles.

|  |  | Overall | Profile 1: Low meat | Profile 2: Poultry | Profile 3: Fish | Profile 4: Ruminant meat | Profile 5: Pork |
| --- | --- | --- | --- | --- | --- | --- | --- |
| **Refined grains** | g/day | 6.9 (3.5) | 7.1 (4) | 6.5 (3.3) | 7 (3.5) | 6.9 (4.2) | 6.8 (2.9) |
|  | % EI | 1.3% (0.6) | 1.3% (0.7) | 1.2% (0.6) | 1.3% (0.7) | 1.3% (0.8) | 1.2% (0.5) |
| **Whole grains** | g/day | 1.2 (2.6) | 2 (3.3) | 1.2 (2.7) | 1.1 (2.5) | 1.1 (2.6) | 1.2 (2.4) |
|  | % EI | 0.2% (0.5) | 0.4% (0.6) | 0.2% (0.5) | 0.2% (0.5) | 0.2% (0.5) | 0.2% (0.4) |
| **Dairy products** | g/day | 9.8 (7.1) | 13.9 (10.9) | 9.4 (6.2) | 8.7 (6.1) | 9.6 (6.5) | 9.6 (6.1) |
|  | % EI | 1.8% (1.2) | 2.5% (1.6) | 1.7% (1) | 1.6% (1.1) | 1.7% (1.2) | 1.7% (1.1) |
| **Eggs** | g/day | 1.6 (2.6) | 1.8 (2.5) | 1.5 (2.6) | 1.8 (2.7) | 1.4 (2.7) | 1.6 (2.5) |
|  | % EI | 0.3% (0.5) | 0.4% (0.5) | 0.3% (0.5) | 0.3% (0.5) | 0.2% (0.5) | 0.3% (0.5) |
| **Ruminant meat** | g/day | 6.1 (5.9) | 0.4 (2) | 5 (5.4) | 6.5 (4.6) | 12.4 (6.1) | 5.4 (5) |
|  | % EI | 1.2% (1.3) | 0.1% (0.3) | 0.9% (1) | 1.3% (1) | 2.5% (1.5) | 1% (1) |
| **Pork** | g/day | 2.7 (5) | 0.5 (2.7) | 0 (0.1) | 0.1 (1.1) | 0.5 (2.5) | 9.6 (2.9) |
|  | % EI | 0.5% (0.9) | 0.1% (0.4) | 0% (0) | 0% (0.2) | 0.1% (0.4) | 1.8% (0.8) |
| **Poultry** | g/day | 5.3 (6.2) | 0.3 (1.8) | 10.8 (4.6) | 6.4 (5.9) | 1.7 (4.6) | 4.6 (5.7) |
|  | % EI | 1% (1.2) | 0% (0.3) | 2% (0.9) | 1.2% (1.2) | 0.3% (0.9) | 0.8% (1.1) |
| **Processed meat** | g/day | 6.7 (6) | 7.8 (7.5) | 6 (5.3) | 5.6 (4.9) | 7.3 (6.4) | 7.7 (6.2) |
|  | % EI | 1.2% (1.2) | 1.5% (1.5) | 1.1% (0.9) | 1.1% (1.1) | 1.4% (1.3) | 1.3% (1.1) |
| **Fish** | g/day | 4.4 (5.8) | 3.1 (4.2) | 0.6 (2.1) | 9.6 (5.2) | 1.2 (3.4) | 3.6 (5.2) |
|  | % EI | 0.8% (1.2) | 0.7% (1) | 0.1% (0.5) | 1.9% (1.2) | 0.2% (0.6) | 0.6% (1) |
| **Fruits - vegetables** | g/day | 1.7 (2.3) | 2.6 (3.3) | 1.6 (2.3) | 1.7 (2.1) | 1.4 (1.8) | 1.6 (1.9) |
|  | % EI | 0.3% (0.5) | 0.5% (0.6) | 0.3% (0.6) | 0.3% (0.4) | 0.3% (0.3) | 0.3% (0.4) |
| **Pulses** | g/day | 0.5 (1.3) | 0.4 (1.1) | 0.5 (1.2) | 0.6 (1.4) | 0.4 (1.1) | 0.5 (1.3) |
|  | % EI | 0.1% (0.2) | 0.1% (0.2) | 0.1% (0.2) | 0.1% (0.2) | 0.1% (0.2) | 0.1% (0.2) |
| **Other** | g/day | 7.6 (5.8) | 9.1 (5.9) | 7.6 (5.6) | 7.2 (5.6) | 7.5 (6.6) | 7.4 (5.4) |
|  | % EI | 1.5% (1.3) | 1.8% (1.3) | 1.5% (1.3) | 1.4% (1.3) | 1.5% (1.4) | 1.3% (1.1) |

Supplemental table 5 Consumption by food group for the total population and by profile. Profile 1: Low meat, Profile 2: Poultry, Profile 3: Fish, Profile 4: Ruminant meat, Profile 5: Pork. All data are in g/day.

|  | **No adjustment** | | | | | | **Adjustment for total energy intake without alcohol** | | | | | |
| --- | --- | --- | --- | --- | --- | --- | --- | --- | --- | --- | --- | --- |
|  | **Overall** | **Profile 1** | **Profile 2** | **Profile 3** | **Profile 4** | **Profile 5** | **Overall** | **Profile 1** | **Profile 2** | **Profile 3** | **Profile 4** | **Profile 5** |
| **Vegetable** | 168.8 (129) | 173.7 (142.1) | 161.4 (117.4) | 177.6 (129.3) | 140.1 (122) | 178.6 (130.8) | 181.3 (153.6) | 196 (183.7) | 172.7 (138.1) | 191.6 (160.9) | 151.3 (123.4) | 186.4 (152.3) |
| **Fruits** | 134.1 (151.5) | 116.9 (135.3) | 124.6 (145.4) | 140 (170.5) | 121.5 (123.6) | 149.1 (154) | 139.3 (154.3) | 117.9 (126.4) | 135.8 (154.2) | 140.2 (156.7) | 136.5 (159) | 151.7 (160.9) |
| **Nuts. seeds and oleaginous fruits** | 2.7 (8.8) | 5.8 (13.6) | 2.8 (10.5) | 2.5 (7.4) | 1.9 (7.6) | 1.9 (5.9) | 2.7 (8.8) | 6.3 (15.4) | 2.6 (8.9) | 2.5 (7.4) | 1.7 (6.4) | 2 (6) |
| **Refined bread and bread products** | 144.4 (110.4) | 131.2 (99) | 157.8 (117.8) | 132.9 (102.6) | 138.9 (115.7) | 156.3 (113.1) | 139.1 (87.7) | 132.1 (87.2) | 154.1 (97.2) | 131.5 (87.3) | 133.5 (89) | 143.3 (79.5) |
| **Whole and semi-whole bread and bread products** | 12.4 (33) | 15.2 (32.4) | 10.4 (28.5) | 12.8 (34.9) | 13.2 (35.1) | 11.7 (33.3) | 12.6 (31.6) | 15.4 (33.8) | 10.6 (28.2) | 13.2 (33.9) | 13.5 (31.2) | 11.6 (30.7) |
| **Other refined starchy foods** | 152.6 (124) | 130.6 (111.3) | 134.2 (97.4) | 156.3 (119) | 167.2 (133.8) | 162.8 (141.9) | 154 (116.9) | 139.9 (123.6) | 140.4 (105.5) | 163 (128) | 169.6 (117.4) | 151.2 (107.6) |
| **Other whole and semi-wholemeal starchy foods** | 4.2 (17.6) | 10.4 (28) | 4.7 (19.3) | 3.1 (13.3) | 1.4 (8.9) | 3.8 (16.6) | 4.6 (20.6) | 13.2 (39.5) | 4.6 (18.8) | 3.2 (13.7) | 1.6 (10.1) | 4 (17.5) |
| **Starch products. processed** | 24.1 (37.1) | 28.9 (38.6) | 22.8 (37.1) | 23.3 (38.4) | 32.7 (45.9) | 18.7 (28.4) | 23.6 (34) | 29.9 (40) | 23.2 (34.5) | 21.8 (32.6) | 29.9 (37.9) | 19.4 (28.7) |
| **Legumes** | 10.1 (31.8) | 5.5 (20.4) | 10.8 (29.5) | 15.2 (43.2) | 4.4 (19.3) | 9.5 (28) | 9.1 (26.4) | 5.9 (24) | 10 (27.4) | 12.8 (32) | 4.3 (15.9) | 8.8 (24.4) |
| **Poultry** | 30.9 (41.7) | 6.6 (17.7) | 62.9 (46) | 38.2 (41.8) | 8.9 (25.6) | 24 (34.4) | 32.9 (46.3) | 6.8 (17.6) | 68.6 (54) | 40.9 (46.2) | 9.5 (27.8) | 24.2 (37.1) |
| **Meat excluding poultry** | 63 (70.6) | 13 (23.3) | 38 (44.1) | 51.8 (65.1) | 92 (81.7) | 98.5 (72.3) | 63 (64.5) | 13.8 (24.1) | 39.9 (44) | 52.5 (59.6) | 94.4 (75.5) | 94.8 (60.9) |
| **Delicatessen** | 41.3 (46.5) | 46.4 (46.4) | 38 (43.5) | 32.8 (35.5) | 51.7 (55.5) | 44.5 (51.2) | 41.5 (45.3) | 50.4 (55.3) | 37.1 (38.6) | 33.5 (36.5) | 55.9 (60.8) | 40.9 (40.6) |
| **Fatty fish** | 7.4 (19.4) | 7.6 (20.4) | 2.4 (10.5) | 13.8 (25.5) | 2.5 (11.2) | 6.5 (17.6) | 8 (22.2) | 8.7 (23.8) | 3.6 (17.5) | 14.4 (27.7) | 2.5 (10.7) | 7.1 (20.3) |
| **Lean fish** | 23.7 (40.6) | 23 (36.7) | 5.5 (18.4) | 44.2 (47.8) | 9.7 (32) | 22.3 (38.4) | 24.3 (41.4) | 27 (44) | 7.2 (27.7) | 45.2 (47.1) | 9.6 (31.4) | 20.6 (35.1) |
| **Eggs and egg dishes** | 13.9 (24.3) | 18.5 (26.1) | 13.6 (23.3) | 14.3 (24.5) | 12.9 (26.9) | 12.1 (22.2) | 15.3 (28.7) | 21.5 (35.2) | 15 (27.6) | 15.7 (28.7) | 12.9 (27.6) | 13.6 (26.3) |
| **Milk** | 80.2 (141.2) | 78.2 (125.3) | 78.4 (128.5) | 93.1 (158.4) | 57.1 (115.2) | 81.4 (148.4) | 80.1 (138.6) | 88.1 (139.1) | 84.1 (144.1) | 86.5 (131.9) | 59.2 (129.6) | 78.8 (145) |
| **Fresh dairy products** | 81 (85.3) | 74.5 (74.6) | 75.9 (75.9) | 89.2 (93.3) | 65.8 (76.4) | 87.3 (90.4) | 85.9 (94.2) | 78.7 (88.1) | 84.7 (93.7) | 91 (88.4) | 72.7 (85.1) | 92.1 (106) |
| **Sweet dairy desserts** | 17.7 (34.6) | 16.7 (27.1) | 17.3 (35.8) | 17.7 (39.7) | 17.8 (36.2) | 18.5 (30.7) | 17.1 (32.6) | 17.6 (29) | 15.7 (29.1) | 17.5 (38.7) | 16.9 (33.1) | 17.5 (29.5) |
| **Cheese** | 43.2 (37.1) | 53.7 (45.6) | 38.8 (33.6) | 39.4 (31.9) | 48.9 (43.7) | 42.5 (35) | 42.3 (32.5) | 53.7 (38.7) | 37.8 (27.4) | 39.6 (31) | 47 (35.6) | 40.7 (30.9) |
| **Animal fats** | 10.5 (12.9) | 9.2 (11.7) | 12.8 (16.8) | 10 (11.4) | 9.7 (11.8) | 10.3 (12.1) | 10.3 (11.8) | 9.1 (10.5) | 12.3 (14.6) | 10 (10.9) | 9.2 (10.5) | 10.2 (11.5) |
| **Vegetable fats rich in ALA** | 0.3 (1.6) | 0.3 (1.1) | 0.1 (1) | 0.3 (1.8) | 0.4 (1.6) | 0.4 (2) | 0.4 (1.9) | 0.3 (1.1) | 0.1 (0.9) | 0.3 (1.9) | 0.5 (2) | 0.5 (2.5) |
| **Vegetable fats low in ALA** | 11.3 (11.3) | 10.5 (9.3) | 10.4 (9.4) | 10.2 (9.5) | 11.4 (14.8) | 13.4 (12.6) | 11.9 (12.3) | 11.6 (11.8) | 11.3 (10.7) | 10.9 (10.6) | 12.9 (19) | 12.9 (10.6) |
| **Sauces and fresh creams** | 33.4 (38) | 33.3 (39.3) | 35.3 (38.1) | 38 (36.3) | 35.4 (50.2) | 26.2 (30.3) | 35.3 (40.7) | 37 (45.9) | 36.8 (38.8) | 40.6 (42.6) | 37.4 (48.2) | 26.6 (30.9) |
| **Sweet products or sweet and fatty products** | 94.4 (78.1) | 101.3 (84.8) | 102.7 (86.9) | 94.5 (83.8) | 85 (70.3) | 90.9 (65.2) | 90.4 (62.7) | 99.6 (66.9) | 97.3 (66.1) | 88.9 (66.2) | 83.2 (58.1) | 87 (55.8) |
| **Drinking water** | 972.9 (671.9) | 936.8 (741.4) | 986.3 (703.7) | 931.2 (601.7) | 1020.9 (761.8) | 997.4 (634.6) | 1073.7 (970.1) | 1044.2 (911.9) | 1057.3 (852.9) | 1058.7 (941.8) | 1248.3 (1527.3) | 1016 (698.6) |
| **Sweet drinks and fruit juices** | 215.1 (339.1) | 220.7 (248.4) | 190 (226.7) | 287.4 (499.3) | 187 (293.7) | 168.2 (233.5) | 223.3 (336.9) | 248.3 (297) | 202.2 (238.8) | 281.9 (463.5) | 204.1 (318.7) | 174.7 (248.6) |
| **Hot drinks** | 499.4 (384.1) | 589.2 (469.8) | 449.2 (341.4) | 471.1 (386.5) | 465 (369.4) | 543.9 (360.7) | 536.9 (455.2) | 632.1 (482) | 473.8 (376.3) | 512.7 (512.6) | 539.3 (498.1) | 562.7 (396.9) |
| **Salt** | 1.4 (1.5) | 1.6 (1.8) | 1.4 (1.3) | 1.3 (1.3) | 1.3 (1.7) | 1.4 (1.7) | 1.5 (1.7) | 1.8 (2.2) | 1.6 (1.5) | 1.4 (1.6) | 1.4 (1.6) | 1.4 (1.8) |
| **Condiments** | 4.9 (10.7) | 5.3 (10.8) | 4.9 (10.4) | 4.6 (9) | 6.6 (16.1) | 3.9 (8.7) | 5.1 (10.9) | 5.4 (9.9) | 5.3 (10.8) | 4.7 (9.1) | 7.5 (17.9) | 3.8 (7.8) |
| **Soups and broths** | 77.3 (172) | 110.3 (236.5) | 82.6 (192) | 57.3 (130.3) | 75.3 (181.8) | 81.5 (150.7) | 81.9 (185.6) | 128.1 (278.6) | 88.1 (187) | 59.3 (134.6) | 73.3 (191.9) | 86.1 (167) |
| **Plant-based substitutes** | 3.5 (27.6) | 5.8 (28.2) | 3.3 (31.8) | 1.9 (16.3) | 3.4 (19.3) | 4.3 (35.6) | 3.9 (31) | 6.3 (30.3) | 3.2 (27.1) | 2.3 (21.4) | 3.4 (18.8) | 5.4 (44.1) |
| **Others foods** | 2.9 (7.2) | 2.7 (7.7) | 3.5 (8.4) | 2.4 (5.9) | 3.6 (8.9) | 2.5 (6) | 2.9 (7.5) | 2.9 (9) | 3.6 (9) | 2.6 (6.6) | 3.6 (8.2) | 2.5 (5.8) |
| **Alcoholic beverages** | 147 (306.8) | 92.9 (180.1) | 128.7 (234.8) | 125.1 (221.2) | 158.8 (296) | 200.9 (438.9) | 152 (364.2) | 108 (222.9) | 120.9 (221.1) | 124 (219.7) | 213.3 (607.8) | 188.8 (435) |

Values are in grams per day (SDTERR)

Supplemental table 6 Environmental indicators by profile adjusted for total energy intake.

|  | **Total population** | **Profile 1: Low meat** | **Profile 2: Poultry** | **Profile 3: Fish** | **Profile 4: Ruminant meat** | **Profile 5: Pork** |
| --- | --- | --- | --- | --- | --- | --- |
| **GHGe (kg CO2 eq)** | 6.4 (3.3) | 4,6 (1,9) | 5,6 (2,6) | 6,6 (3,4) | 7,7 (4,5) | 6,4 (2,9) |
| **Ionizing radiation (kg U235 eq)** | 1.5 (0.5) | 1,3 (0,2) | 1,3 (0,3) | 1,4 (0,4) | 1,4 (0,5) | 1,7 (0,4) |
| **Ozone depletion (kg CFC-11eq)** | 0.6 (1.1) | 0,4 (1,7) | 0,6 (2,7) | 0,7 (3,4) | 0,5 (4,4) | 0,6 (3) |
| **Photochemical ozone (kg of non-methane volatile organic compounds eq)** | 17.2 (9.6) | 14,6 (0,2) | 13,5 (1,2) | 20 (2) | 16,1 (0,4) | 17,4 (0,4) |
| **Particulate matter (kg of PM2.5emitted)** | 0.6 (0.3) | 0,4 (0,4) | 0,5 (0,5) | 0,6 (0,5) | 0,6 (0,6) | 0,6 (0,6) |
| **Acidification (mol H+ eq)** | 0.1 (0.1) | 0,1 (8,2) | 0,1 (5,8) | 0,1 (10,9) | 0,1 (9,4) | 0,1 (9,7) |
| **Terrestrial eutrophication (mol N eq)** | 0.3 (0.2) | 0,2 (0,2) | 0,3 (0,3) | 0,3 (0,3) | 0,4 (0,4) | 0,3 (0,3) |
| **Freshwater eutrophication (kg P eq)** | 1.0 (0.6) | 0,9 (0,1) | 0,9 (0,1) | 1,2 (0,1) | 0,9 (0,1) | 1,1 (0,1) |
| **Marine eutrophication (kg N eq)** | 24.5 (12.2) | 19,2 (0,1) | 21,1 (0,2) | 25,2 (0,2) | 25,9 (0,3) | 26 (0,2) |
| **Freshwater ecotoxicity (USEtox model)** | 151.4 (58.4) | 132,2 (0,5) | 140,8 (0,4) | 146,2 (0,8) | 151,2 (0,5) | 164,2 (0,5) |
| **Land use (kg C deficit)** | 314.4 (191.8) | 210,3 (7,4) | 283,5 (9) | 309,2 (11,4) | 396,7 (13,6) | 319,6 (14,2) |
| **Water use (m3)** | 6.7 (3.2) | 6 (94,9) | 6,2 (156,9) | 7,1 (199,2) | 5,9 (251,2) | 6,9 (175,3) |
| **Energy use (MJ)** | 61.8 (21.9) | 53,7 (55,1) | 56,5 (53) | 61,9 (52,7) | 58,4 (60,5) | 67,1 (61,7) |
| **Metals and minerals use (kg SB eq)** | 9.7 (4.0) | 8,4 (2,8) | 8,7 (3,1) | 10,1 (3,5) | 9,3 (2,8) | 10,3 (3,2) |

GHG is the emission of greenhouse gases in kg CO_2_ eq. Ionizing radiation is in equivalent of kilobecquerels of Uranium 235. The ozone depletion is in kg CFC-11eq. The photochemical ozone formation is in kg of non-methane volatile organic compounds eq. Particulate matter is in kg of PM_2.5_emitted. The acidification is in mol H^+^ eq. The terrestrial eutrophication is in mol N eq, the freshwater eutrophication is in kg P eq, and the marine eutrophication is in kg N eq. The freshwater ecotoxicity is based on the USEtox model. Land use is in kg C deficit, water use in m^3^, fossils resource use in MJ, and metals and minerals use in kg SB eq.

Supplemental table 7 Percentage of the environmental impact by profile and by protein food groups

|  | **Profile 1: Low meat** | | | | | | | | | | |
| --- | --- | --- | --- | --- | --- | --- | --- | --- | --- | --- | --- |
|  | **Refined grains** | **Whole grains** | **Dairy products** | **Eggs** | **Ruminant meat** | **Pork** | **Poultry** | **Processed meat** | **Fish** | **Fruits - vegetables** | **Pulses** |
| **GHG** | 10,7% (8,6%) | 1,4% (2,8%) | 34,1% (21,4%) | 3,5% (8%) | 3,3% (13,1%) | 0,3% (1,7%) | 0,4% (2,7%) | 19,8% (23,8%) | 11,1% (17,6%) | 15,3% (17,6%) | 0,3% (1,2%) |
| **Ionizing radiation** | 17% (13,4%) | 1,8% (4,1%) | 18,4% (14,9%) | 3,5% (7,7%) | 1,7% (9,4%) | 1% (6,9%) | 0,4% (2,1%) | 19,4% (22,3%) | 9,3% (18,4%) | 26,7% (25,2%) | 0,8% (3,9%) |
| **Ozone depletion** | 13,5% (11%) | 1,6% (3,3%) | 22,4% (17,5%) | 3,5% (8%) | 2,2% (10,9%) | 0,6% (4,5%) | 0,6% (3,6%) | 14,8% (19,7%) | 17,5% (26,9%) | 22,7% (23,4%) | 0,5% (2,5%) |
| **Photochemical ozone** | 11,9% (11%) | 1,5% (3,4%) | 21,2% (18,8%) | 3,5% (8,7%) | 3% (13,1%) | 0,4% (2,6%) | 0,6% (3,7%) | 15,3% (21,6%) | 24,1% (34,8%) | 18% (22,6%) | 0,4% (1,6%) |
| **Particulate matter** | 12,8% (11,4%) | 1,4% (3%) | 25% (19,4%) | 5,7% (11,8%) | 3,4% (14,2%) | 0,8% (5,6%) | 0,6% (3,5%) | 22,5% (25,8%) | 15,4% (25,4%) | 12,4% (18,1%) | 0,2% (0,7%) |
| **Acidification** | 13,2% (12,1%) | 1,3% (2,8%) | 25,2% (20,1%) | 6,1% (12,4%) | 3,5% (14,4%) | 0,7% (5,6%) | 0,6% (3,5%) | 23,5% (26,6%) | 15,6% (25,9%) | 10,2% (17,1%) | 0% (0,2%) |
| **Terrestrial eutrophication** | 13,3% (12%) | 1,4% (3%) | 27,4% (20,6%) | 6,4% (12,8%) | 3,6% (14,6%) | 0,8% (6,2%) | 0,6% (3,5%) | 24,5% (26,8%) | 12,9% (22,8%) | 9% (15,5%) | 0,1% (0,4%) |
| **Freshwater eutrophication** | 18,1% (15,1%) | 2,2% (4,8%) | 21,6% (17,2%) | 4,2% (9,2%) | 2,3% (11,5%) | 0,8% (6,3%) | 0,8% (4,4%) | 15,5% (20,2%) | 15,7% (29%) | 18,2% (22%) | 0,5% (1,9%) |
| **Marine eutrophication** | 17,4% (14,1%) | 2,2% (4,7%) | 24,4% (18,3%) | 3,8% (8,7%) | 3,3% (13,9%) | 0,6% (5%) | 0,5% (3,1%) | 17,4% (22,3%) | 16,8% (27,1%) | 13,2% (18,8%) | 0,4% (1,4%) |
| **Freshwater ecotoxicity** | 13,9% (10,6%) | 1,6% (3,2%) | 26,7% (19,5%) | 5% (10,8%) | 2,5% (11,8%) | 0,8% (6%) | 0,8% (4,3%) | 16,5% (20,4%) | 8,7% (17,8%) | 23,2% (24,2%) | 0,4% (1,6%) |
| **Land use** | 20,7% (15,8%) | 2,9% (6%) | 30% (20,2%) | 4,4% (9,5%) | 3,9% (15,3%) | 0,8% (6,4%) | 0,4% (2,6%) | 20,4% (24,5%) | 4,8% (14%) | 11,3% (17,8%) | 0,4% (1,6%) |
| **Water use** | 11,3% (11,9%) | 1,3% (2,9%) | 19,1% (17,1%) | 5,4% (11,8%) | 2,1% (11%) | 0,8% (6,4%) | 0,6% (3,1%) | 16,5% (21,2%) | 11,2% (24,6%) | 31,3% (26,2%) | 0,3% (1,7%) |
| **Energy use** | 15,7% (12,3%) | 1,6% (3,4%) | 19,9% (15,7%) | 3,3% (7,4%) | 2% (10,4%) | 0,8% (5,9%) | 0,6% (3,3%) | 16,7% (20,6%) | 14,1% (23,3%) | 24,6% (24,1%) | 0,6% (2,9%) |
| **Metals and minerals use** | 15,3% (13,6%) | 1,7% (3,5%) | 19,4% (16%) | 3% (6,8%) | 2,3% (11,1%) | 0,5% (3,8%) | 0,7% (3,8%) | 13,6% (18,7%) | 15,6% (25,7%) | 27,7% (24,8%) | 0,3% (1,3%) |

|  | **Profile 2: Poultry** | | | | | | | | | | |
| --- | --- | --- | --- | --- | --- | --- | --- | --- | --- | --- | --- |
|  | **Refined grains** | **Whole grains** | **Dairy products** | **Eggs** | **Ruminant meat** | **Pork** | **Poultry** | **Processed meat** | **Fish** | **Fruits - vegetables** | **Pulses** |
| **GHG** | 7,5% (6,4%) | 0,5% (1,3%) | 19,8% (14,7%) | 2,2% (7,8%) | 29,7% (30,1%) | 0% (0%) | 16,2% (13,1%) | 12,8% (15%) | 0,9% (4,4%) | 10,3% (12,9%) | 0,1% (0,5%) |
| **Ionizing radiation** | 15,7% (11,3%) | 1,1% (2,9%) | 13,7% (9,8%) | 2,8% (7,9%) | 10,2% (14%) | 0% (0%) | 13,2% (9,8%) | 15,2% (16,1%) | 1% (5,5%) | 26,6% (24,3%) | 0,5% (1,8%) |
| **Ozone depletion** | 12,6% (9,4%) | 0,9% (2,4%) | 17,1% (13,1%) | 2,7% (7,6%) | 13,4% (16,9%) | 0% (0%) | 18,6% (12,5%) | 10,6% (11,9%) | 1,9% (9,2%) | 22% (22,1%) | 0,3% (1,1%) |
| **Photochemical ozone** | 10,6% (8,6%) | 0,8% (1,9%) | 14,7% (11,1%) | 2,6% (8%) | 21,7% (24%) | 0% (0%) | 19% (13,1%) | 10,6% (12,6%) | 2,9% (13,1%) | 16,9% (20,4%) | 0,2% (0,9%) |
| **Particulate matter** | 9,2% (8%) | 0,6% (1,5%) | 14,3% (11,4%) | 3,5% (9,5%) | 26,5% (27,8%) | 0% (0%) | 19,7% (13,9%) | 14,7% (16,2%) | 1,5% (8%) | 9,9% (14,7%) | 0,1% (0,3%) |
| **Acidification** | 9,2% (8,2%) | 0,6% (1,5%) | 13,9% (11,1%) | 3,6% (9,8%) | 27,5% (28,6%) | 0% (0%) | 20,2% (14,5%) | 15,3% (16,8%) | 1,6% (8,2%) | 8,3% (13,8%) | 0% (0,1%) |
| **Terrestrial eutrophication** | 8,9% (7,9%) | 0,5% (1,3%) | 14,8% (12,1%) | 3,6% (9,8%) | 28,2% (29,1%) | 0% (0%) | 20,2% (14,6%) | 15,6% (17%) | 1,2% (6,6%) | 7% (12,2%) | 0,1% (0,2%) |
| **Freshwater eutrophication** | 15,4% (11,4%) | 1,1% (2,7%) | 14,2% (9,9%) | 3,1% (8,5%) | 14,5% (17,6%) | 0% (0%) | 22,1% (13,7%) | 10,6% (11,8%) | 2% (11,3%) | 16,8% (19,3%) | 0,3% (1,2%) |
| **Marine eutrophication** | 14,3% (10,9%) | 1% (2,7%) | 16,4% (12,1%) | 2,7% (8,2%) | 23,9% (25,7%) | 0% (0%) | 15,9% (11,4%) | 11,8% (13,5%) | 1,8% (8,9%) | 11,9% (16,2%) | 0,3% (1,1%) |
| **Freshwater ecotoxicity** | 11,1% (8,3%) | 0,8% (2%) | 16,9% (13,3%) | 3,2% (8,4%) | 15,9% (19,3%) | 0% (0%) | 20,8% (13,9%) | 10,6% (12%) | 0,8% (4,8%) | 19,7% (21,6%) | 0,2% (0,9%) |
| **Land use** | 13,7% (11,1%) | 1,1% (2,9%) | 16,2% (12,8%) | 2,5% (8,4%) | 29,6% (30,3%) | 0% (0%) | 14,9% (12,2%) | 12,6% (15,3%) | 0,3% (2,3%) | 8,8% (14%) | 0,3% (1,3%) |
| **Water use** | 8,2% (7,6%) | 0,6% (1,6%) | 11,4% (9,2%) | 3,6% (8,8%) | 12,2% (15,9%) | 0% (0%) | 21,8% (14,8%) | 10,9% (12,6%) | 1,2% (8,2%) | 29,5% (24,4%) | 0,3% (2%) |
| **Energy use** | 14,2% (10,3%) | 1% (2,5%) | 14,4% (10%) | 2,7% (7,7%) | 11,5% (15,2%) | 0% (0%) | 18,4% (12,1%) | 12,2% (13,3%) | 1,4% (7,2%) | 23,9% (22,7%) | 0,3% (1,2%) |
| **Metals and minerals use** | 14,1% (11,6%) | 0,8% (2,1%) | 13,8% (10,3%) | 2,4% (6,9%) | 13,4% (16,8%) | 0% (0%) | 17,5% (11,9%) | 9,3% (10,9%) | 1,5% (8%) | 26,9% (23,3%) | 0,2% (0,8%) |

|  | **Profile 3: Fish** | | | | | | | | | | |
| --- | --- | --- | --- | --- | --- | --- | --- | --- | --- | --- | --- |
|  | **Refined grains** | **Whole grains** | **Dairy products** | **Eggs** | **Ruminant meat** | **Pork** | **Poultry** | **Processed meat** | **Fish** | **Fruits - vegetables** | **Pulses** |
| **GHG** | 5,4% (5%) | 0,7% (1,7%) | 15,1% (11,6%) | 2,3% (8,4%) | 36% (29%) | 0,3% (2,5%) | 8,7% (11,6%) | 8,9% (13,4%) | 14,2% (14,1%) | 8,4% (9,5%) | 0,1% (0,4%) |
| **Ionizing radiation** | 11,6% (9,7%) | 1,3% (3%) | 11,7% (11,3%) | 3% (9,8%) | 11% (14,6%) | 0,3% (3%) | 6,9% (9,5%) | 10,7% (14,3%) | 19,9% (26,7%) | 23,2% (21,8%) | 0,5% (1,7%) |
| **Ozone depletion** | 7,4% (6,8%) | 0,8% (2,1%) | 12,5% (12,9%) | 2,5% (8,7%) | 12,5% (16,3%) | 0,1% (1,4%) | 8,2% (11,2%) | 6,2% (9,6%) | 33,3% (29,2%) | 16,3% (19,5%) | 0,2% (0,6%) |
| **Photochemical ozone** | 4,9% (5,3%) | 0,6% (1,8%) | 8,1% (9,2%) | 2% (7,7%) | 17,2% (20,7%) | 0,1% (0,8%) | 7,2% (10,8%) | 4,9% (8,6%) | 44,1% (31,8%) | 10,9% (17%) | 0,1% (0,3%) |
| **Particulate matter** | 5,8% (6,1%) | 0,6% (1,8%) | 10,5% (10,7%) | 3,2% (10,1%) | 28% (27,1%) | 0,2% (2,2%) | 9,8% (13,7%) | 9% (13,1%) | 25,6% (29%) | 7,3% (12,2%) | 0% (0,2%) |
| **Acidification** | 5,8% (6,2%) | 0,6% (1,7%) | 10,2% (10,5%) | 3,3% (10,3%) | 29,3% (27,9%) | 0,2% (2,2%) | 10,1% (14,3%) | 9,4% (13,6%) | 25,6% (29,2%) | 5,6% (10,7%) | 0% (0,1%) |
| **Terrestrial eutrophication** | 5,9% (6,4%) | 0,6% (1,7%) | 11,3% (11,4%) | 3,5% (10,5%) | 31,4% (28,9%) | 0,2% (2,6%) | 10,6% (14,8%) | 10% (14,3%) | 21,3% (27,8%) | 5,1% (9,5%) | 0% (0,1%) |
| **Freshwater eutrophication** | 8,9% (9,5%) | 1% (3%) | 9,6% (9,9%) | 2,9% (10%) | 14% (17,8%) | 0,2% (2,2%) | 9,1% (12,3%) | 5,9% (9,6%) | 36% (35%) | 12,1% (16,6%) | 0,2% (0,7%) |
| **Marine eutrophication** | 8,5% (8,3%) | 1,1% (3,1%) | 11,3% (11,1%) | 2,5% (9,1%) | 24,1% (24,3%) | 0,1% (1,9%) | 7,2% (9,9%) | 6,9% (10,8%) | 30,1% (29,5%) | 8,1% (13,1%) | 0,1% (0,5%) |
| **Freshwater ecotoxicity** | 8,2% (7,1%) | 0,9% (2,3%) | 15% (14,4%) | 3,3% (10,2%) | 18,1% (20,1%) | 0,2% (1,9%) | 11% (14%) | 7,4% (10,8%) | 18,2% (25,9%) | 17,4% (19,6%) | 0,2% (0,6%) |
| **Land use** | 10,5% (10,3%) | 1,5% (4%) | 13,8% (12,6%) | 2,7% (9,5%) | 36,7% (30,4%) | 0,3% (2,7%) | 8,7% (12,6%) | 9,2% (14,1%) | 9,8% (20,2%) | 6,6% (11,3%) | 0,2% (0,7%) |
| **Water use** | 6% (8,2%) | 0,6% (1,7%) | 9% (9,7%) | 3,6% (11%) | 13% (16,8%) | 0,2% (2,2%) | 10,1% (13,5%) | 6,9% (11%) | 25,4% (32,4%) | 25% (23,3%) | 0,2% (0,9%) |
| **Energy use** | 9,3% (8%) | 1% (2,5%) | 11,1% (10,7%) | 2,6% (9,1%) | 11,8% (15,5%) | 0,2% (2,1%) | 8,7% (11,4%) | 7,9% (11,3%) | 27,6% (27,9%) | 19,5% (20,4%) | 0,3% (0,9%) |
| **Metals and minerals use** | 8,1% (8,3%) | 0,9% (2,3%) | 10% (10,7%) | 2,1% (8,4%) | 12,5% (16,5%) | 0,1% (1,6%) | 7,7% (10,5%) | 5,2% (8%) | 32,4% (29,6%) | 20,8% (21,3%) | 0,1% (0,4%) |

|  | **Profile 4: Ruminant meat** | | | | | | | | | | |
| --- | --- | --- | --- | --- | --- | --- | --- | --- | --- | --- | --- |
|  | **Refined grains** | **Whole grains** | **Dairy products** | **Eggs** | **Ruminant meat** | **Pork** | **Poultry** | **Processed meat** | **Fish** | **Fruits - vegetables** | **Pulses** |
| **GHG** | 4,7% (4,4%) | 0,5% (1,2%) | 12,4% (9,4%) | 1% (2,3%) | 58,7% (20,5%) | 0,6% (2,8%) | 1,2% (4,3%) | 12,2% (14,9%) | 1,7% (4,9%) | 6,9% (10,6%) | 0,1% (0,4%) |
| **Ionizing radiation** | 14,1% (11,1%) | 1,2% (2,7%) | 11,7% (9,7%) | 2,1% (5%) | 23,8% (20,8%) | 1,6% (7,1%) | 1,4% (4,9%) | 18,1% (17,7%) | 2,9% (12%) | 22,6% (22,2%) | 0,5% (2,1%) |
| **Ozone depletion** | 11,4% (9,5%) | 1% (2,3%) | 15,4% (13,1%) | 1,9% (4,3%) | 30,8% (21,5%) | 0,8% (3,8%) | 1,8% (6,3%) | 12,7% (13,9%) | 5,4% (15,8%) | 18,5% (20,7%) | 0,3% (1,3%) |
| **Photochemical ozone** | 8% (7,3%) | 0,8% (1,9%) | 10,9% (9,3%) | 1,6% (3,6%) | 45% (23,9%) | 0,5% (2,5%) | 1,5% (5,8%) | 11,1% (13,6%) | 7,4% (19,9%) | 13% (18,9%) | 0,2% (0,7%) |
| **Particulate matter** | 6,3% (6%) | 0,5% (1,3%) | 9,4% (7,7%) | 2,1% (4,7%) | 54,3% (22,8%) | 0,8% (3,9%) | 1,5% (5,5%) | 14,4% (15,9%) | 3,6% (12,9%) | 7% (12,7%) | 0% (0,2%) |
| **Acidification** | 6,1% (5,9%) | 0,5% (1,3%) | 9,1% (7,5%) | 2,1% (4,8%) | 56,1% (22,6%) | 0,8% (3,8%) | 1,5% (5,4%) | 14,6% (16,2%) | 3,6% (12,9%) | 5,6% (11,8%) | 0% (0,2%) |
| **Terrestrial eutrophication** | 5,8% (5,7%) | 0,5% (1,2%) | 9,3% (7,5%) | 2,1% (4,9%) | 57,8% (22,3%) | 0,9% (4,1%) | 1,4% (5,3%) | 14,8% (16,3%) | 2,8% (11,7%) | 4,5% (10,1%) | 0% (0,1%) |
| **Freshwater eutrophication** | 13,9% (11,5%) | 1,4% (3%) | 13% (10,3%) | 2,1% (4,5%) | 34,3% (22,1%) | 0,9% (4,1%) | 2,1% (7,2%) | 12,9% (13,8%) | 4,4% (16,2%) | 14,7% (18,7%) | 0,3% (1,1%) |
| **Marine eutrophication** | 10% (8,5%) | 1,1% (2,5%) | 11,3% (9,1%) | 1,5% (3,2%) | 50,1% (22,6%) | 0,7% (3%) | 1,2% (4,9%) | 12,1% (14,3%) | 3,8% (13,4%) | 8% (14,1%) | 0,1% (0,7%) |
| **Freshwater ecotoxicity** | 9,4% (7,8%) | 0,8% (1,9%) | 15,6% (13,2%) | 2,3% (5,3%) | 37,3% (22,6%) | 0,9% (4,3%) | 1,9% (6,7%) | 12,4% (13,7%) | 2,5% (11,1%) | 16,6% (19,7%) | 0,2% (0,7%) |
| **Land use** | 8,2% (7%) | 0,9% (2,1%) | 9,5% (7,9%) | 1,2% (2,8%) | 60,2% (21,4%) | 0,7% (3,3%) | 1% (4%) | 11,7% (15%) | 1,2% (8,3%) | 5,3% (11,2%) | 0,1% (0,6%) |
| **Water use** | 7,4% (8,1%) | 0,8% (2%) | 11,3% (9,8%) | 3% (6,9%) | 31,1% (22,3%) | 1% (4,7%) | 2,1% (6,7%) | 13,9% (15%) | 3,2% (14,2%) | 26,1% (23,4%) | 0,2% (0,8%) |
| **Energy use** | 13,3% (10,6%) | 1,1% (2,7%) | 12,7% (10,1%) | 1,9% (4,5%) | 27,6% (21,2%) | 1,1% (5%) | 1,8% (6,3%) | 15% (15,5%) | 4,3% (14,1%) | 20,8% (21,3%) | 0,3% (1,5%) |
| **Metals and minerals use** | 13,1% (11,8%) | 1,1% (2,5%) | 12,5% (11,2%) | 1,5% (3,4%) | 30,9% (21,7%) | 0,6% (2,7%) | 1,7% (6,1%) | 11,1% (12,9%) | 5,3% (15,2%) | 22,1% (21,9%) | 0,1% (0,5%) |

|  | **Profile 5: Pork** | | | | | | | | | | |
| --- | --- | --- | --- | --- | --- | --- | --- | --- | --- | --- | --- |
|  | **Refined grains** | **Whole grains** | **Dairy products** | **Eggs** | **Ruminant meat** | **Pork** | **Poultry** | **Processed meat** | **Fish** | **Fruits - vegetables** | **Pulses** |
| **GHG** | 5,4% (4%) | 0,5% (1,1%) | 18% (12,7%) | 1,4% (2,9%) | 25,8% (27,8%) | 16,4% (12,5%) | 5,2% (8,6%) | 11,1% (14,1%) | 5,9% (10,7%) | 10,1% (13,8%) | 0,2% (0,5%) |
| **Ionizing radiation** | 9% (6,8%) | 0,7% (1,9%) | 9,5% (8%) | 1,4% (3%) | 7,6% (14,4%) | 27,1% (20%) | 3,4% (5,8%) | 10,8% (13,7%) | 6,2% (16,4%) | 23,7% (24,1%) | 0,6% (2,3%) |
| **Ozone depletion** | 7,8% (6,1%) | 0,7% (1,7%) | 13,7% (11,4%) | 1,5% (3,3%) | 10,9% (17,3%) | 18,7% (17,6%) | 5,3% (8,9%) | 8,1% (11,1%) | 12,2% (22,7%) | 20,8% (23,1%) | 0,4% (1,3%) |
| **Photochemical ozone** | 6,3% (5,5%) | 0,6% (1,6%) | 11,3% (10,1%) | 1,5% (3,5%) | 17,1% (22,9%) | 15,9% (17,1%) | 5,6% (9,8%) | 7,9% (11,3%) | 17,4% (28,7%) | 16,2% (22,2%) | 0,2% (0,8%) |
| **Particulate matter** | 5,6% (4,6%) | 0,4% (1,2%) | 11,5% (9,8%) | 2,1% (4,4%) | 21,8% (26,1%) | 21,8% (18,7%) | 5,8% (9,5%) | 11,2% (14%) | 9,7% (20,7%) | 10,2% (17,3%) | 0,1% (0,3%) |
| **Acidification** | 5,6% (4,6%) | 0,4% (1,1%) | 11,3% (9,7%) | 2,2% (4,6%) | 22,7% (26,9%) | 22% (18,9%) | 5,9% (9,7%) | 11,7% (14,4%) | 9,7% (20,8%) | 8,4% (16%) | 0% (0,2%) |
| **Terrestrial eutrophication** | 5,5% (4,6%) | 0,4% (1,1%) | 12,1% (10,3%) | 2,2% (4,7%) | 23,6% (27,4%) | 22,9% (19%) | 6% (9,7%) | 12,1% (14,8%) | 8% (18,8%) | 7,3% (14,7%) | 0% (0,2%) |
| **Freshwater eutrophication** | 9,7% (7,9%) | 0,9% (2,5%) | 11,4% (9,5%) | 1,8% (4%) | 12,2% (18,4%) | 19% (17,5%) | 6,7% (10,7%) | 8,2% (11,3%) | 13% (26,1%) | 16,9% (21,6%) | 0,4% (1,2%) |
| **Marine eutrophication** | 8,9% (6,9%) | 0,9% (2,3%) | 13,7% (11,3%) | 1,5% (3,4%) | 19,9% (24,6%) | 17,6% (17,1%) | 4,8% (8,2%) | 9,2% (12,3%) | 11,6% (22,2%) | 11,7% (18,5%) | 0,2% (0,8%) |
| **Freshwater ecotoxicity** | 7,8% (6,4%) | 0,6% (1,5%) | 14,9% (12,2%) | 1,9% (4,2%) | 13,6% (19,4%) | 19,8% (17,7%) | 6,2% (10%) | 8,6% (11,6%) | 6,1% (16,2%) | 20,3% (22,9%) | 0,3% (0,8%) |
| **Land use** | 9,6% (7,7%) | 0,9% (2,3%) | 14,8% (12,5%) | 1,5% (3,2%) | 26,2% (29,3%) | 18,8% (17,8%) | 4,7% (7,9%) | 10,6% (14,2%) | 3,2% (12%) | 9,5% (16,5%) | 0,2% (0,8%) |
| **Water use** | 5,6% (6,3%) | 0,5% (1,5%) | 9,2% (8,1%) | 2,2% (5%) | 10,5% (17,1%) | 19,5% (18%) | 6,5% (10,7%) | 8,5% (11,9%) | 8,8% (21,6%) | 28,3% (25,5%) | 0,3% (1,3%) |
| **Energy use** | 8,6% (6,5%) | 0,7% (1,8%) | 10,9% (8,8%) | 1,4% (3,1%) | 9,3% (15,8%) | 22,6% (18,6%) | 5,1% (8,5%) | 9,2% (12,2%) | 9,5% (19,7%) | 22,4% (23,5%) | 0,4% (1,6%) |
| **Metals and minerals use** | 8,8% (7,5%) | 0,7% (1,9%) | 11,7% (10,1%) | 1,4% (3,1%) | 11,3% (17,8%) | 16,2% (16,9%) | 5,3% (9%) | 7,2% (10,4%) | 12,1% (23%) | 25,1% (24,6%) | 0,2% (0,8%) |

Supplemental table 8 Mean (SD) of nutritional and health indicators and Mean (SD) of probabilities of nutritional adequacy by profile

|  | **Overall** | **Profile 1: Low meat** | **Profile 2: Poultry** | **Profile 3: Fish** | **Profile 4: Ruminant meat** | **Profile 5: Pork** |
| --- | --- | --- | --- | --- | --- | --- |
| **PANDiet** | 68.6 (5.9) | 66.9 (5.7) | 67.2 (5.5) | 69.9 (6.2) | 67.2 (6.1) | 69.9 (5.2) |
| **SecDiet** | 0.952 (0.081) | 0.946 (0.078) | 0.948 (0.08) | 0.953 (0.077) | 0.936 (0.118) | 0.967 (0.056) |
| **HiDiet** | 0.004 (0.12) | 0.031 (0.11) | 0.018 (0.124) | 0.027 (0.122) | -0.038 (0.107) | -0.021 (0.116) |
| **LAMD** | 8.1 (2.8) | 8.8 (2.9) | 7.3 (2.6) | 9 (2.8) | 7.4 (2.7) | 7.8 (2.5) |
| **AHEI** | 43.4 (12.2) | 43.9 (11.6) | 42.3 (13.6) | 46.5 (12.2) | 40 (10.6) | 42.6 (11.3) |
| **sPNNS-GS2** | 6.3 (1.8) | 5.9 (1.6) | 6.1 (2.1) | 6.4 (1.8) | 6 (1.5) | 6.5 (1.7) |

Supplemental table 9. Correlations between protein food groups consumptions, nutritional, health and, environmental factors

|  | Refined grains | Whole grains | Dairy | Eggs | Ruminant meat | Pork | Poultry | Processed meat | Fish | Fruits and vegetables | Pulses | PANDiet | SecDiet | HiDiet | LAMD | AHEI | sPNNSGS2 | GHG | Score EF | Ionizing radiation | Ozone depletion | Photochemical ozone | Particulate matter | Acidification | Terrestrial eutrophication | Freshwater eutrophication | Marine eutrophication | Freshwater ecotoxicity | Land use | Water use | Energy use | Metals and minerals use |
| --- | --- | --- | --- | --- | --- | --- | --- | --- | --- | --- | --- | --- | --- | --- | --- | --- | --- | --- | --- | --- | --- | --- | --- | --- | --- | --- | --- | --- | --- | --- | --- | --- |
| Refined grains | 1 | -0.12 | 0.28 | 0.05 | 0.04 | 0.07 | 0.04 | 0.17 | 0.05 | 0.07 | 0.07 | -0.01 | 0.18 | -0.05 | 0.11 | -0.03 | -0.09 | 0.06 | 0.10 | 0.13 | 0.05 | 0.06 | 0.07 | 0.07 | 0.06 | 0.07 | 0.10 | 0.18 | 0.07 | 0.16 | 0.11 | 0.09 |
| Whole grains | -0.12 | 1 | 0.15 | 0.02 | -0.06 | 0.00 | -0.03 | 0.06 | 0.05 | 0.05 | 0.06 | 0.11 | 0.08 | 0.38 | 0.15 | 0.30 | 0.18 | -0.08 | -0.04 | 0.00 | 0.02 | 0.01 | -0.06 | -0.06 | -0.07 | 0.03 | -0.05 | 0.03 | -0.07 | 0.06 | 0.00 | 0.03 |
| Dairy | 0.28 | 0.15 | 1 | 0.06 | 0.05 | 0.06 | -0.02 | 0.17 | -0.01 | 0.04 | 0.08 | -0.09 | 0.20 | 0.01 | 0.01 | 0.03 | -0.06 | 0.08 | 0.11 | 0.13 | 0.02 | 0.07 | 0.10 | 0.09 | 0.08 | 0.01 | 0.09 | 0.18 | 0.08 | 0.05 | 0.11 | 0.13 |
| Eggs | 0.05 | 0.02 | 0.06 | 1 | -0.03 | -0.01 | 0.01 | 0.04 | 0.10 | 0.01 | -0.02 | 0.05 | 0.10 | -0.01 | 0.05 | 0.02 | 0.01 | 0.01 | 0.02 | 0.04 | 0.01 | 0.01 | 0.02 | 0.02 | 0.02 | 0.01 | 0.01 | 0.04 | 0.00 | 0.04 | 0.04 | 0.07 |
| Ruminant meat | 0.04 | -0.06 | 0.05 | -0.03 | 1 | -0.02 | -0.03 | -0.02 | -0.01 | -0.05 | 0.04 | 0.06 | -0.02 | -0.19 | -0.12 | -0.13 | 0.02 | 0.48 | 0.40 | 0.14 | 0.03 | 0.18 | 0.43 | 0.44 | 0.46 | 0.11 | 0.36 | 0.19 | 0.50 | 0.06 | 0.16 | 0.14 |
| Pork | 0.07 | 0.00 | 0.06 | -0.01 | -0.02 | 1 | -0.07 | 0.09 | -0.01 | -0.02 | 0.06 | 0.07 | 0.05 | -0.10 | -0.07 | -0.02 | 0.05 | 0.03 | 0.09 | 0.28 | 0.00 | 0.06 | 0.09 | 0.09 | 0.08 | 0.03 | 0.09 | 0.15 | 0.04 | 0.02 | 0.18 | 0.07 |
| Poultry | 0.04 | -0.03 | -0.02 | 0.01 | -0.03 | -0.07 | 1 | -0.02 | -0.04 | 0.00 | 0.06 | 0.11 | 0.08 | -0.01 | -0.12 | 0.04 | 0.04 | 0.01 | 0.05 | 0.04 | 0.03 | -0.02 | 0.04 | 0.04 | 0.04 | 0.06 | 0.03 | 0.06 | 0.04 | 0.06 | 0.07 | 0.04 |
| Processed meat | 0.17 | 0.06 | 0.17 | 0.04 | -0.02 | 0.09 | -0.02 | 1 | -0.05 | -0.02 | 0.03 | -0.06 | 0.13 | -0.22 | -0.14 | -0.22 | -0.16 | 0.08 | 0.11 | 0.21 | -0.03 | 0.03 | 0.10 | 0.09 | 0.10 | 0.04 | 0.10 | 0.18 | 0.07 | 0.05 | 0.16 | 0.09 |
| Fish | 0.05 | 0.05 | -0.01 | 0.10 | -0.01 | -0.01 | -0.04 | -0.05 | 1 | 0.09 | 0.01 | 0.24 | 0.04 | 0.20 | 0.32 | 0.29 | 0.16 | 0.05 | 0.09 | 0.05 | 0.10 | 0.31 | 0.07 | 0.06 | 0.02 | 0.30 | 0.15 | 0.06 | -0.03 | 0.18 | 0.14 | 0.21 |
| Fruits and vegetables | 0.07 | 0.05 | 0.04 | 0.01 | -0.05 | -0.02 | 0.00 | -0.02 | 0.09 | 1 | 0.03 | 0.13 | 0.08 | 0.18 | 0.17 | 0.25 | 0.08 | -0.04 | -0.01 | 0.06 | 0.13 | 0.04 | -0.04 | -0.05 | -0.06 | 0.03 | 0.04 | 0.01 | -0.06 | 0.17 | 0.07 | 0.10 |
| Pulses | 0.07 | 0.06 | 0.08 | -0.02 | 0.04 | 0.06 | 0.06 | 0.03 | 0.01 | 0.03 | 1 | 0.09 | 0.07 | 0.07 | 0.21 | 0.23 | -0.01 | 0.05 | 0.06 | 0.04 | 0.02 | 0.08 | 0.06 | 0.06 | 0.05 | 0.01 | 0.05 | 0.06 | 0.05 | 0.06 | 0.04 | 0.07 |
| PANDiet | -0.01 | 0.11 | -0.09 | 0.05 | 0.06 | 0.07 | 0.11 | -0.06 | 0.24 | 0.13 | 0.09 | 1 | 0.46 | 0.29 | 0.30 | 0.36 | 0.50 | 0.05 | 0.12 | 0.11 | 0.03 | 0.21 | 0.08 | 0.07 | 0.05 | 0.23 | 0.15 | 0.12 | 0.03 | 0.25 | 0.17 | 0.21 |
| SecDiet | 0.18 | 0.08 | 0.20 | 0.10 | -0.02 | 0.05 | 0.08 | 0.13 | 0.04 | 0.08 | 0.07 | 0.46 | 1 | 0.10 | 0.15 | 0.05 | 0.06 | 0.15 | 0.21 | 0.25 | 0.04 | 0.19 | 0.16 | 0.15 | 0.14 | 0.19 | 0.20 | 0.27 | 0.11 | 0.25 | 0.25 | 0.25 |
| HiDiet | -0.05 | 0.38 | 0.01 | -0.01 | -0.19 | -0.10 | -0.01 | -0.22 | 0.20 | 0.18 | 0.07 | 0.29 | 0.10 | 1 | 0.40 | 0.54 | 0.34 | -0.20 | -0.17 | -0.11 | 0.04 | 0.02 | -0.22 | -0.23 | -0.25 | 0.08 | -0.14 | -0.11 | -0.25 | 0.20 | -0.06 | 0.00 |
| LAMD | 0.11 | 0.15 | 0.01 | 0.05 | -0.12 | -0.07 | -0.12 | -0.14 | 0.32 | 0.17 | 0.21 | 0.30 | 0.15 | 0.40 | 1 | 0.63 | 0.37 | -0.15 | -0.07 | -0.04 | 0.04 | 0.19 | -0.12 | -0.13 | -0.18 | 0.12 | -0.06 | -0.01 | -0.20 | 0.32 | 0.02 | 0.12 |
| AHEI | -0.03 | 0.30 | 0.03 | 0.02 | -0.13 | -0.02 | 0.04 | -0.22 | 0.29 | 0.25 | 0.23 | 0.36 | 0.05 | 0.54 | 0.63 | 1 | 0.49 | -0.20 | -0.12 | -0.09 | 0.05 | 0.07 | -0.17 | -0.18 | -0.21 | 0.10 | -0.07 | -0.03 | -0.22 | 0.26 | -0.04 | 0.07 |
| sPNNSGS2 | -0.09 | 0.18 | -0.06 | 0.01 | 0.02 | 0.05 | 0.04 | -0.16 | 0.16 | 0.08 | -0.01 | 0.50 | 0.06 | 0.34 | 0.37 | 0.49 | 1 | -0.09 | -0.05 | -0.05 | 0.01 | 0.04 | -0.07 | -0.07 | -0.09 | 0.07 | -0.05 | -0.07 | -0.09 | 0.21 | -0.02 | 0.03 |
| GHG | 0.06 | -0.08 | 0.08 | 0.01 | 0.48 | 0.03 | 0.01 | 0.08 | 0.05 | -0.04 | 0.05 | 0.05 | 0.15 | -0.20 | -0.15 | -0.20 | -0.09 | 1 | 0.95 | 0.62 | 0.18 | 0.59 | 0.95 | 0.95 | 0.96 | 0.45 | 0.85 | 0.66 | 0.94 | 0.33 | 0.68 | 0.56 |
| Score EF | 0.10 | -0.04 | 0.11 | 0.02 | 0.40 | 0.09 | 0.05 | 0.11 | 0.09 | -0.01 | 0.06 | 0.12 | 0.21 | -0.17 | -0.07 | -0.12 | -0.05 | 0.95 | 1 | 0.76 | 0.21 | 0.70 | 0.98 | 0.97 | 0.96 | 0.56 | 0.91 | 0.79 | 0.92 | 0.47 | 0.82 | 0.70 |
| Ionizing radiation | 0.13 | 0.00 | 0.13 | 0.04 | 0.14 | 0.28 | 0.04 | 0.21 | 0.05 | 0.06 | 0.04 | 0.11 | 0.25 | -0.11 | -0.04 | -0.09 | -0.05 | 0.62 | 0.76 | 1 | 0.17 | 0.53 | 0.68 | 0.65 | 0.63 | 0.52 | 0.68 | 0.72 | 0.57 | 0.46 | 0.93 | 0.68 |
| Ozone depletion | 0.05 | 0.02 | 0.02 | 0.01 | 0.03 | 0.00 | 0.03 | -0.03 | 0.10 | 0.13 | 0.02 | 0.03 | 0.04 | 0.04 | 0.04 | 0.05 | 0.01 | 0.18 | 0.21 | 0.17 | 1 | 0.18 | 0.18 | 0.17 | 0.16 | 0.16 | 0.18 | 0.20 | 0.14 | 0.15 | 0.23 | 0.22 |
| Photochemical ozone | 0.06 | 0.01 | 0.07 | 0.01 | 0.18 | 0.06 | -0.02 | 0.03 | 0.31 | 0.04 | 0.08 | 0.21 | 0.19 | 0.02 | 0.19 | 0.07 | 0.04 | 0.59 | 0.70 | 0.53 | 0.18 | 1 | 0.70 | 0.69 | 0.59 | 0.41 | 0.68 | 0.52 | 0.45 | 0.34 | 0.72 | 0.67 |
| Particulate matter | 0.07 | -0.06 | 0.10 | 0.02 | 0.43 | 0.09 | 0.04 | 0.10 | 0.07 | -0.04 | 0.06 | 0.08 | 0.16 | -0.22 | -0.12 | -0.17 | -0.07 | 0.95 | 0.98 | 0.68 | 0.18 | 0.70 | 1 | 1.00 | 0.99 | 0.46 | 0.88 | 0.70 | 0.94 | 0.34 | 0.75 | 0.63 |
| Acidification | 0.07 | -0.06 | 0.09 | 0.02 | 0.44 | 0.09 | 0.04 | 0.09 | 0.06 | -0.05 | 0.06 | 0.07 | 0.15 | -0.23 | -0.13 | -0.18 | -0.07 | 0.95 | 0.97 | 0.65 | 0.17 | 0.69 | 1.00 | 1 | 0.99 | 0.44 | 0.87 | 0.68 | 0.95 | 0.32 | 0.71 | 0.59 |
| Terrestrial eutrophication | 0.06 | -0.07 | 0.08 | 0.02 | 0.46 | 0.08 | 0.04 | 0.10 | 0.02 | -0.06 | 0.05 | 0.05 | 0.14 | -0.25 | -0.18 | -0.21 | -0.09 | 0.96 | 0.96 | 0.63 | 0.16 | 0.59 | 0.99 | 0.99 | 1 | 0.42 | 0.85 | 0.66 | 0.98 | 0.30 | 0.66 | 0.54 |
| Freshwater eutrophication | 0.07 | 0.03 | 0.01 | 0.01 | 0.11 | 0.03 | 0.06 | 0.04 | 0.30 | 0.03 | 0.01 | 0.23 | 0.19 | 0.08 | 0.12 | 0.10 | 0.07 | 0.45 | 0.56 | 0.52 | 0.16 | 0.41 | 0.46 | 0.44 | 0.42 | 1 | 0.61 | 0.50 | 0.41 | 0.51 | 0.58 | 0.49 |
| Marine eutrophication | 0.10 | -0.05 | 0.09 | 0.01 | 0.36 | 0.09 | 0.03 | 0.10 | 0.15 | 0.04 | 0.05 | 0.15 | 0.20 | -0.14 | -0.06 | -0.07 | -0.05 | 0.85 | 0.91 | 0.68 | 0.18 | 0.68 | 0.88 | 0.87 | 0.85 | 0.61 | 1 | 0.73 | 0.82 | 0.41 | 0.75 | 0.65 |
| Freshwater ecotoxicity | 0.18 | 0.03 | 0.18 | 0.04 | 0.19 | 0.15 | 0.06 | 0.18 | 0.06 | 0.01 | 0.06 | 0.12 | 0.27 | -0.11 | -0.01 | -0.03 | -0.07 | 0.66 | 0.79 | 0.72 | 0.20 | 0.52 | 0.70 | 0.68 | 0.66 | 0.50 | 0.73 | 1 | 0.63 | 0.47 | 0.76 | 0.73 |
| Land use | 0.07 | -0.07 | 0.08 | 0.00 | 0.50 | 0.04 | 0.04 | 0.07 | -0.03 | -0.06 | 0.05 | 0.03 | 0.11 | -0.25 | -0.20 | -0.22 | -0.09 | 0.94 | 0.92 | 0.57 | 0.14 | 0.45 | 0.94 | 0.95 | 0.98 | 0.41 | 0.82 | 0.63 | 1 | 0.28 | 0.58 | 0.47 |
| Water use | 0.16 | 0.06 | 0.05 | 0.04 | 0.06 | 0.02 | 0.06 | 0.05 | 0.18 | 0.17 | 0.06 | 0.25 | 0.25 | 0.20 | 0.32 | 0.26 | 0.21 | 0.33 | 0.47 | 0.46 | 0.15 | 0.34 | 0.34 | 0.32 | 0.30 | 0.51 | 0.41 | 0.47 | 0.28 | 1 | 0.49 | 0.46 |
| Energy use | 0.11 | 0.00 | 0.11 | 0.04 | 0.16 | 0.18 | 0.07 | 0.16 | 0.14 | 0.07 | 0.04 | 0.17 | 0.25 | -0.06 | 0.02 | -0.04 | -0.02 | 0.68 | 0.82 | 0.93 | 0.23 | 0.72 | 0.75 | 0.71 | 0.66 | 0.58 | 0.75 | 0.76 | 0.58 | 0.49 | 1 | 0.79 |
| Metals and minerals use | 0.09 | 0.03 | 0.13 | 0.07 | 0.14 | 0.07 | 0.04 | 0.09 | 0.21 | 0.10 | 0.07 | 0.21 | 0.25 | 0.00 | 0.12 | 0.07 | 0.03 | 0.56 | 0.70 | 0.68 | 0.22 | 0.67 | 0.63 | 0.59 | 0.54 | 0.49 | 0.65 | 0.73 | 0.47 | 0.46 | 0.79 | 1 |

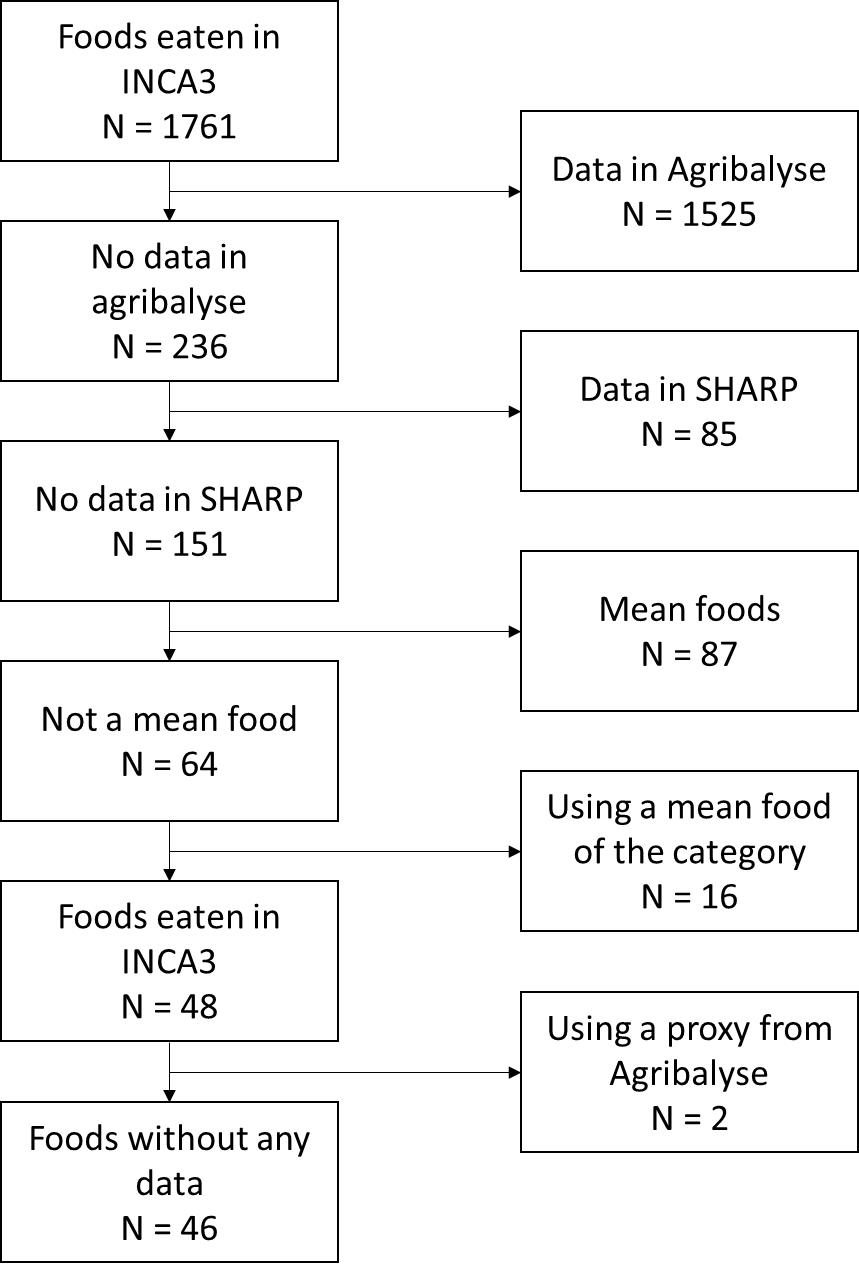

Supplemental Figure 1 Data attribution of the 1761 foods in INCA3 for the greenhouse gases.

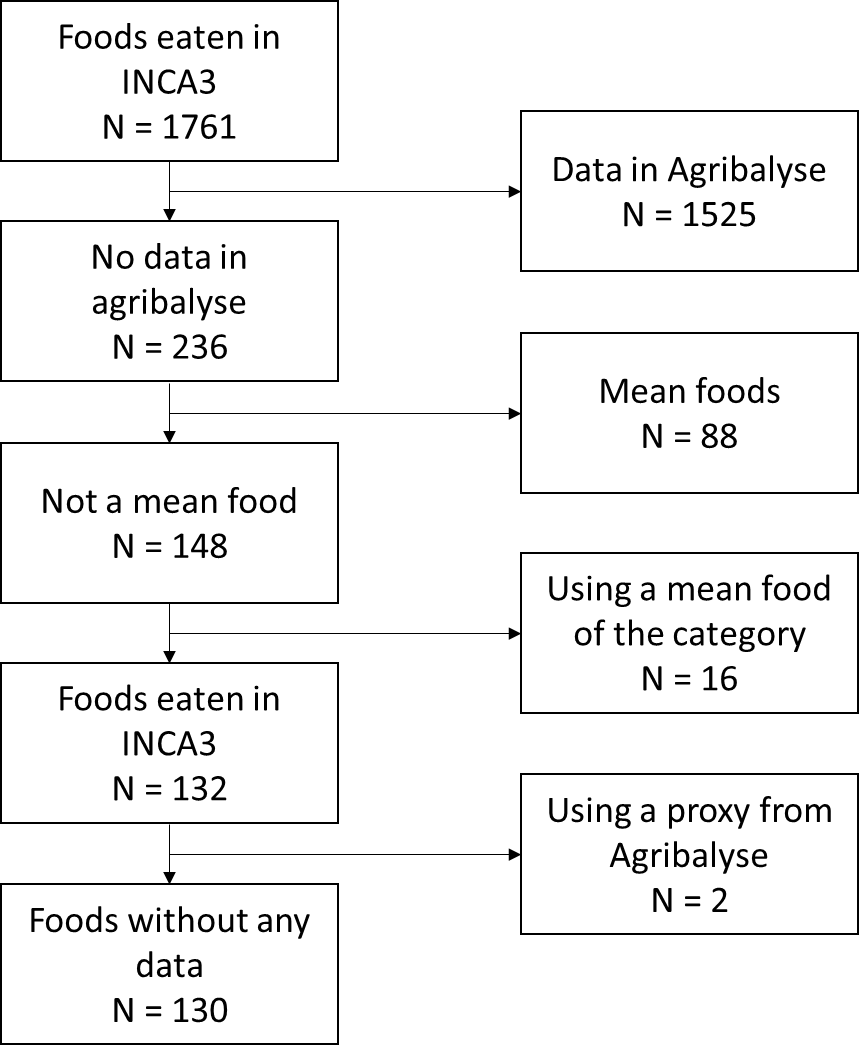

Supplemental Figure 2 Data attribution of the 1761 foods in INCA3 for the other environmental indicators.

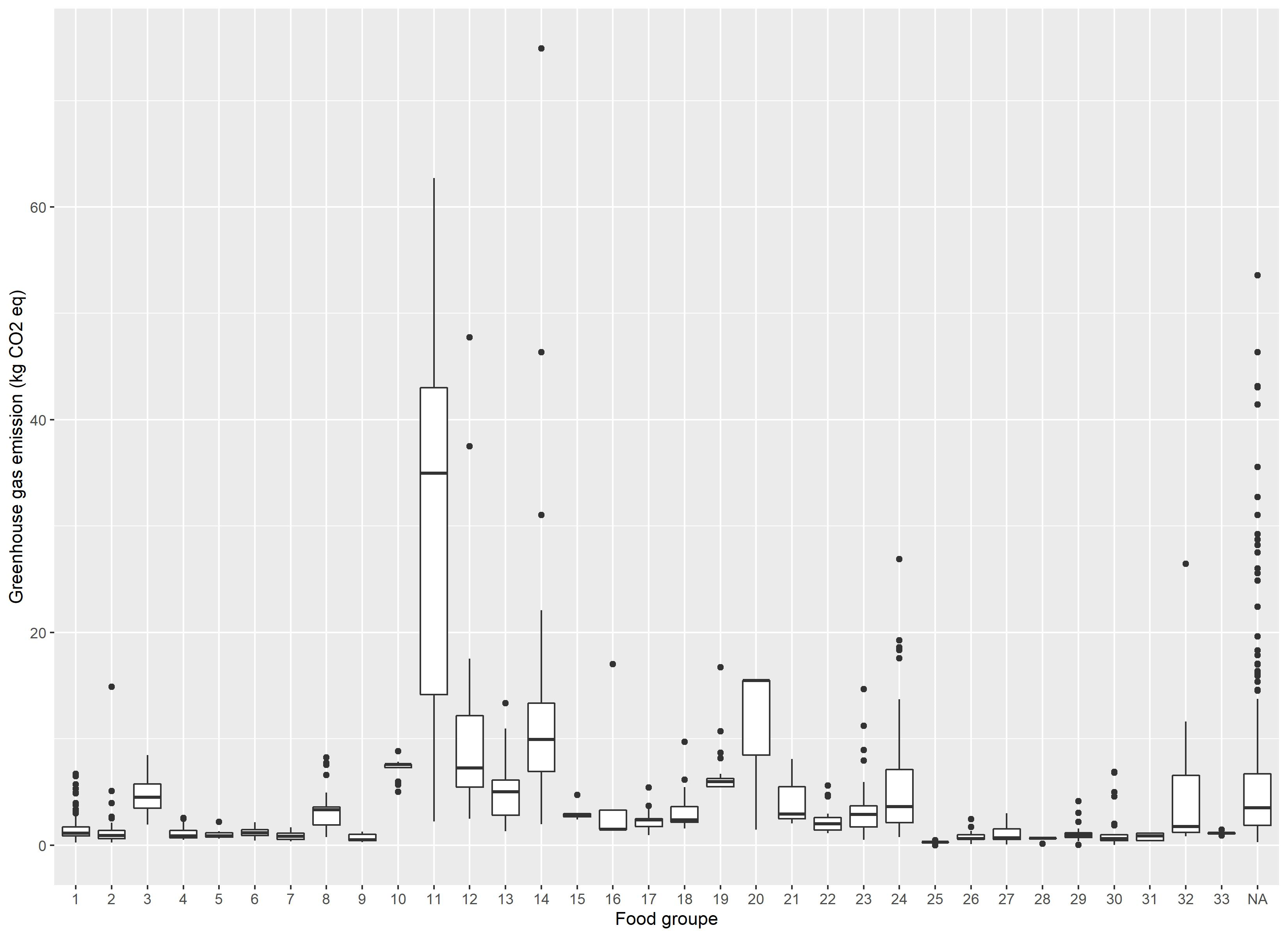

Supplemental Figure 3 Variability of Greenhouse gas emission by food group (1: Vegetable, 2: Fruits, 3: Nuts, seeds and, oleaginous fruits, 4: Refined bread and bread products, 5: Whole and semi-wholemeal bread and bread products, 6: Other refined starchy foods, 7: Other whole and semi-wholemeal starchy foods, 8: Starch products, processed, 9: Legumes, 10: Poultry, 11: Meat excluding poultry, 12: Delicatessen, 13: Fatty fish, 14: Lean fish, 15: Eggs and egg dishes, 16: Milk, 17: Fresh dairy products, 18: Sweet dairy desserts, 19: Cheese, 20: Animal fats, 21: Vegetable fats rich in ALA, 22: Vegetable fats low in ALA, 23: Sauces and fresh creams, 24: Sweet products or sweet and fatty products, 25: Drinking water, 26: Sweet drinks and fruit juices, 27: Hot drinks, 28:Salt, 29: Condiments, 30: Soups and broths, 31: Animal products substitutes, 32: Others foods, 33: Alcoholic beverages.)

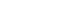

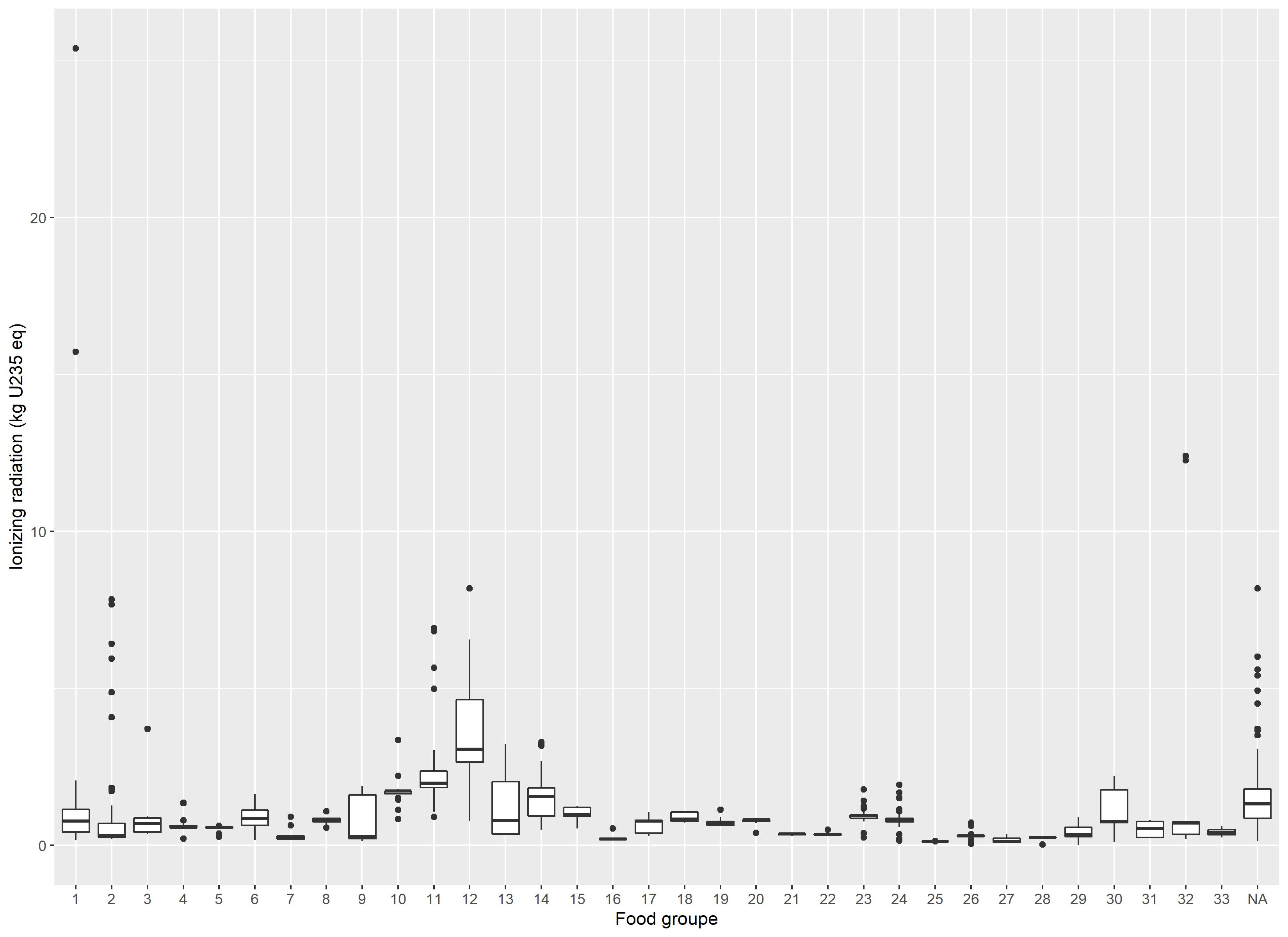

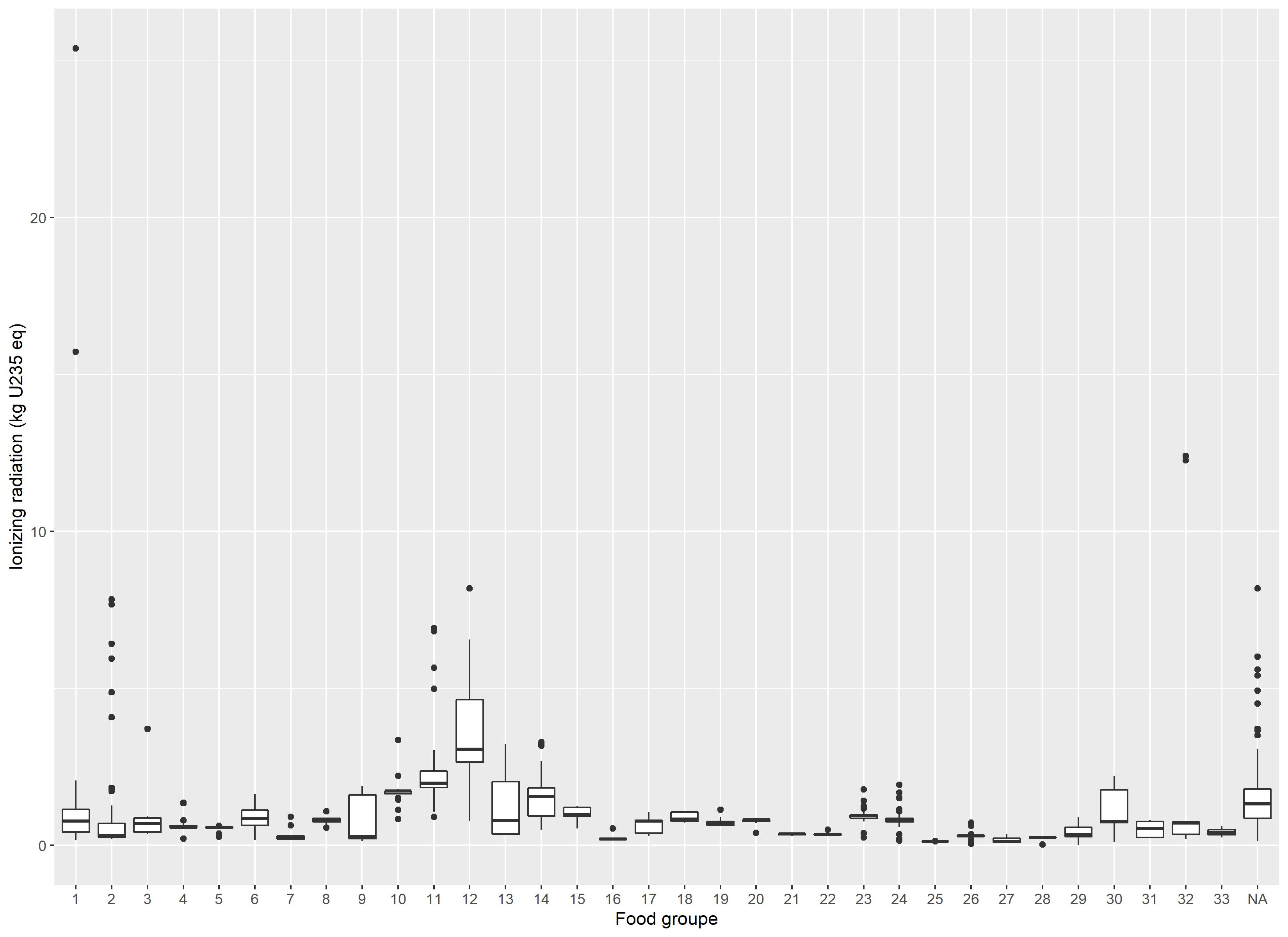

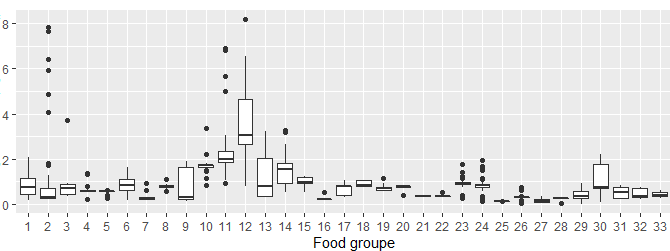

Supplemental Figure 4 Variability of ionizing radiation by food group (1: Vegetable, 2: Fruits, 3: Nuts, seeds and, oleaginous fruits, 4: Refined bread and bread products, 5: Whole and semi-wholemeal bread and bread products, 6: Other refined starchy foods, 7: Other whole and semi-wholemeal starchy foods, 8: Starch products, processed, 9: Legumes, 10: Poultry, 11: Meat excluding poultry, 12: Delicatessen, 13: Fatty fish, 14: Lean fish, 15: Eggs and egg dishes, 16: Milk, 17: Fresh dairy products, 18: Sweet dairy desserts, 19: Cheese, 20: Animal fats, 21: Vegetable fats rich in ALA, 22: Vegetable fats low in ALA, 23: Sauces and fresh creams, 24: Sweet products or sweet and fatty products, 25: Drinking water, 26: Sweet drinks and fruit juices, 27: Hot drinks, 28:Salt, 29: Condiments, 30: Soups and broths, 31: Animal products substitutes, 32: Others foods, 33: Alcoholic beverages.)

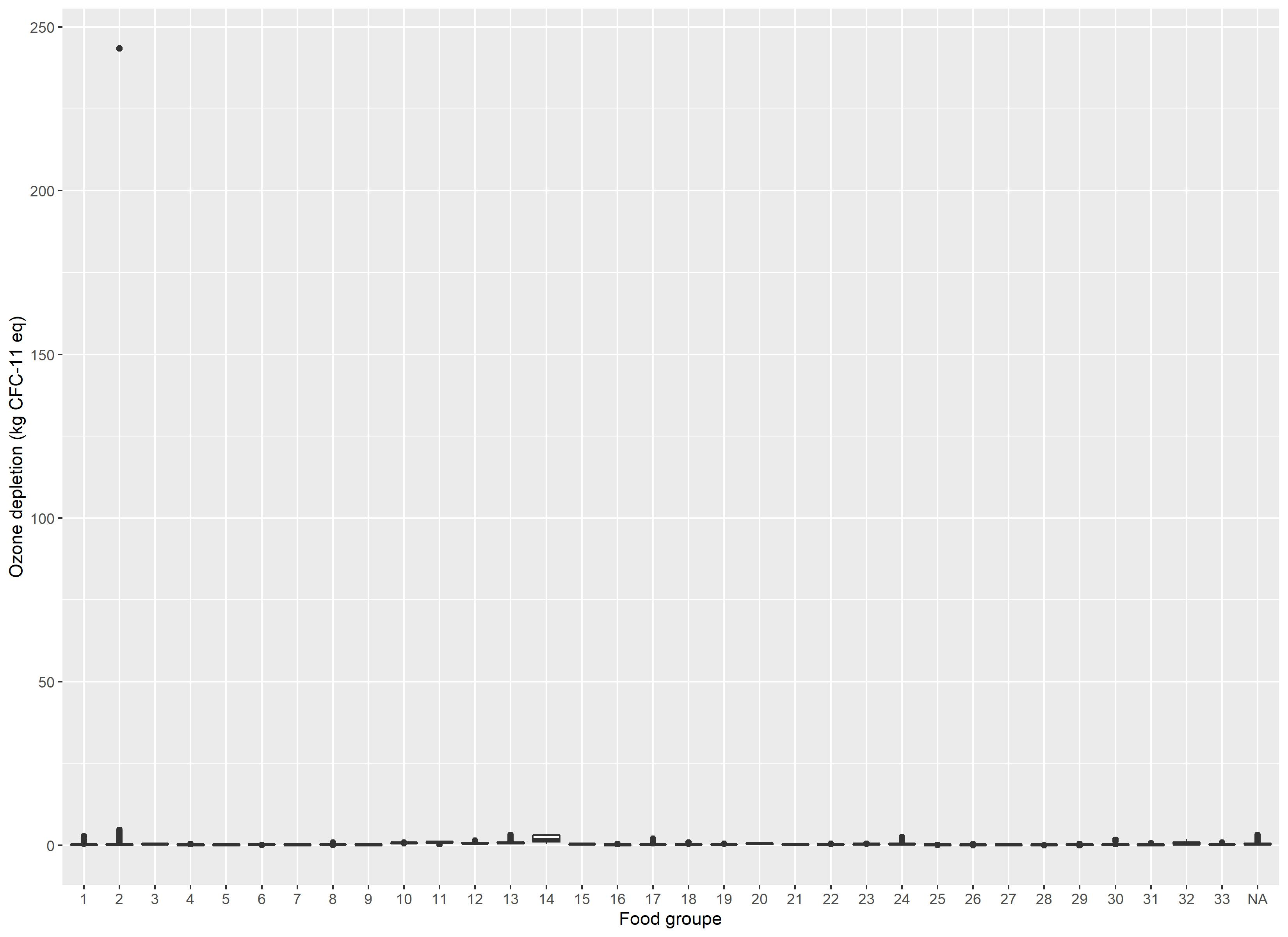

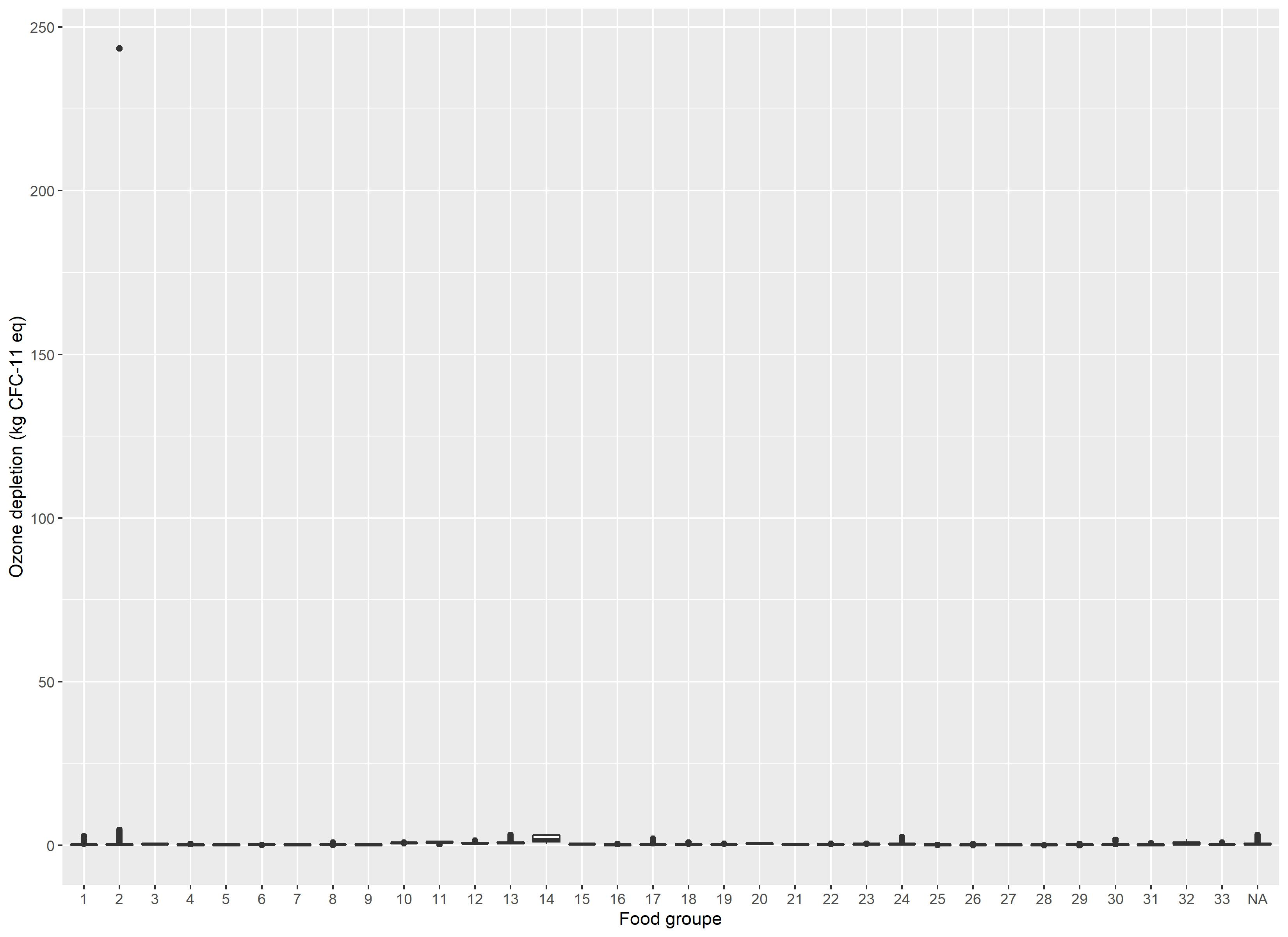

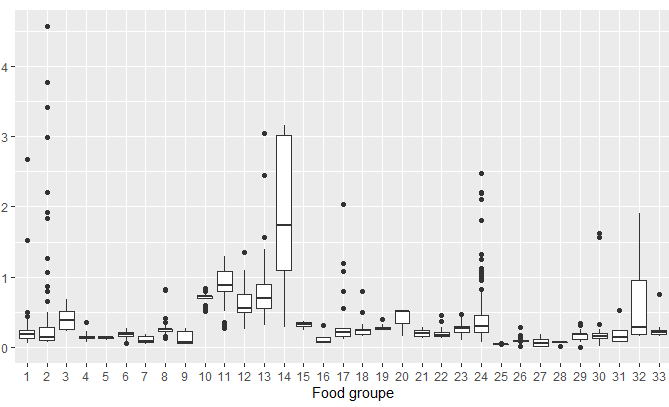

Supplemental Figure 5 Variability ozone depletion by food group (1: Vegetable, 2: Fruits, 3: Nuts, seeds and, oleaginous fruits, 4: Refined bread and bread products, 5: Whole and semi-wholemeal bread and bread products, 6: Other refined starchy foods, 7: Other whole and semi-wholemeal starchy foods, 8: Starch products, processed, 9: Legumes, 10: Poultry, 11: Meat excluding poultry, 12: Delicatessen, 13: Fatty fish, 14: Lean fish, 15: Eggs and egg dishes, 16: Milk, 17: Fresh dairy products, 18: Sweet dairy desserts, 19: Cheese, 20: Animal fats, 21: Vegetable fats rich in ALA, 22: Vegetable fats low in ALA, 23: Sauces and fresh creams, 24: Sweet products or sweet and fatty products, 25: Drinking water, 26: Sweet drinks and fruit juices, 27: Hot drinks, 28:Salt, 29: Condiments, 30: Soups and broths, 31: Animal products substitutes, 32: Others foods, 33: Alcoholic beverages.)

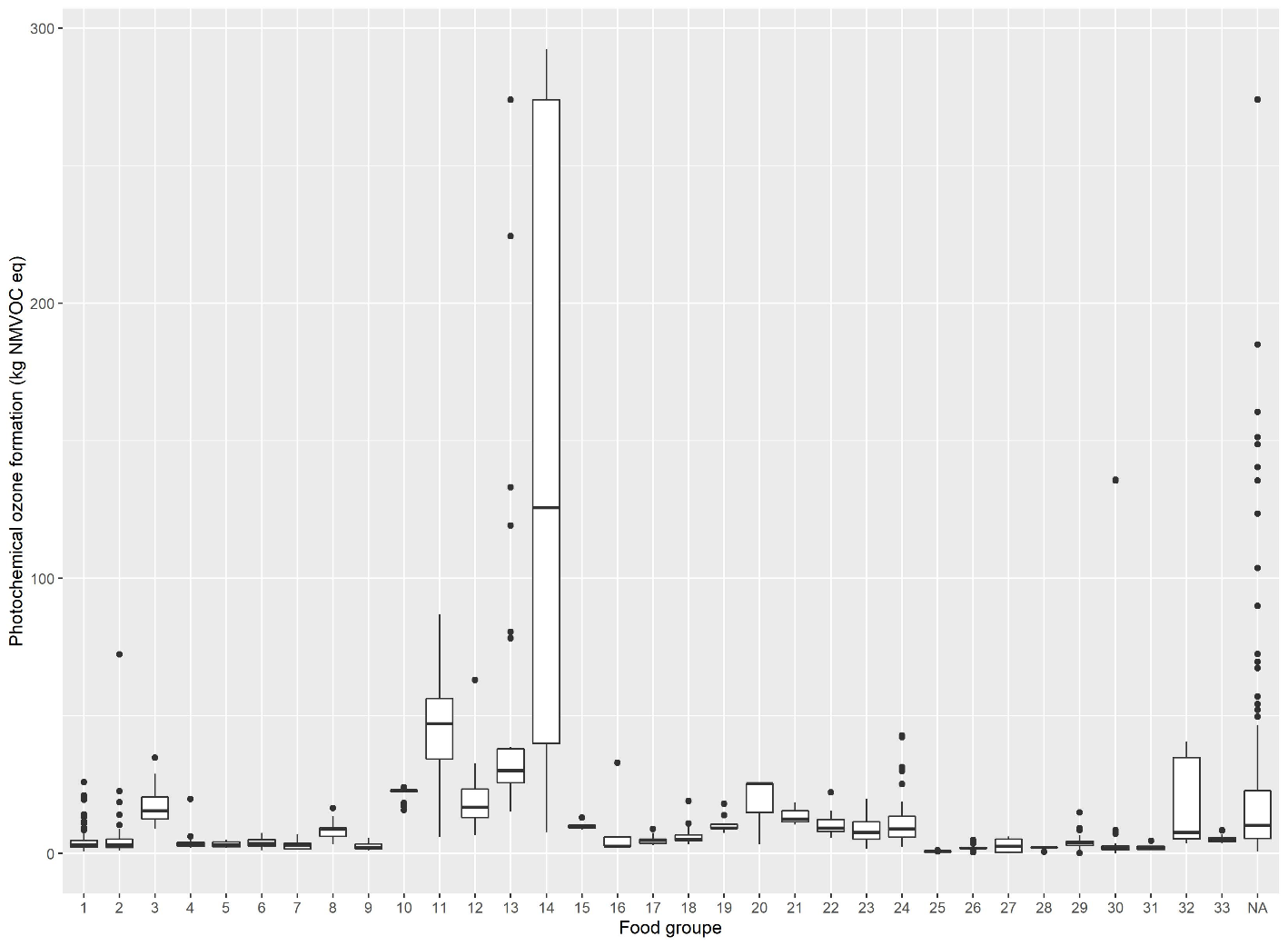

Supplemental Figure 6 Variability of photochemical ozone formation by food group (1: Vegetable, 2: Fruits, 3: Nuts, seeds and, oleaginous fruits, 4: Refined bread and bread products, 5: Whole and semi-wholemeal bread and bread products, 6: Other refined starchy foods, 7: Other whole and semi-wholemeal starchy foods, 8: Starch products, processed, 9: Legumes, 10: Poultry, 11: Meat excluding poultry, 12: Delicatessen, 13: Fatty fish, 14: Lean fish, 15: Eggs and egg dishes, 16: Milk, 17: Fresh dairy products, 18: Sweet dairy desserts, 19: Cheese, 20: Animal fats, 21: Vegetable fats rich in ALA, 22: Vegetable fats low in ALA, 23: Sauces and fresh creams, 24: Sweet products or sweet and fatty products, 25: Drinking water, 26: Sweet drinks and fruit juices, 27: Hot drinks, 28:Salt, 29: Condiments, 30: Soups and broths, 31: Animal products substitutes, 32: Others foods, 33: Alcoholic beverages.)

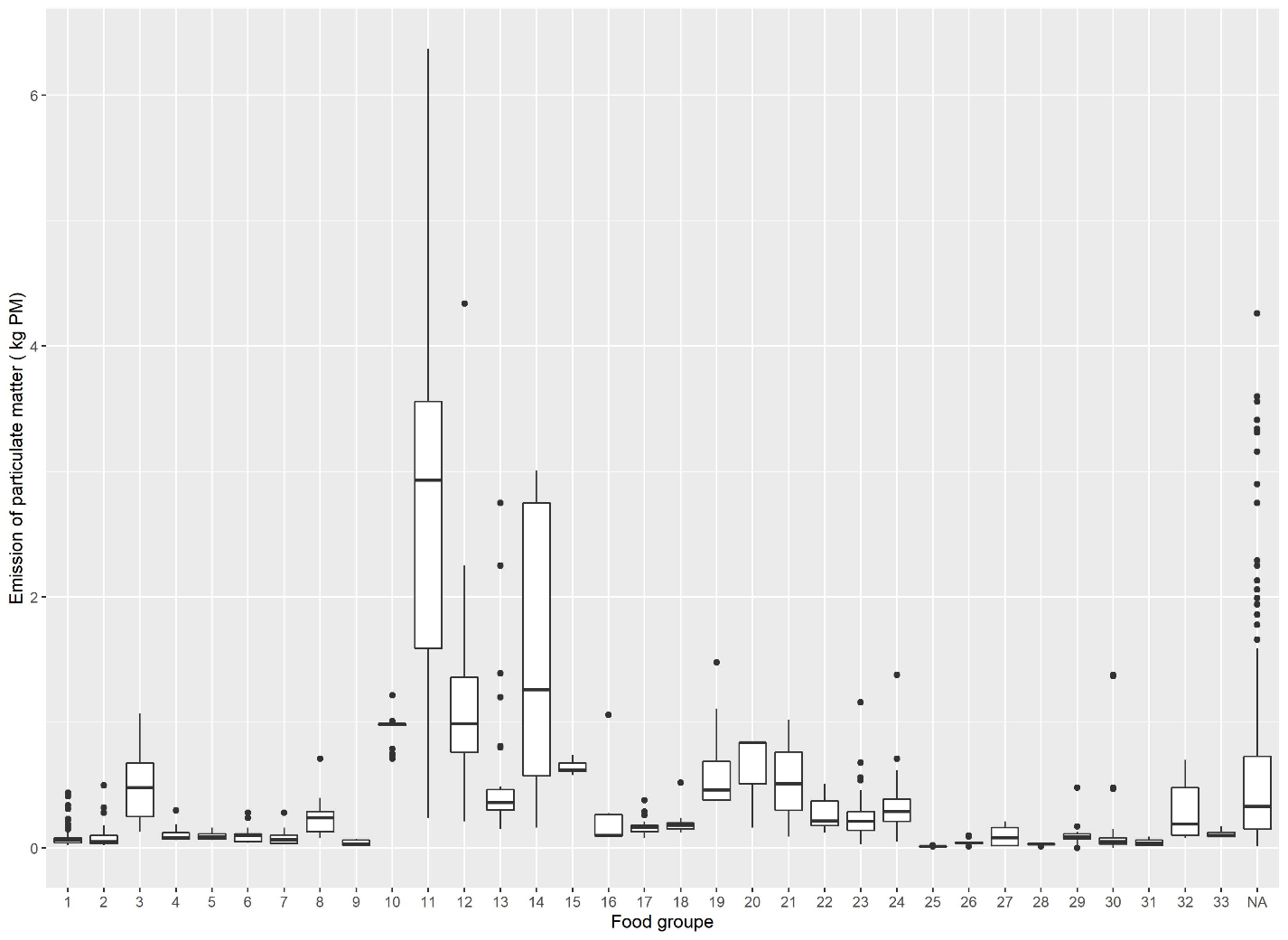

Supplemental Figure 7 Variability of the emission of particulate matter by food group (1: Vegetable, 2: Fruits, 3: Nuts, seeds and, oleaginous fruits, 4: Refined bread and bread products, 5: Whole and semi-wholemeal bread and bread products, 6: Other refined starchy foods, 7: Other whole and semi-wholemeal starchy foods, 8: Starch products, processed, 9: Legumes, 10: Poultry, 11: Meat excluding poultry, 12: Delicatessen, 13: Fatty fish, 14: Lean fish, 15: Eggs and egg dishes, 16: Milk, 17: Fresh dairy products, 18: Sweet dairy desserts, 19: Cheese, 20: Animal fats, 21: Vegetable fats rich in ALA, 22: Vegetable fats low in ALA, 23: Sauces and fresh creams, 24: Sweet products or sweet and fatty products, 25: Drinking water, 26: Sweet drinks and fruit juices, 27: Hot drinks, 28:Salt, 29: Condiments, 30: Soups and broths, 31: Animal products substitutes, 32: Others foods, 33: Alcoholic beverages.)

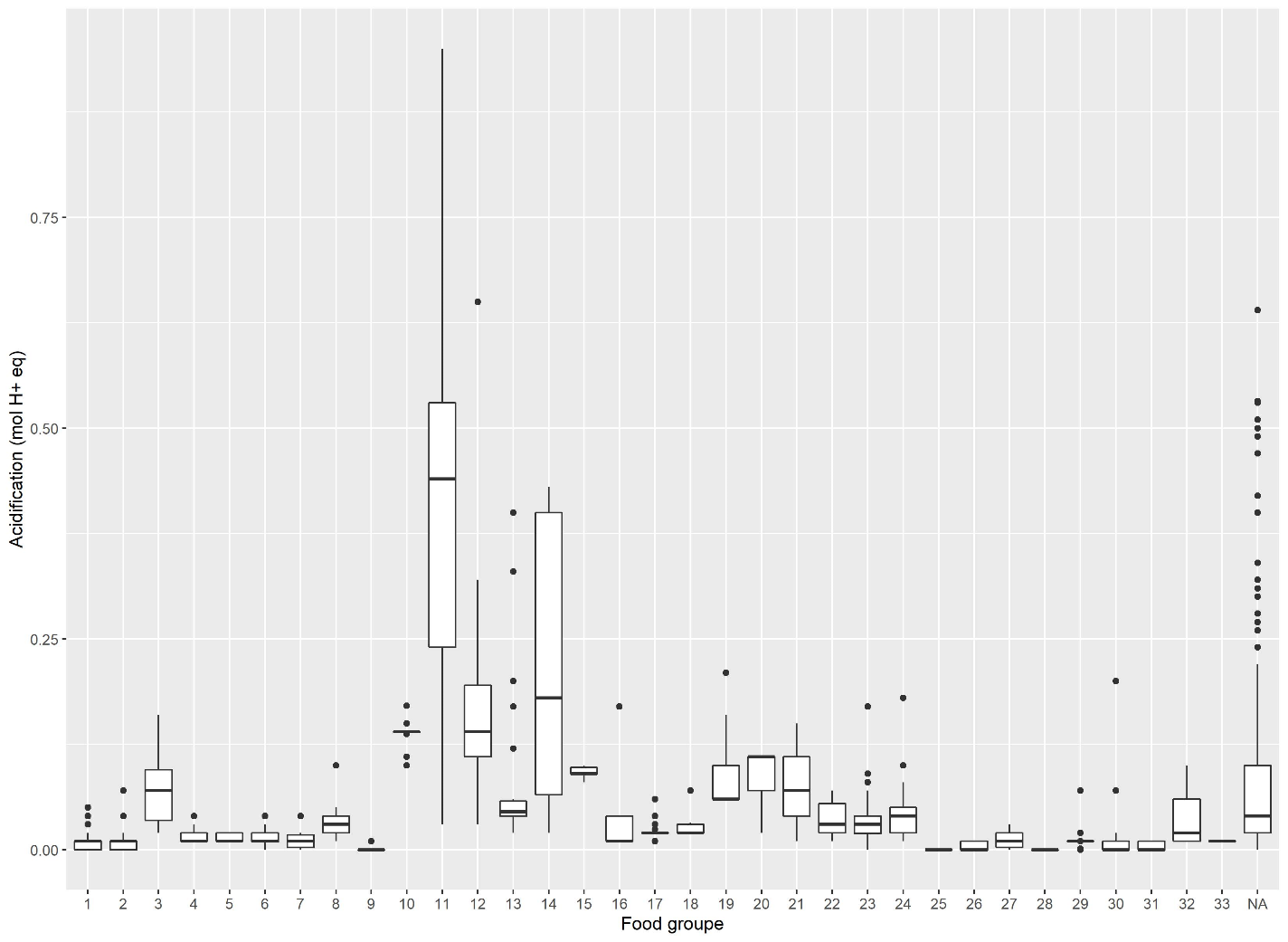

Supplemental Figure 8 Variability of the acidification by food group (1: Vegetable, 2: Fruits, 3: Nuts, seeds and, oleaginous fruits, 4: Refined bread and bread products, 5: Whole and semi-wholemeal bread and bread products, 6: Other refined starchy foods, 7: Other whole and semi-wholemeal starchy foods, 8: Starch products, processed, 9: Legumes, 10: Poultry, 11: Meat excluding poultry, 12: Delicatessen, 13: Fatty fish, 14: Lean fish, 15: Eggs and egg dishes, 16: Milk, 17: Fresh dairy products, 18: Sweet dairy desserts, 19: Cheese, 20: Animal fats, 21: Vegetable fats rich in ALA, 22: Vegetable fats low in ALA, 23: Sauces and fresh creams, 24: Sweet products or sweet and fatty products, 25: Drinking water, 26: Sweet drinks and fruit juices, 27: Hot drinks, 28:Salt, 29: Condiments, 30: Soups and broths, 31: Animal products substitutes, 32: Others foods, 33: Alcoholic beverages.)

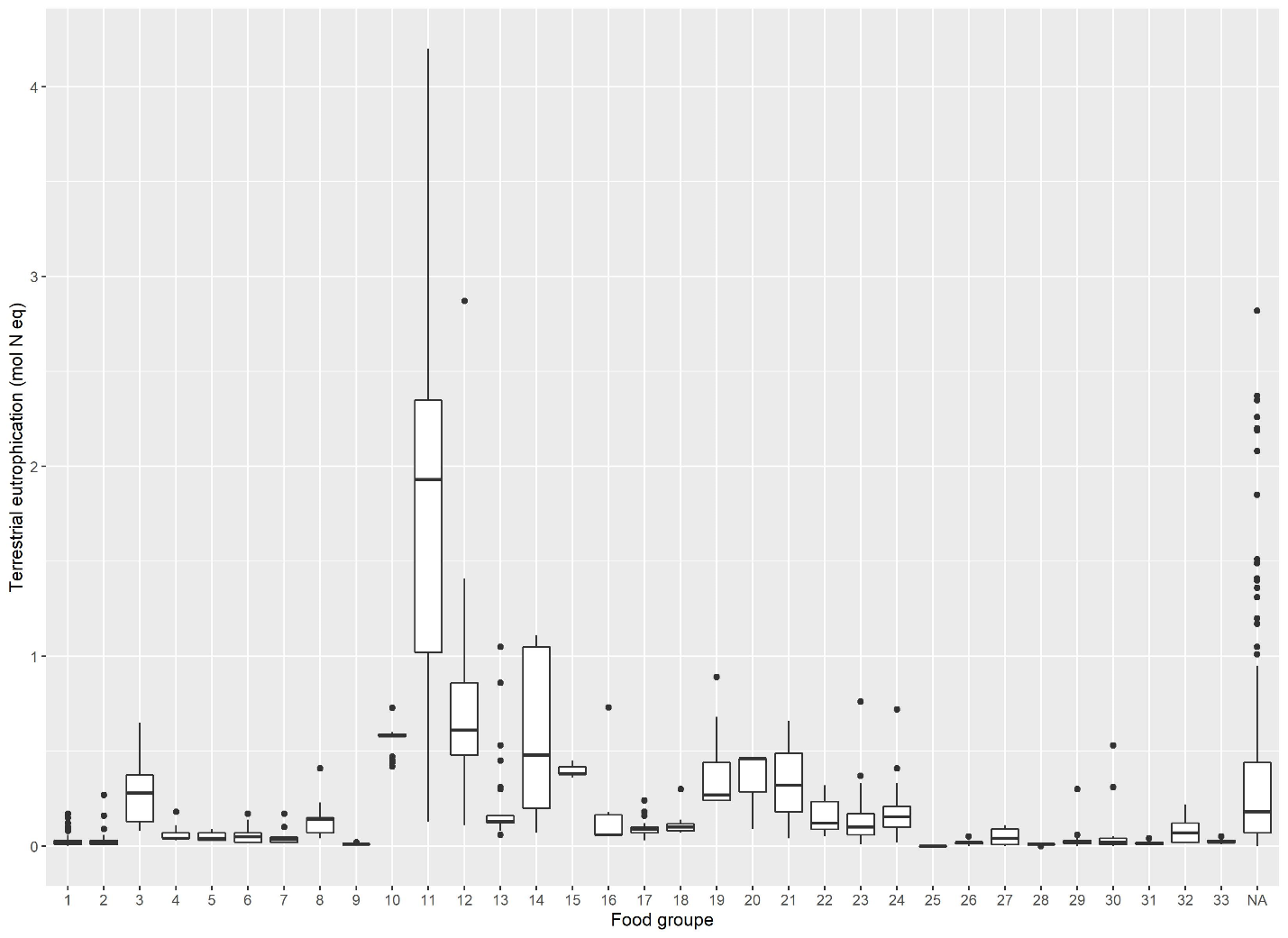

Supplemental Figure 9 Variability of terrestrial eutrophication by food group (1: Vegetable, 2: Fruits, 3: Nuts, seeds and, oleaginous fruits, 4: Refined bread and bread products, 5: Whole and semi-wholemeal bread and bread products, 6: Other refined starchy foods, 7: Other whole and semi-wholemeal starchy foods, 8: Starch products, processed, 9: Legumes, 10: Poultry, 11: Meat excluding poultry, 12: Delicatessen, 13: Fatty fish, 14: Lean fish, 15: Eggs and egg dishes, 16: Milk, 17: Fresh dairy products, 18: Sweet dairy desserts, 19: Cheese, 20: Animal fats, 21: Vegetable fats rich in ALA, 22: Vegetable fats low in ALA, 23: Sauces and fresh creams, 24: Sweet products or sweet and fatty products, 25: Drinking water, 26: Sweet drinks and fruit juices, 27: Hot drinks, 28:Salt, 29: Condiments, 30: Soups and broths, 31: Animal products substitutes, 32: Others foods, 33: Alcoholic beverages.)

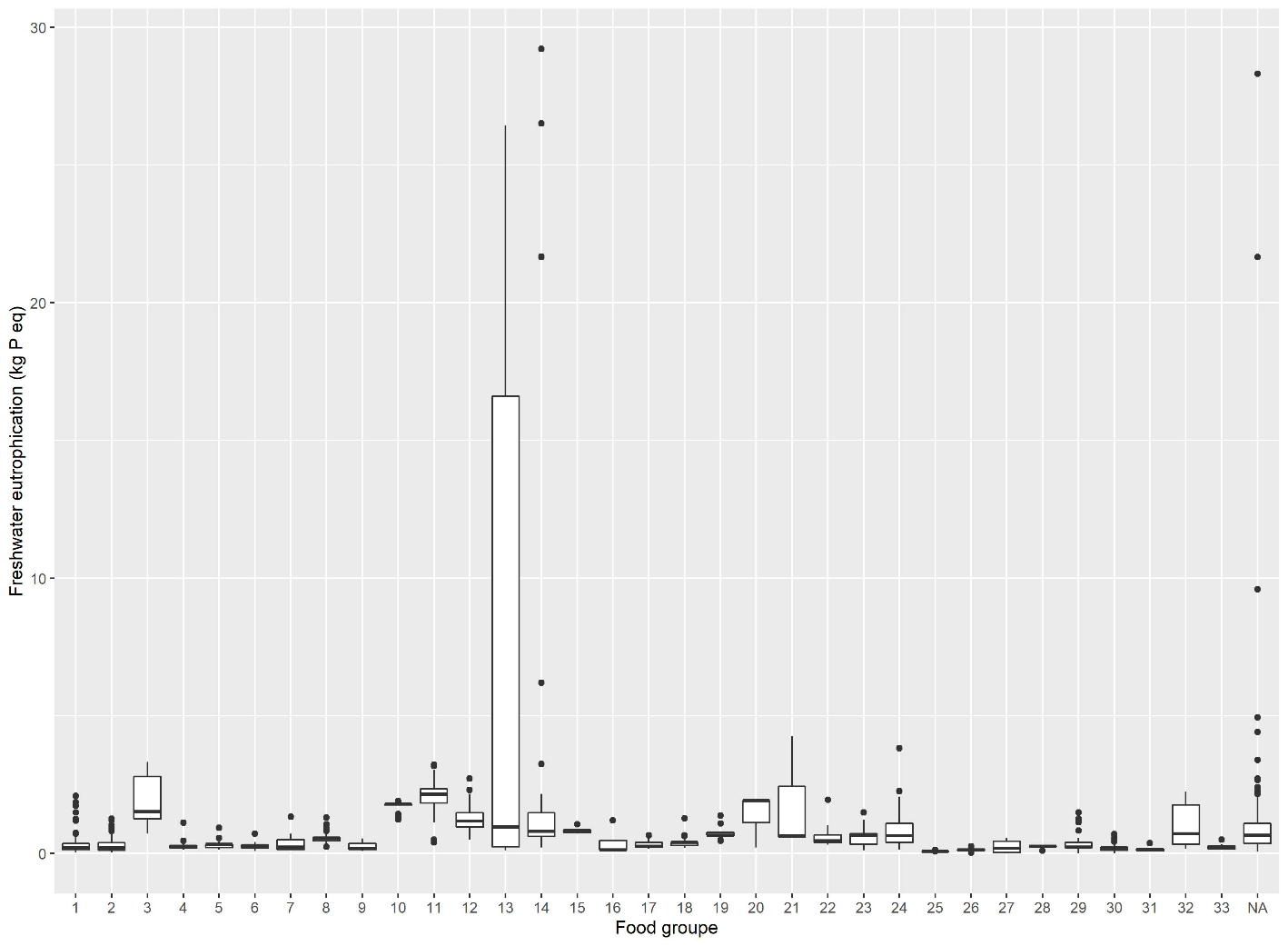

Supplemental Figure 10 Variability of freshwater eutrophication by food group (1: Vegetable, 2: Fruits, 3: Nuts, seeds and, oleaginous fruits, 4: Refined bread and bread products, 5: Whole and semi-wholemeal bread and bread products, 6: Other refined starchy foods, 7: Other whole and semi-wholemeal starchy foods, 8: Starch products, processed, 9: Legumes, 10: Poultry, 11: Meat excluding poultry, 12: Delicatessen, 13: Fatty fish, 14: Lean fish, 15: Eggs and egg dishes, 16: Milk, 17: Fresh dairy products, 18: Sweet dairy desserts, 19: Cheese, 20: Animal fats, 21: Vegetable fats rich in ALA, 22: Vegetable fats low in ALA, 23: Sauces and fresh creams, 24: Sweet products or sweet and fatty products, 25: Drinking water, 26: Sweet drinks and fruit juices, 27: Hot drinks, 28:Salt, 29: Condiments, 30: Soups and broths, 31: Animal products substitutes, 32: Others foods, 33: Alcoholic beverages.)

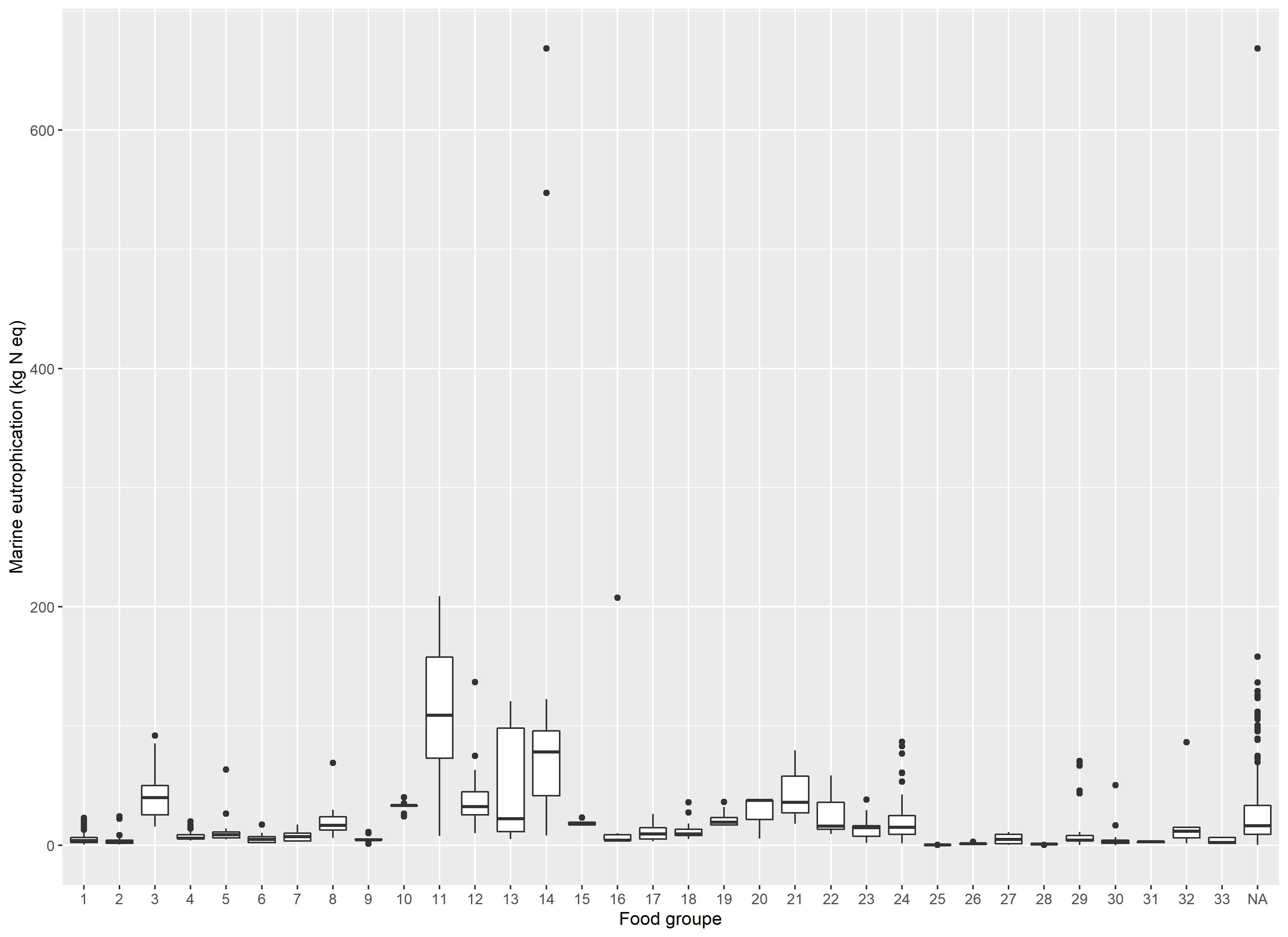

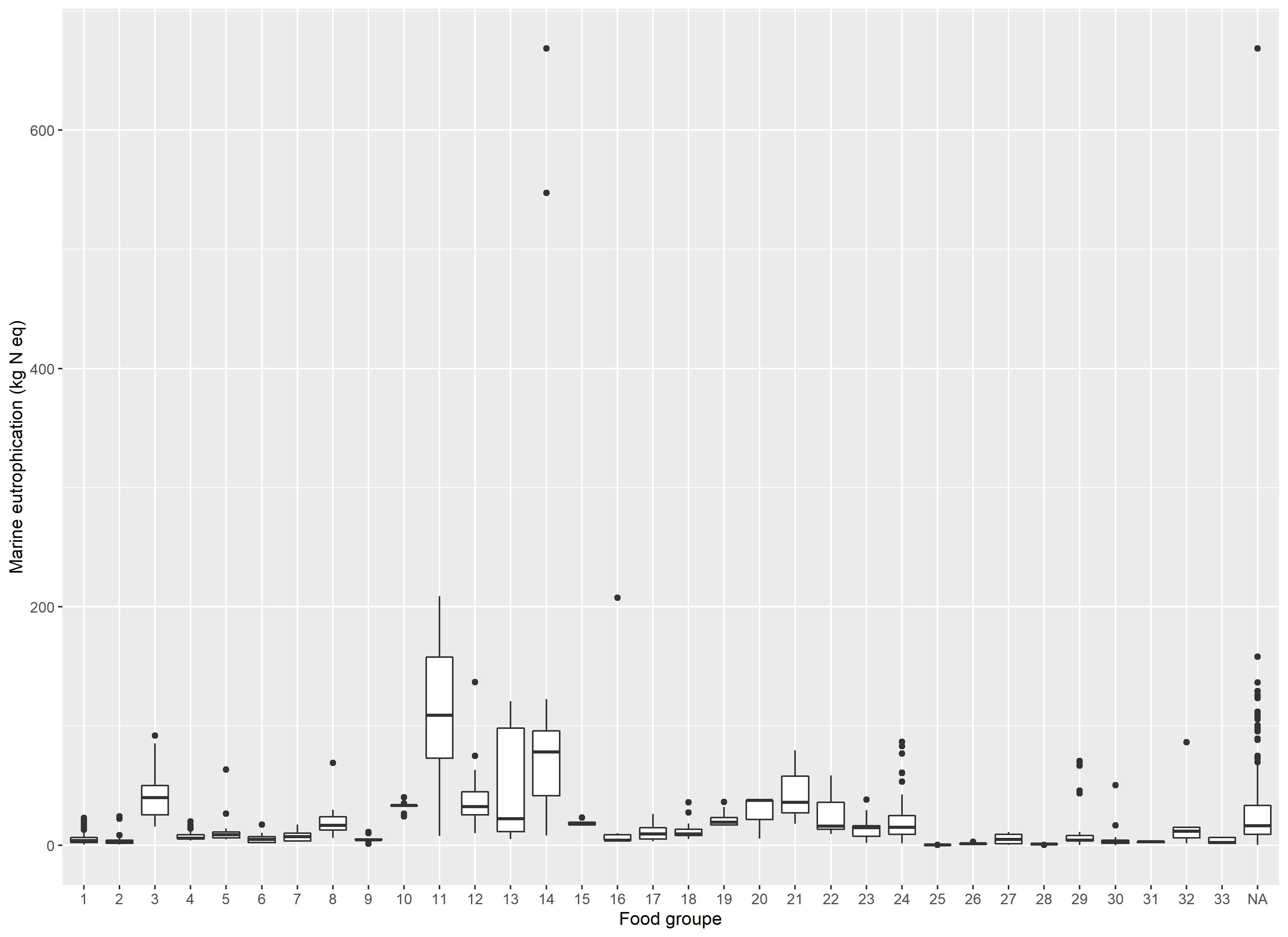

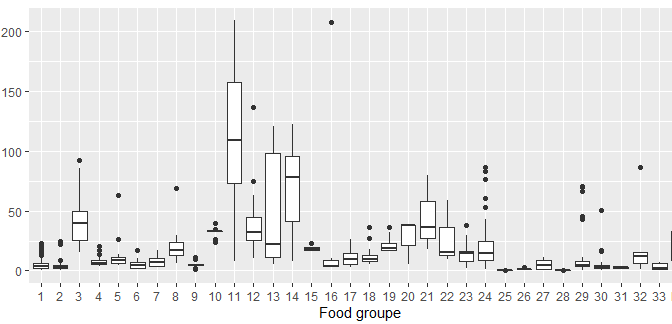

Supplemental Figure 11 Variability of marine eutrophication by food group (1: Vegetable, 2: Fruits, 3: Nuts, seeds and, oleaginous fruits, 4: Refined bread and bread products, 5: Whole and semi-wholemeal bread and bread products, 6: Other refined starchy foods, 7: Other whole and semi-wholemeal starchy foods, 8: Starch products, processed, 9: Legumes, 10: Poultry, 11: Meat excluding poultry, 12: Delicatessen, 13: Fatty fish, 14: Lean fish, 15: Eggs and egg dishes, 16: Milk, 17: Fresh dairy products, 18: Sweet dairy desserts, 19: Cheese, 20: Animal fats, 21: Vegetable fats rich in ALA, 22: Vegetable fats low in ALA, 23: Sauces and fresh creams, 24: Sweet products or sweet and fatty products, 25: Drinking water, 26: Sweet drinks and fruit juices, 27: Hot drinks, 28:Salt, 29: Condiments, 30: Soups and broths, 31: Animal products substitutes, 32: Others foods, 33: Alcoholic beverages.)

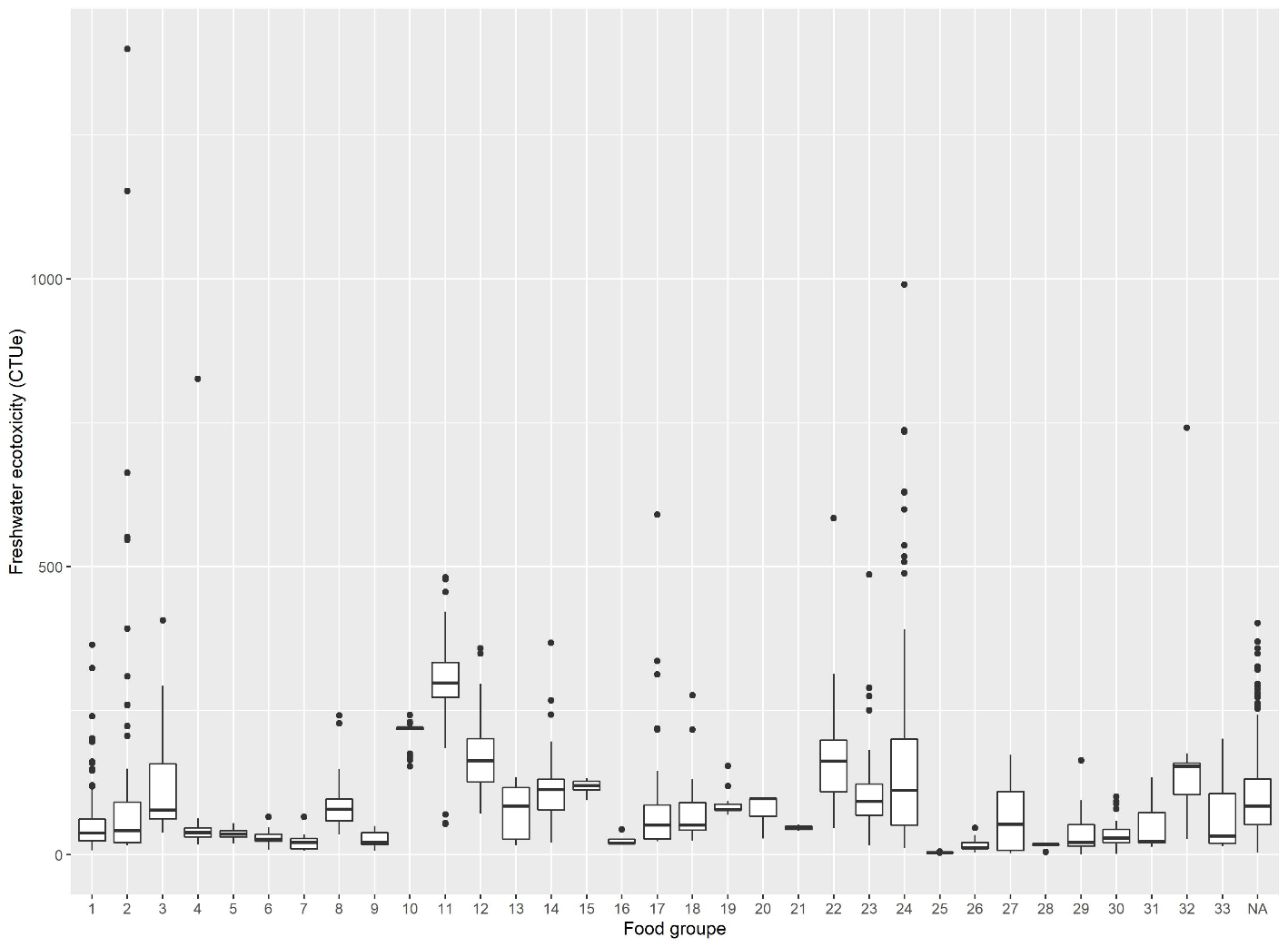

Supplemental Figure 12 Variability of freshwater ecotoxicity by food group (1: Vegetable, 2: Fruits, 3: Nuts, seeds and, oleaginous fruits, 4: Refined bread and bread products, 5: Whole and semi-wholemeal bread and bread products, 6: Other refined starchy foods, 7: Other whole and semi-wholemeal starchy foods, 8: Starch products, processed, 9: Legumes, 10: Poultry, 11: Meat excluding poultry, 12: Delicatessen, 13: Fatty fish, 14: Lean fish, 15: Eggs and egg dishes, 16: Milk, 17: Fresh dairy products, 18: Sweet dairy desserts, 19: Cheese, 20: Animal fats, 21: Vegetable fats rich in ALA, 22: Vegetable fats low in ALA, 23: Sauces and fresh creams, 24: Sweet products or sweet and fatty products, 25: Drinking water, 26: Sweet drinks and fruit juices, 27: Hot drinks, 28:Salt, 29: Condiments, 30: Soups and broths, 31: Animal products substitutes, 32: Others foods, 33: Alcoholic beverages.)

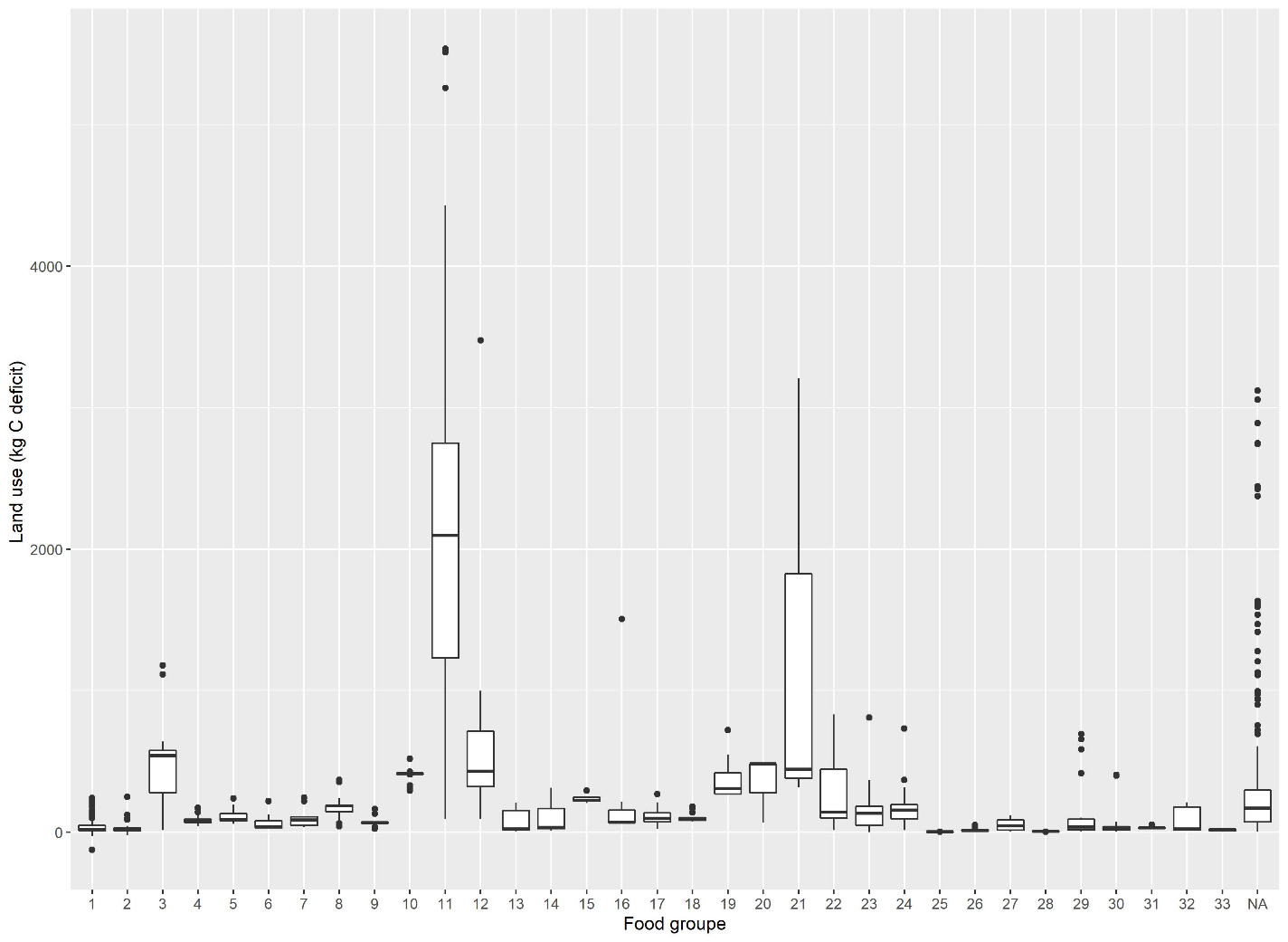

Supplemental Figure 13 Variability of land use by food group (1: Vegetable, 2: Fruits, 3: Nuts, seeds and, oleaginous fruits, 4: Refined bread and bread products, 5: Whole and semi-wholemeal bread and bread products, 6: Other refined starchy foods, 7: Other whole and semi-wholemeal starchy foods, 8: Starch products, processed, 9: Legumes, 10: Poultry, 11: Meat excluding poultry, 12: Delicatessen, 13: Fatty fish, 14: Lean fish, 15: Eggs and egg dishes, 16: Milk, 17: Fresh dairy products, 18: Sweet dairy desserts, 19: Cheese, 20: Animal fats, 21: Vegetable fats rich in ALA, 22: Vegetable fats low in ALA, 23: Sauces and fresh creams, 24: Sweet products or sweet and fatty products, 25: Drinking water, 26: Sweet drinks and fruit juices, 27: Hot drinks, 28:Salt, 29: Condiments, 30: Soups and broths, 31: Animal products substitutes, 32: Others foods, 33: Alcoholic beverages.)

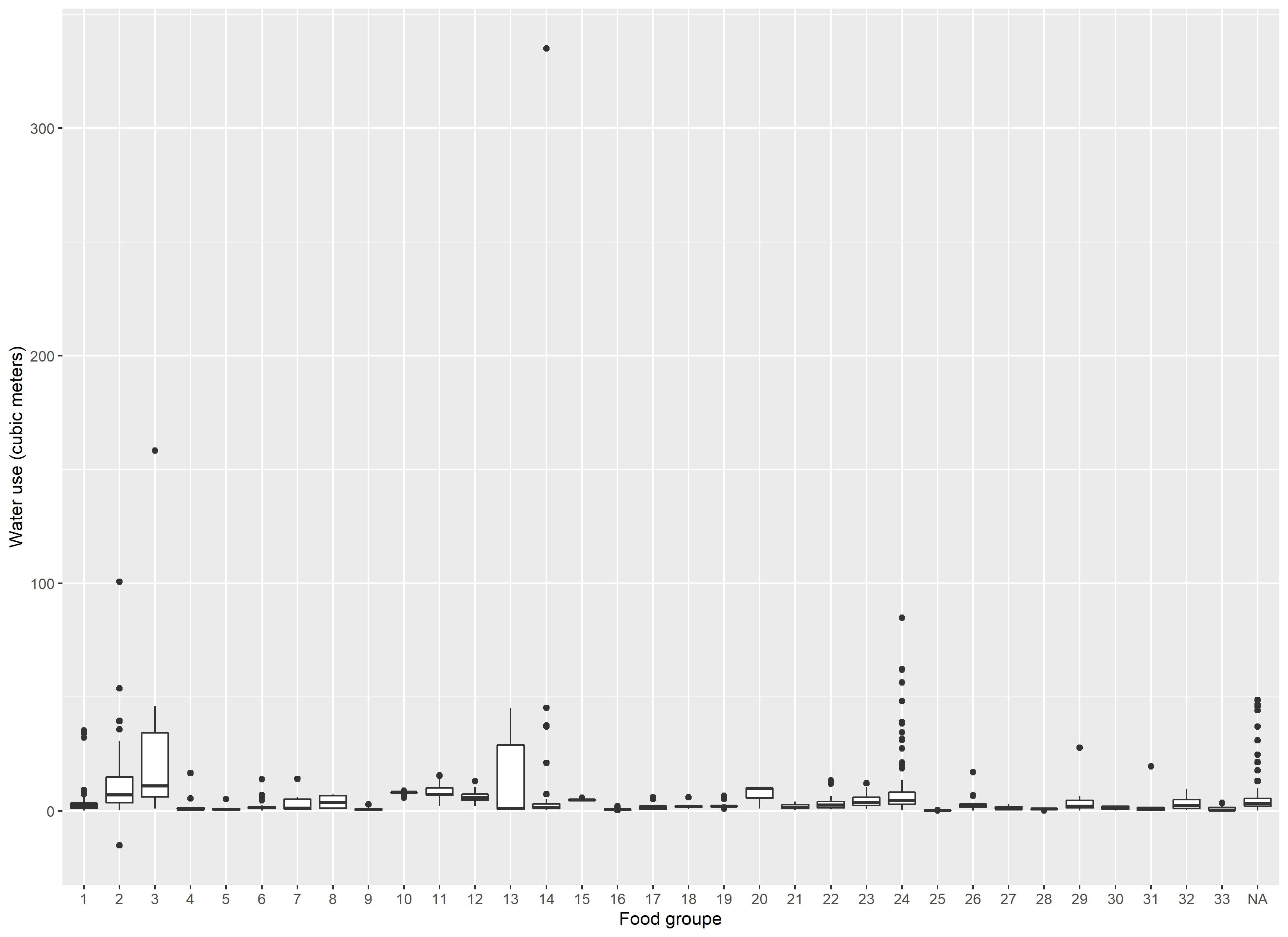

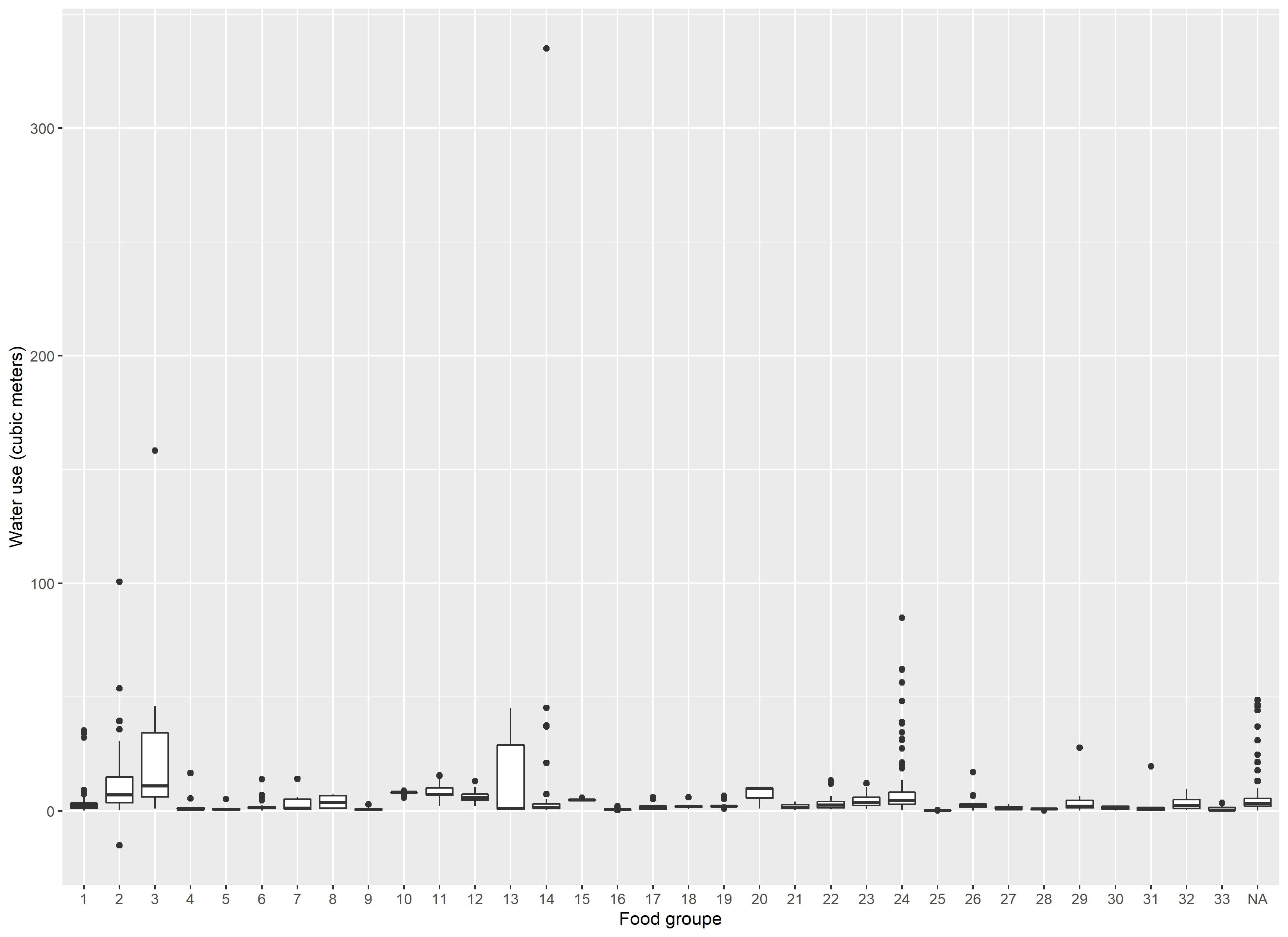

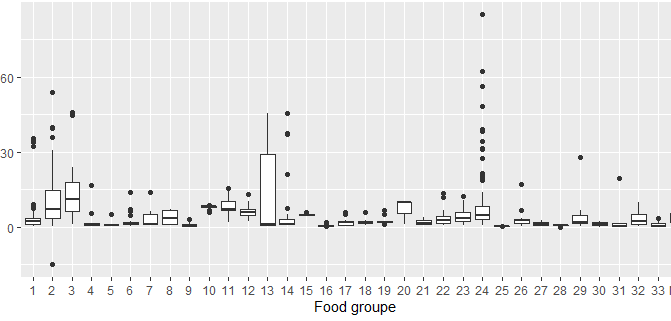

Supplemental Figure 14 Variability of water use by food group (1: Vegetable, 2: Fruits, 3: Nuts, seeds and, oleaginous fruits, 4: Refined bread and bread products, 5: Whole and semi-wholemeal bread and bread products, 6: Other refined starchy foods, 7: Other whole and semi-wholemeal starchy foods, 8: Starch products, processed, 9: Legumes, 10: Poultry, 11: Meat excluding poultry, 12: Delicatessen, 13: Fatty fish, 14: Lean fish, 15: Eggs and egg dishes, 16: Milk, 17: Fresh dairy products, 18: Sweet dairy desserts, 19: Cheese, 20: Animal fats, 21: Vegetable fats rich in ALA, 22: Vegetable fats low in ALA, 23: Sauces and fresh creams, 24: Sweet products or sweet and fatty products, 25: Drinking water, 26: Sweet drinks and fruit juices, 27: Hot drinks, 28:Salt, 29: Condiments, 30: Soups and broths, 31: Animal products substitutes, 32: Others foods, 33: Alcoholic beverages.)

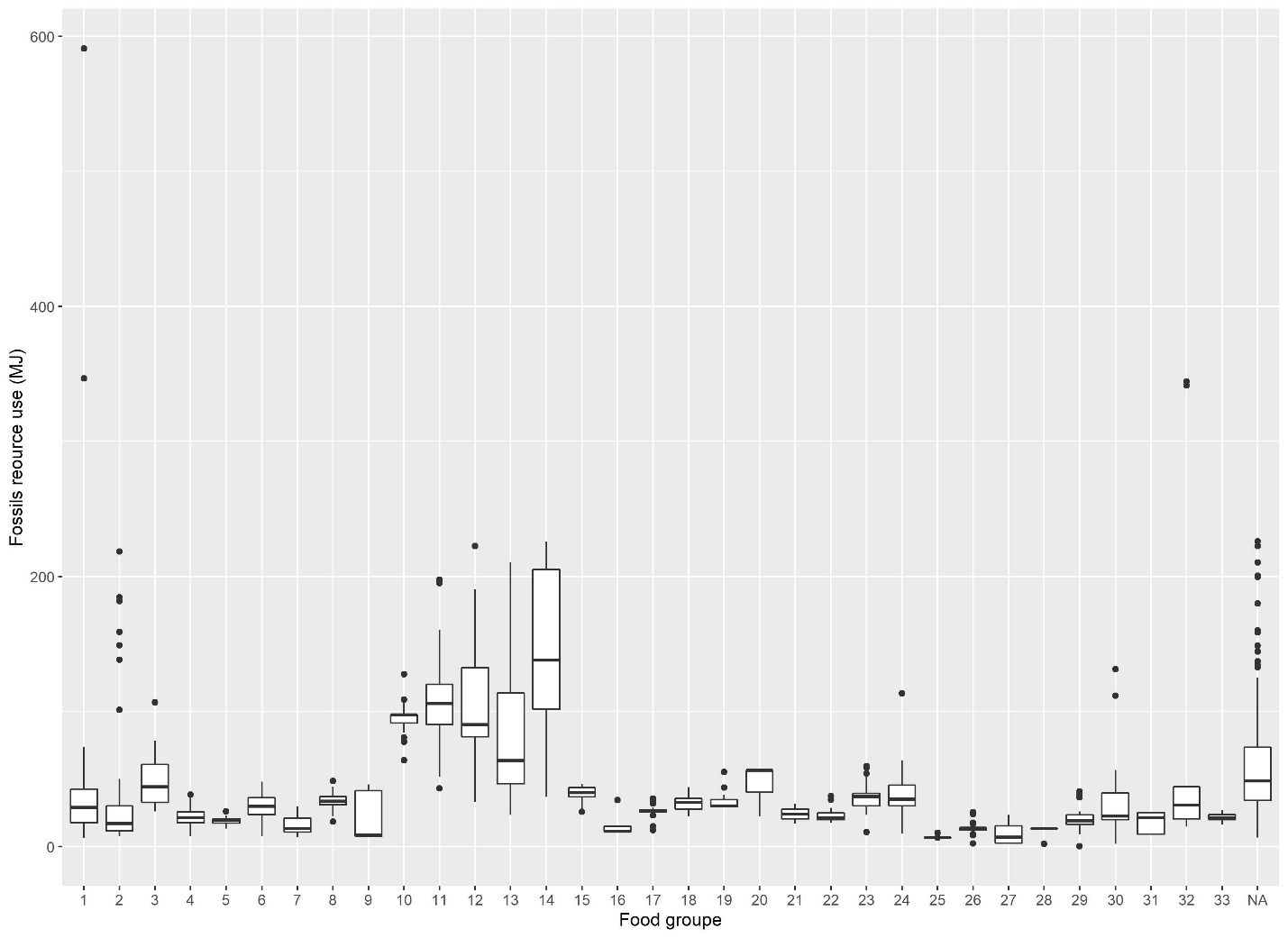

Supplemental Figure 15 Variability of fossils resource use by food group (1: Vegetable, 2: Fruits, 3: Nuts, seeds and, oleaginous fruits, 4: Refined bread and bread products, 5: Whole and semi-wholemeal bread and bread products, 6: Other refined starchy foods, 7: Other whole and semi-wholemeal starchy foods, 8: Starch products, processed, 9: Legumes, 10: Poultry, 11: Meat excluding poultry, 12: Delicatessen, 13: Fatty fish, 14: Lean fish, 15: Eggs and egg dishes, 16: Milk, 17: Fresh dairy products, 18: Sweet dairy desserts, 19: Cheese, 20: Animal fats, 21: Vegetable fats rich in ALA, 22: Vegetable fats low in ALA, 23: Sauces and fresh creams, 24: Sweet products or sweet and fatty products, 25: Drinking water, 26: Sweet drinks and fruit juices, 27: Hot drinks, 28:Salt, 29: Condiments, 30: Soups and broths, 31: Animal products substitutes, 32: Others foods, 33: Alcoholic beverages.)

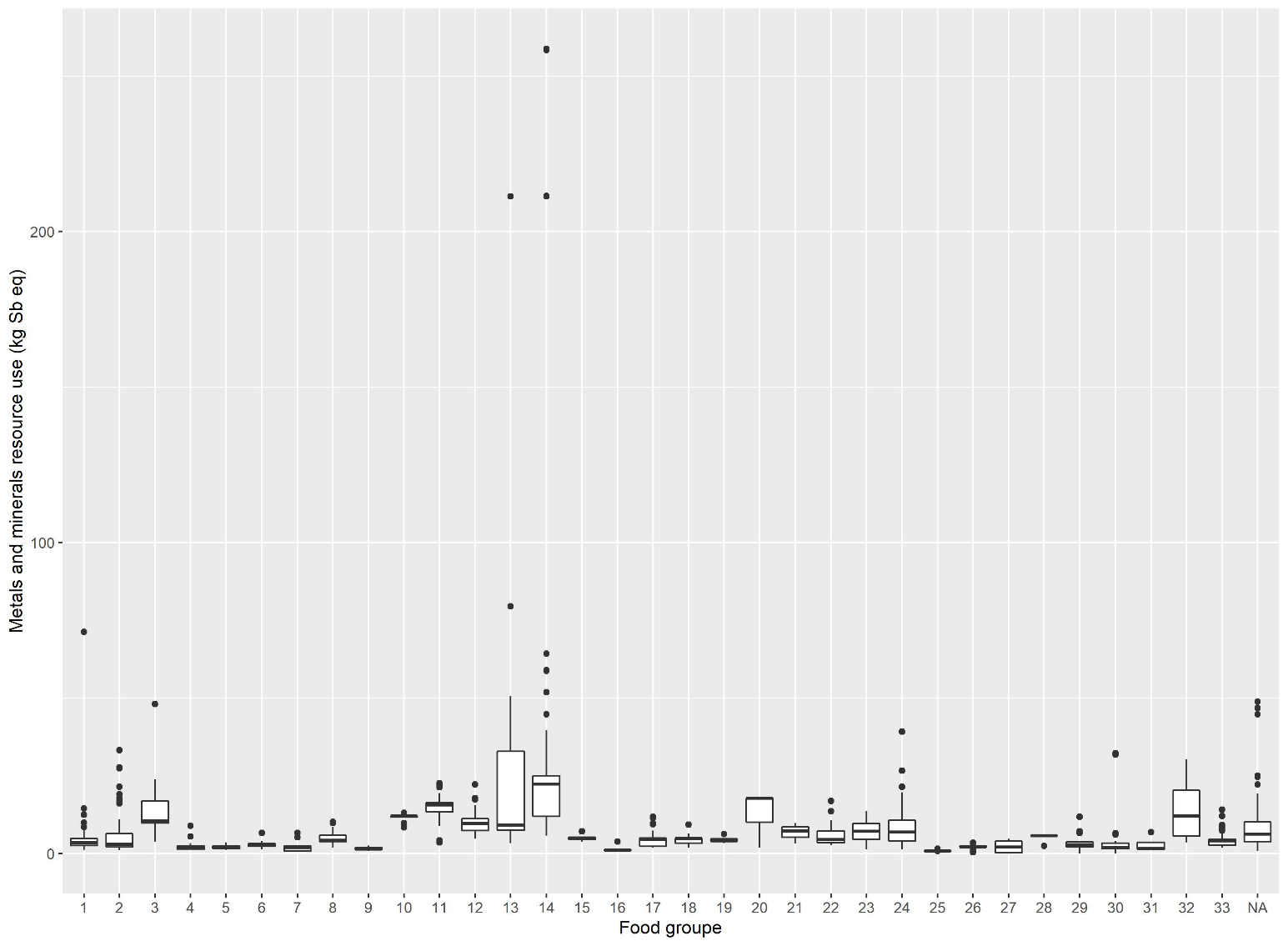

Supplemental Figure 16 Variability of metals and minerals resource use by food group (1: Vegetable, 2: Fruits, 3: Nuts, seeds and, oleaginous fruits, 4: Refined bread and bread products, 5: Whole and semi-wholemeal bread and bread products, 6: Other refined starchy foods, 7: Other whole and semi-wholemeal starchy foods, 8: Starch products, processed, 9: Legumes, 10: Poultry, 11: Meat excluding poultry, 12: Delicatessen, 13: Fatty fish, 14: Lean fish, 15: Eggs and egg dishes, 16: Milk, 17: Fresh dairy products, 18: Sweet dairy desserts, 19: Cheese, 20: Animal fats, 21: Vegetable fats rich in ALA, 22: Vegetable fats low in ALA, 23: Sauces and fresh creams, 24: Sweet products or sweet and fatty products, 25: Drinking water, 26: Sweet drinks and fruit juices, 27: Hot drinks, 28:Salt, 29: Condiments, 30: Soups and broths, 31: Animal products substitutes, 32: Others foods, 33: Alcoholic beverages.)

**Supplemental Method 1 Agribalyse database**

The environmental pressures associated with diets were estimated by matching INCA3 consumption data with the French database Agribalyse® 3.0.1 developed by the French Agency for the Environment and Energy Management (ADEME). This allowed for a connection to be made between the environmental and nutritional properties of foods for 2497 foods consumed in France (1).

Environmental indicator estimations are based on the method of Life Cycle Assessment (LCA), whose scope is "from field to plate." The perimeter of the indicators covers each process of the value chain: production of raw materials, transport, transformation, packaging, distribution and retailing, food preparation by the consumer, and disposal of packaging. These processes are split into two phases 1) production and 2) post-farm. Of note for Agribalyse, losses and wastes (other than non-edible parts) at home, as well as transport from the retail to the household are not considered.

Overall, Agribalyse construction is based on the international LCA standards: ISO 14040 (2) and ISO 14044(3), LEAP guidelines (4), and PEF (5). The finalized indicators are provided per kg of product and are detailed per process.

For the agricultural phase of plant products, all upstream processes (notably input production), excluding storage or drying, are included except for ingredients in processed food. In the case of animal products, all operations including the phases of production, transport and storage of feed, fattening of animals, milking, construction and maintenance of buildings and machinery, have been considered. The scope chosen is consistent with those defined in GESTIM (6) and ecoinvent® (7).

The LCI (life cycle inventory) data of AGRIBALYSE v3.0.1 covered the period 2005-2009, except for perennial crops (2000-2010). The variety of production systems was considered by applying coefficients based on the share of systems in national production. The allocation rules are varied and are based on international recommendations as described by the ISO 14040/14044 standards (2,3). In particular, allocations have been developed in order to distribute organic nitrogen fertilizers and mineral fertilizers (P and K) between crop sequences. Biophysical allocations were used for animal production (milk versus meat). The biophysical models used for animal production and allocations by type of productions are presented in the full report (8) according to the reference AFNOR-BPX 30-323 (9)(AFNOR, 2011) and in compliance with the ISO 14044 standard (3) according to three rules in descending order: 1) allocation avoidance, 2) : biophysic allocation and 3) economic allocation.

A characterization method recommended by the European Commission (Environmental Footprint 3.0) translates the input and output flows of the inventory into impacts. For the background data (inputs in construction, raw materials, etc.) the ecoinvent® database is used to assess the indirect emissions (off-field emissions). The full methodology and methodological choices for the use of the ecoinvent® database have been described elsewhere (8).

The transition from commodities to consumed food results in two coefficients related to the edible food part, and economic allocations between co-products. The recipes are then disaggregated into ingredients. For feasibility reasons, a threshold of 95% of the ingredients covered was used. Similarly, for the origin of the ingredients, a threshold of 70% coverage was used followed by a standardization step. See (10) for the entire description of the methodology and methodological adoptions. Post-farm estimations are based on the PEF guidelines (5)

A total of 14 midpoint indicators are available in Agribalyse: climate change, ozone depletion, particulate matters, ionizing radiation (effect on human health), ecotoxicity, photochemical ozone formation (effect on human health), acidification, terrestrial eutrophication, freshwater eutrophication, marine eutrophication, land use, water use, minerals and metals use, and fossil resources use.

Description of the environmental indicators

| **Environmental indicator** | **Description** | **Unit** |
| --- | --- | --- |
| Greenhouse gas emission | Linked to the increase in the average global temperatures | Carbon dioxide equivalent (kg CO2 eq) |
| Exposure ionizing radiation | The impact on human health due to emissions of radiation under normal operating conditions | Equivalent of kilobecquerels of Uranium 235 (kg U235 eq) |
| Photochemical ozone (O3) formation | Ozone on the ground has a deleterious impact on organic compounds including animals and plants. It increases for example the frequency of respiratory problems for humans | Equivalent of kilograms of non-methane volatile organic compounds (kg NMVOC eq) |
| Ozone depletion | Stratospheric ozone protects the earth from hazardous ultraviolet radiation. Its depletion increases the rate of skin cancer in humans, can damage some plants, and may play a role in global warming | Equivalent of kilograms of trichlorofluromethane (Freon-11) |
| Emission of particulate matter | Airborne particulate matter is responsible mostly for respiratory problems in humans and animals and can cause a considerable amount of damage to plants | Change in mortality due to particulate matter emissions |
| Acidification | The acidification contributes to a decline of plant species like coniferous forests and increases fish mortality | Equivalent of moles hydron (mol H+ eq) |
| Terrestrial eutrophication | Terrestrial eutrophication affects all ecosystems. For example, it impacts strongly the lichen communities or forest tree health | Equivalent of moles of nitrogen (mol N eq) |
| Freshwater eutrophication | Freshwater eutrophication causes algae proliferation reducing the oxygen content in water. This reduced oxygen concentration can cause fish deaths. It also increases the drinking water treatment costs, reduces the recreational value of water bodies, and is a source of greenhouse gases | Equivalent of kilograms of phosphorus (kg P eq) |
| Marine eutrophication | Marine eutrophication causes algae proliferation. It affects the physiology and growth of marine organisms which has a cascading effect on the ecosystem functioning | Equivalent of kilograms of nitrogen (kg N eq) |
| Freshwater ecotoxicity | It refers to the potential toxic impacts on an ecosystem. | Comparative Toxic Unit for ecosystems (CTUe) a score based on a model called USEtox |
| Water use | The water use is related to the local scarcity of water | Cubic meters of water |
| Land use | Land use refers to the use and transformation of land | Loss of soil organic matter content in kilograms of carbon deficit (kg C deficit) |
| Fossils resource use | Use of non-renewable resources such as coal, oil, and gas expressed | MJ |
| Metals and minerals resource use | Use of metals and minerals resource | Equivalent of kilograms of antimony (kg Sb eq) |

**Supplemental Method 2 Pairing between INCA3 and the environmental indicators**

With Agribalyse and SHARP, 1610 out of 1761 foods eaten in INCA3 had GHGe values.

Some foods had no value because they were average foods (for example, “average bread”). Their GHGe value was assigned as the average value of the foods in the category weighted by the consumption of these foods in the population. This provided an additional 87 values.

When an average food existed in a category and some of the food of the category had no values, their value was taken as that of the average food.

Agribalyse used some proxies as the measurement of the indicator of some recipes. These proxies were used for two of the foods.

Finally, 46 of the foods had no value. The GHGe of individual diets were calculated using the food consumption survey and the GHGe production for each consumed food. To complete the missing data, all foods were classified into 45 categories. The missing data value took the mean GHGe production value of their category.

For the other environmental impacts, the same protocol was used, without the use of SHARP database (Supplemental Figure 2).
